# Stay-at-home and face mask policies intentions inconsistent with incidence and fatality during US COVID-19 pandemic

**DOI:** 10.1101/2020.10.25.20219279

**Authors:** Samuel X. Wu, Xin Wu

**Author notes:** Address correspondence to: Xin Wu, MD, MS, Department of Neuroscience and Experimental Therapeutics, College of Medicine, Texas A&M University Health Science Center, 2012 Medical Research and Education Building, 8447 Riverside Parkway, Bryan, TX 77807.

## Abstract

In addition to the United States CDC recommendations, many states imposed stay-at-home (SAH) and mandatory face mask (MFM) orders during COVID-19 pandemic. The purpose of this study was to characterize the relationship between SAH and MFM approaches with the incidence and fatality during the pandemic period until the 2020-08-23 (about 171 days). States with SAH orders showed potential decreases of infection and fatality during SAH period (about 45 days). However, many results in this study were inconsistent with the intent of public health strategies of SAH and MFM. There were similar incidence rates among SAH + MFM, SAH + no-MFM and no-SAH + no-MFM states. After normalized to population density, there were no significant differences in total positive cases, average daily new cases and average daily fatality among the 3 groups during the pandemic period. This study suggested that SAH and MFM orders in the general public alone, probably have limited effects in lowering transmission and fatality. From the policy-making level, if we cannot strictly isolate contagious patients with effective contact tracing, we presume that following the CDC recommendations could be appropriate in helping mitigate the COVID-19 disaster and limiting collateral social-economic damage with close monitoring of healthcare capacity.

## Introduction

Coronavirus disease 2019 (COVID-19), now recognized as a multi-system disease, is caused by severe acute respiratory syndrome coronavirus-2 (SARS-CoV-2), and has infected 50 million and killed more than a million people worldwide. Due to the lack of prophylactic and effective therapeutic approaches so far, non-pharmaceutical interventions are the only option for potentially mitigating pandemic transmission and fatality. It is believed to spread primarily from person-to-person mainly through close contact, utilizing the respiratory route ^1^. Community mitigation strategies following the United States Centers for Disease Control and Prevention (US, CDC) recommendations can lower the risk for disease transmission by limiting or preventing person-to-person interactions. The CDC recommends practicing hand hygiene, social distancing, wearing face coverings anywhere they will be around other people (especially when social distancing is difficult), monitoring personal health, and staying at home when sick. However, increased mask use in public during COVID-19, along with a global supply shortage, has led to widespread use of homemade masks and face covering alternatives ^2, 3, 4^. In conjunction with gowns, gloves, and eye protection, the mask is a core component of the personal protective equipment (PPE) that clinicians and researchers in infectious diseases fields require when caring for symptomatic patients. Although wearing a face mask has been controversial in the general public during influenza-like illness season, it is believed to reduce the spread of COVID-19 because asymptomatic positive patients can also spread the virus ^5, 6, 7, 8^. Many papers describing public health strategies either focus on masking models, indirect stay-at-home and masking data, or school closure ^9, 10, 11, 12, 13^. There is no direct investigation on how COVID-19 mitigation efforts at the policy making level affect infection and fatality on the entire US pandemic database with a total population basis.

Because the COVID-19 pandemic is asynchronous and varies in transmission across the US, states differ in whether or not they require their citizens to follow lockdown or stay-at-home (SAH) orders and/or mandated face masks (MFM) to limit COVID-19 spread. Apart from the CDC recommendations, individual states began implementing various community mitigation policies including SAH and MFM orders as of March 2020. The SAH and MFM orders can help reduce activities associated with community spread of COVID-19 by limiting or preventing person-to-person interactions outside the household ^9, 14, 15^.

Combating the pandemic will involve reducing both incidence rates and severity of the disease. However, direct investigations on how COVID-19 mitigation efforts affect its transmission (e.g. incidence rate) and severity (e.g. case-fatality-ratios (CFR)) are lacking. The purpose of this study was to characterize the relationship between mitigation strategies and COVID-19 consequences around the US.

## Results

### Trends in the daily new COVID-19 cases in the US decreased due to stay-at-home orders and increased following some social gathering events

The incubation period was defined as the interval between the potential earliest date of contact with the transmission source, and the potential earliest date of symptom onset or the time that most likely will be received by positive test results. The incubation period for COVID-19 was thought to be from 5 days extending to 14-23 days ^16, 17^. There were some cause and effect phenomena in this curve (Fig. 1). The curve trends were going up following spring break (late February to early April) and peaked on 2020-04-11 in the 7-day average curve. In Fig. 1, the daily new cases curve of the nation was flattened about 16 days after SAH (n=43 states) with the first peak date of 2020-04-11. 6 out of 43 SAH states had the peak (or first peak. Table 1 and Fig. 1) before the date of 2020-04-11. The second rise of the curve followed 23 days after ending the SAH order during a society phased reopening, and 8 days post-Memorial Day. The trends generally increased following nation-wide gathering events during May and June, and the July 4^th^ holiday. The most recent peak date (or second peak date if a state had already peaked) was about on 2020-07-23, 19 days post July 4^th^.

**Table 1.** Characteristics of daily new positive case curve trend and population information.

|  | SAH + MFM | SAH + no-MFM | No-SAH + no-MFM |
| --- | --- | --- | --- |
| # of States and DC | 34 | 9 | 7 |
| States with second peak | 13 | 0 * | 2 |
| Days to first peak (or most recent peak if only 1 peak of that state) | 97.82 ± 9.2 | 122.78 ± 9.66 | 121.85 ± 17.64 |
| How many states with peak before the first peak on the date of 20200411 in Fig. 1 | 6 | 0 | 0 |
| Days to most recent peak in Fig. 1 | 133.06 ± 7.93 | 122.78 ± 9.66 | 150.15 ± 5.67 & |
| Averaged Population density (residents/mile<sup>2</sup>) | 561.09 ± 308.13<br>(257.24 ± 52.78 if excluded DC<sup>##</sup>) | 129.93 ± 36.24 * | 28.43 ± 7.96*, & |

MFM: mandating face mask; SAH: Stay at home; DC: District of Columbia. \* $p < 0.05$ vs SAH + MFM; & $p < 0.05$ vs SAH + no-MFM. <sup>##</sup>: There was no significant difference among 3 groups if excluded DC population density in SAH + MFM states. Washington DC population density was included in all normalization in the results session. Each number is represented as the mean ± SEM. Population density data see supplemental eTable 1 and eTable 2.

**Fig. 1.**
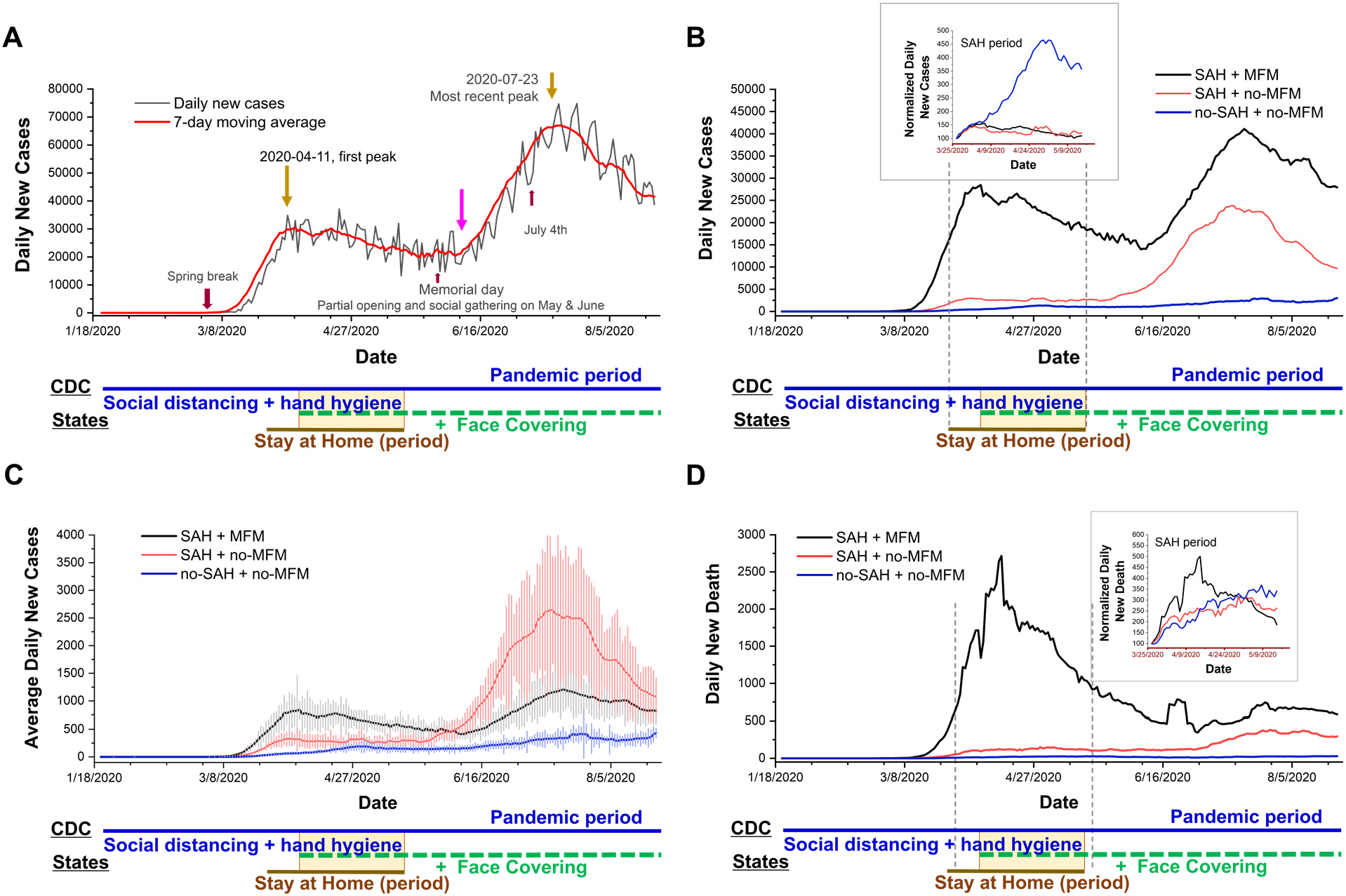
Daily Trends in Number of COVID-19 New Cases in the US Reported to CDC. Date is the time of COVID-19 pandemic period (averaged at 171 days): from case 1 in the USA to 2020-08-23. Daily cases collected from 50 states and Washington DC. (A) Daily new cases raw data and 7-day averaged data. The first peak was on 2020-04-11, the turning points for second rise was 2020-06-08 and second peak (i.e. most recent peak) was 2020-07-23 from 7 days average curve. All holidays or events are indicated in the figure. The overlapping between states mandating stay-at-home and mandating face masks is emphasized with a yellow rectangle box. (B) The daily new case curves of 7-day averaged were showed for mandating stay-at-home (SAH) + mandating face mask (MFM) states (n=34), SAH + no-MFM states (n=9), and no-SAH + no-MFM states (n=7) during the pandemic period. The curves were bending downward in the group of MFM + SAH states, and were flattened in the group of MFM + no-MFM states during SAH periods (averaged at 45 days). The upper inset indicated normalized daily case data to the case of day 1 of SAH (i.e. using same dates as in the SAH groups) which shown significant increased trend of new daily cases in the no-SAH + no-MFM states during SAH period. (C) The average daily new case curves were showed for the 3 groups of the states during pandemic periods. Each data represents the mean ± SEM. (D) The daily new death curves of 7-day averaged were showed for the 3 groups of the states. The upper inset indicated normalized daily new death data to the death number in day 1 of SAH which shown curve-bending downward in the SAH + MFM states during SAH period.

The daily new total and daily average case curves (Fig. 1B and 1C), and daily death curve (Fig. 1 D) have showed to bend downward in the group of MFM + SAH states (n=34), and was flattened in the group of MFM + no-MFM states (n=9) during SAH periods. Normalized daily case data to the case number in day 1 of SAH (i.e. using same dates as in the SAH groups) in the upper inset of Fig. 1B revealed significant increased trend of new daily cases in the no-SAH + no-MFM states (n=7). The SAH + MFM and SAH + no-MFM states showed 56.5% and 57.7% decrease in daily infection trends when comparing to no-SAH + no-MFM, respectively. There were similar daily infection trends between SAH + MFM and SAH + no-MFM states during SAH period. Normalized daily death to the death in day 1 of SAH in the inset of Figure 1D showed significant curve-bending downward in the SAH + MFM states. This downward trend could indicate 61.1% decrease in death numbers in the SAH + MFM states during SAH periods alone.

13 out of 34 SAH + MFM states and 2 out of 7 no-SAH + no-MFM states had two peaks. 0 out of 9 SAH + no-MFM states had a second peak (p=0.03 vs SAH + MFM. Table 1) during pandemic period. There was a significant difference in the number of days from the date of the first case in each state to the most recent peak between SAH + no-MFM states and no-SAH + no-MFM states (Table 1). Thus, these results indicated that SAH helped flatten the transmission and save the lives.

### States with mandatory stay-at-home orders and face masks policies have mixed results of daily positive cases and fatality during pandemic periods

To evaluate whether SAH and MFM are associated with prevention of infection and severity of disease, we investigated test positivity rate, incidence rate, mortality and CFR among each state group during the pandemic period (about 171 days until 2020-08-23). There was no significant difference in test percentage in the population of the states (24.01±1.48% in SAH + MFM states, 23.49 ± 3.21% in SAH + no-MFM states and 24.85±5.40% in no-SAH + no-MFM states (Fig. 2A). The SARS-CoV-2 polymerase chain reaction antigen testing positivity rate in SAH + no-MFM states was significantly higher compared to positivity rate in SAH + MFM states (p=0.03. Fig. 2B) and was not significantly different compared to positivity rate in no-SAH + no-MFM states. To compare the COVID-19 infected level, the SARS-CoV-2 antibody positivity rate from available studies was also examined^18^. The results indicated that there were no significant differences in antibody levels in sampled population among SAH + MFM (n=31), SAH + no-MFM (n=9), and no-SAH + no-MFM (n=7. Fig. 2C), respectively.

**Fig. 2.**
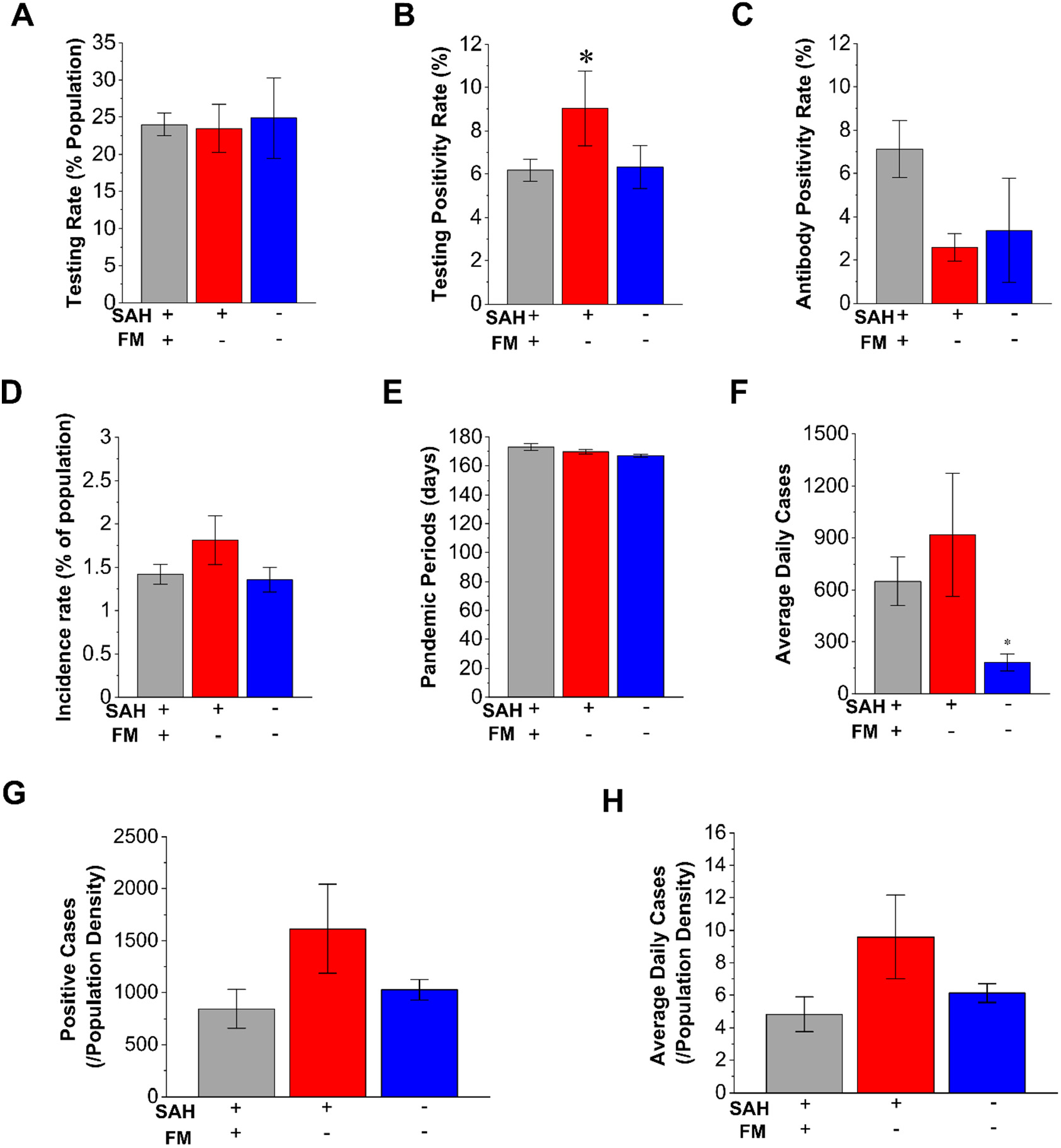
Characteristics of Mandating Face Masks (MFM) and Stay-at-Home (SAH) strategies in average daily cases and incidence rate of COVID-19. A. There is no significant difference in testing percentage of the state population among SAH + MFM (n=34 states), SAH + no-MFM (n=9 states), and no-SAH and no-MFM (n=7) states. B. There is a significantly higher testing positivity rate in SAH + no-MFM state when compared to SAH + MFM and no-SAH + no-MFM states. When compared to SAH + MFM, SAH + no-MFM has about 46% higher in positivity rate. There are no significant differences in antibody positivity rates (C), incidence rate (D) and periods of pandemic (E) among the three groups. F. The average daily positive cases are significantly higher in SAH + MFM and SAH + no-MFM states when compared to no-SAH + no-MFM states. There is no significant difference in total positive cases reported (G) and in average of daily positive cases (H) per state population density during the pandemic among SAH + MFM (Washington DC population density included), SAH + no-MFM, and no-SAH + no-MFM states. FM: face mask. Each bar represents the mean ± SEM. *p<0.05 vs SAH + MFM states.

There were no significant differences in the results of incidence rate among SAH + MFM states (95% CI of incidence rate, 1.19% to 1.64%. n=34), SAH + no-MFM states (95% CI, 1.26% to 2.36%. n=9) and no-SAH + no-MFM (95% CI, 1.08% to 1.63%. n=7) and no significant difference in the dates of pandemic periods (Fig. 2D and E). However, SAH + MFM (p=0.003) and SAH + no-MFM (p=0.07) states have a higher average of daily new cases compared to the no-SAH + no-MFM (Fig. 2F).

A geographic analysis of population density during the pandemic indicated that population density could be a factor that affects transmission and fatality ^19, 20^. The information regarding state population density (residents/mile^2^) have also been considered during analysis (Table 1). There was a significant difference in the population density in the groups of SAH + MFM, SAH + no-MFM, and no-SAH + no-MFM states (p<0.05, Table 1). The SAH + MFM states have a much higher population density when including Washington DC. We examined total positive cases and average daily new cases impacted by population density. There were no differences among total positive cases and the average daily positive cases when normalized with population density among the three groups (Fig. 2G and H).

When comparing case severity level, states with SAH + MFM (95% CI, 2.72% to 4.07%) had a 2.6-fold higher CFR compared to no-SAH + no-MFM states (95% CI, 1.08% to 3.16%. p=0.009. Fig. 3A). The fatality periods in states with no-SAH + no-MFM have 4.8% fewer days compared to SAH + MFM and SAH + no-MFM (p=0.017. Fig. 3B). Because longer fatality periods could result more death numbers, we also compared daily deaths among 3 groups. The states with SAH + MFM (p<0.001) or SAH + no-MFM (p=0.055) had 9.53-fold or 6.94-fold higher averages of daily death than no-SAH + no-MFM states, respectively. When total deaths were normalized with population (per 100,000 residents), as mortality rates, the SAH + MFM (3.0-fold, p<0.05) and SAH + no-MFM (2.0-fold) states had higher mortality rates than no-SAH + no-MFM states (Fig. 3D). When CFR was normalized by population density, the no-SAH + no-MFM states had a significantly higher CFR when compared to SAH + MFM states (p=0.033. Fig. 3E). However, there was no significant difference in average daily fatality per population density among the three groups (Fig. 3F). These results suggested that SAH and MFM/no-MFM orders provided mixed effects for prevention of infection and protection on severity of the disease.

**Fig. 3.**
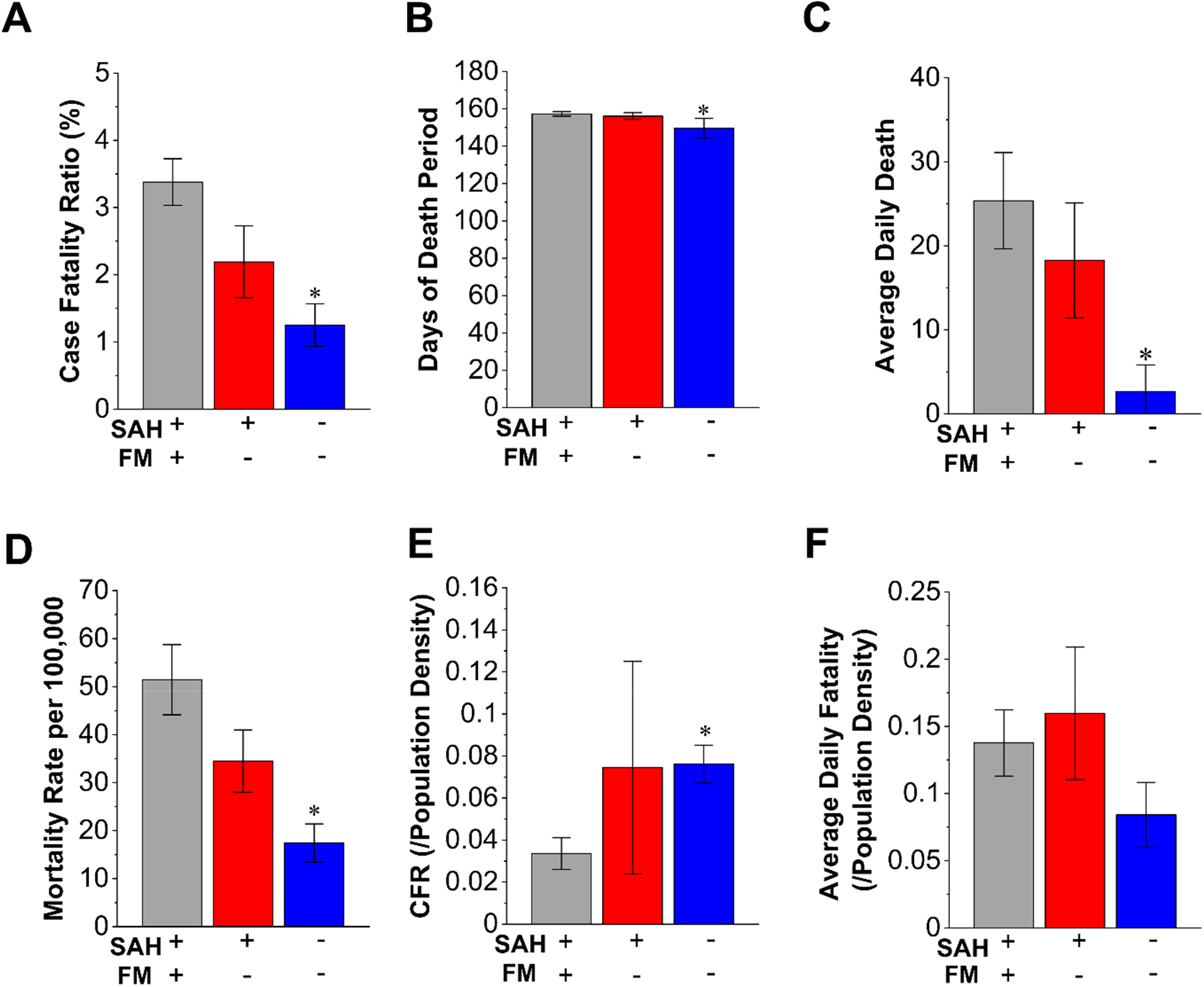
Characteristics of Mandating Face Masks and Stay-at-Home strategies in fatality of COVID-19. A. The case-fatality-ratio (CFR) is higher in SAH + MFM (2.7-fold, p<0.05, n=34) and SAH + no-MFM (1.7-fold, n=9) states when compared to no-SAH + no-MFM (n=7) states. B. Because the fatality periods from the first death in each state to 2020-08-23 might affect CFR, we examined the periods of 3 groups. There is about 5% fewer days when counting fatality in no-SAH + no-MFM than in SAH + MFM states. There is no significant difference in days of fatality periods between SAH + no-MFM and no-SAH + no-MFM states. C. Because there is a difference in the fatality periods between SAH + MFM and no-SAH + no-MFM, we examined the average daily death numbers among 3 groups. The average death number in no-SAH + no-MFM states has significantly fewer death than SAH + MFM and SAH + no-MFM states. D. The mortality rates per 100,000 population of SAH + MFM (3.0-fold, p<0.05) and SAH + no-MFM (2.0-fold) states are higher than mortality in the no-SAH + no-MFM sates. E. The CFR per state population density is significantly higher in no-SAH + no-MFM when compared to SAH + MFM states. No significant difference in CFR per population density between SAH + no-MFM states and SAH + MFM states. F. There is no significant difference in the average daily death cases reported per state population density during pandemic among SAH + MFM, SAH + no-MFM, and no-SAH + no-MFM states. Each bar represents the mean ± SEM. *p<0.05 vs SAH + MFM states.

The state of Arkansas (22.2% testing percentage) issued MFM without a SAH order, and was excluded in the 3 state groups above. Mixed data results were shown with an 8.4% antigen testing positivity rate and 1.87% incidence rate, which were similar to SAH + no-MFM states, and a 1.2% CFR that was similar to no-SAH + no-MFM states.

### States with MFM policies have not shown preventive effects on incidence and average daily cases of COVID-19 during mandatory SAH order periods

To examine whether SAH overlapping with MFM (yellow rectangle box in the lower part of Fig. 1) demonstrated better preventive effects, we examined incidence rate along with daily new cases during the SAH window (averaged about 45 days. n=43 states). The average overlapping periods for SAH and MFM was 28 ± 3 days, which was about 50% of overall SAH days. The incidence rate in MFM states (n=12) was significantly higher than no-MFM states (n=31. p=0.003. Fig. 4A). Because the number of SAH days in MFM states was 23% longer than no-MFM states (p=0.11. Fig. 4B) which could increase the positive cases data, we examined the average daily new cases between the two groups during the SAH periods. The average daily new positive cases in MFM states was 4-fold higher than the no-MFM states (p=0.004. Fig. 4C).

**Fig. 4.**
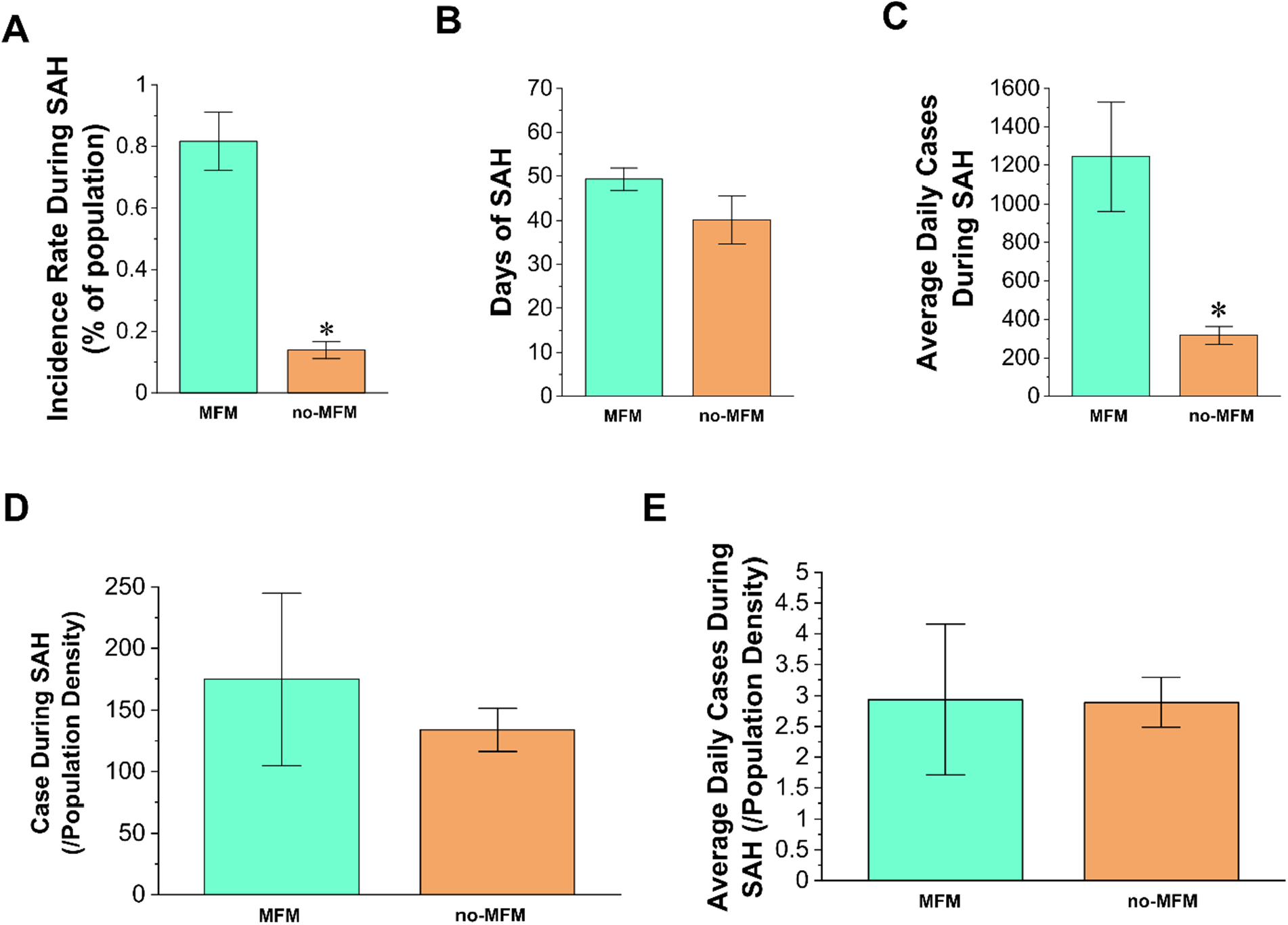
Characteristics of states overlapping with mandating masks in the transmission of COVID-19 during the periods of a mandatory stay at home (SAH) order. A. The infected cases in no-MFM states (n=31) have a significantly lower incidence rate than overlapping MFM states (n=12) with SAH order. B. There is no significant difference in SAH period between no-MFM and MFM states. C. The average daily positive cases in no-MFM states showed significantly fewer numbers than MFM states. There are no significant differences in total positive cases (D) and average daily cases (E) per state population density between MFM and no-MFM states during the periods of SAH. Each bar represents the mean ± SEM. *p<0.05 vs MFM states.

We examined total positive cases and average daily new cases impacted by population density. There were no significant differences in the number of total positive cases (Fig. 4D) and average daily cases (Fig. 4E) impacted by population density between MFM and no-MFM states. Thus, these results suggested that MFM policy during the SAH period may not have had an advantage over the no-MFM policy.

## Discussion

This study is the first report to evaluate all available COVID-19 positive eases and fatality data in the US, not utilizing theoretical modeling or selected regional databases, from policy-making levels during pandemic periods until date of 2020-8-23. Our results were inconsistent with the intent of public health strategies in lowering overall infection rates and fatality. This study provided direct evidence of decreased infection and fatality in SAH + MFM and SAH + no-MFM states during SAH periods (Fig. 1). There were also potential decrease in testing positivity rate when comparing SAH + MFM to SAH + no-MFM groups, and a decrease of fatality rate to potential save lives when normalized by population density through strategies of SAH + MFM orders during the COVID-19 pandemic (Fig. 2). However, no-SAH + no-MFM group had lower incidence rate, average daily news cases, CFR and mortality rates. There are no significant difference in SARS-CoV-2 antigen and antibody positivity rates among the 3 groups. When normalized to population density, there were no significant differences in total positive cases, averaged daily new cases and average daily fatalities among SAH + MFM, SAH + no-MFM and no-SAH + no-MFM state groups during pandemic periods. In addition, there was no difference in total positive cases and average daily new cases between MFM and no-MFM groups during SAH period after being normalized to population density.

Pathogenesis is the process by which an infection leads to asymptomatic or symptomatic signs of the disease. Theoretically, this process is always a balance between viral invasion (i.e., enough quality-virulence factors, enough quantity-virus load and enough contact time with contagious source-15 min as the CDC estimated) and human defense (i.e. physical and mental health). Most vaccines contain a small amount of attenuated or inactivated virus and help boost human immune defense system (preventive and protective efforts). What we can do from a prevention standpoint is to hinder the pathogen from reaching the sufficient virus load by using physical barriers such as face coverings and hand hygiene, and to avoid contact time through SAH, social distancing and reducing exposure to unknown crowds ^21, 22, 23^. Prolonged time in an indoor crowded environment (e.g. bars and restaurants) will increase your contact time with a possible contagious source. Human defense is the selection of a healthy lifestyle to boost an individual’s immune system to fight the virulent factors and to receive medical care in time as a last resort. Around the US, federal and state governments have been fighting the COVID-19 pandemic through a variety of strategies with the assistance of *predictive* modeling. These strategies include testing to find patients, isolating patients (*preemptive*), controlling transmission (*preventive*) and finally providing necessary clinical care when needed (*protective* and *personal care*).

The first step is to find the contagious source - patients. Testing is critical in the public health field’s mitigation efforts, helping investigators to characterize the incidence rate, spread and contagiousness of the disease. Early testing helps identify anyone who came into contact with an infected person so they too can be quickly quarantined or isolated and then treated if needed. However, the disadvantages for unjustified testing are potentially wasting limited amounts of resources and a delaying in the lab results for actual patients. The sooner patients receive test results, the sooner infected individuals can be isolated before they transmit their infection to others. Among all three with or without SAH and MFM states, the testing percentages (about 22%), SARS-CoV-2 antigen and antibody positivity rates, and total COVID-19 positive cases per population density had no significant differences during the pandemic period.

The second step of this strategy is mandatory isolation of patients and voluntary quarantine of close contactors followed by contact tracing and society support services, etc. This is the most important preemptive step because better control of a contagious source will greatly reduce transmission in the community, thereby reducing the positivity rate, incidence rate and fatality in the community. It has been reported that about 30%-62% patients were only partially isolated or not isolated at all ^24^. In addition, one big shortcoming of the US’ response to the coronavirus pandemic right now, at least in some parts of the country, involves a shortage of contact tracers and effectiveness of contact tracing ^25, 26, 27^. When new daily cases reach a certain level (such as 200/day in Houston Area, TX), it is very difficult to effectively trace and help isolate further spread of the virus (TMC in eTable 1). In Texas alone, thousands of more contact tracers are needed.

The third step of this strategy is preventive transmission control through the CDC recommendations of washing your hands often, maintaining good social distance, covering with a mask when around others, and monitoring your health daily. During influenza-like-illness season, including the COVID-19 outbreak before April, 2020, the CDC and World Health Organization (WHO) did not recommend face masks to the general public because there is no evidence from trials of their effectiveness in reducing transmission ^5, 28^. However, the CDC recommends using face masks in general public if you are sick, taking care of a sick patient, have underlying situations such as immunocompromised persons or have to be in a crowded environment. As COVID-19 continues its global spread, the WHO and CDC recommended that it was possible that one of the strategies in pandemic control, such as face masks, might at least help reduce the severity of disease and ensure that a greater proportion of new positive cases are asymptomatic infections ^23, 29, 30^. Furthermore, dismissing a low-cost intervention such as mass masking as ineffective because there is no evidence of effectiveness in clinical trials is potentially harmful ^31^. We believed SAH and MFM in the general public could help with mitigation, and even save lives during the COVID-19 pandemic as shown in part of our data in Figs. 1 to 3 with population density, and in the balance of pathogenesis as mentioned above ^32^. Several studies found a reduction of the spread of COVID-19 after stay-at-home, school closure and social distancing mandates were enacted in most cities and states ^9, 10, 11, 12, 13, 33, 34^. The national overall daily new cases curve was consistent with these reports. There were several significant cause and effect relationships in the daily case trend curve (Fig. 1). The daily new case and death curve trends were bent downward, indicated decreased transmission and fatality, following a mandated short-term SAH order. However, it is possible that economic and non-economic consequences of the long-term SAH orders outweigh the need to protect public health ^35, 36^. When US states began to phased reopen their economies and communities started gathering, the daily new cases observed an upward surge. This suggested the efficacy of short-term SAH and social distancing measures, and could influence future public health policy making ^12, 33^. However, the pandemic results indicated there was no significant difference among 3 groups in positivity rates and incidence rate during pandemic periods.

The impact of population density on emerging highly contagious infectious diseases has rarely been studied. In theory, dense areas over a certain threshold level lead to more close interactions among residents, which makes them potential hotspots for the rapid spread of pandemics. On the other hand, dense cities may have better access to healthcare systems ^19, 20^. Our research design for the current COVID-19 pandemic is a perfect population-based cross-sectional study to investigate these relationships. Our data indicated that the top 8 states in population densities are in the SAH + MFM group with high average daily new cases, mortality, CFR and average daily fatality. However, after being normalized with population density, there were no significant differences in daily new cases and average daily fatality among the 3 groups during pandemic periods (Figs. 2 and 3), as well as between MFM and no-MFM groups during SAH periods (Fig. 4). Many results in this study were inconsistent with the goals of preventive protocol, including SAH and MFM, to combat the pandemic and to reduce both infection rate and severity of disease. A limitation of this study was its focus on the difference of state-level policies. In order to find out why SAH and MFM orders did not show significant changes in infection and fatality, we may need analysis of individual county or city socioeconomic data, patient isolating data, contact tracing data, health care systems data, and law enforcement efforts during these mandating requirements (see limitations section).

Recent studies indicated that not all face masks have equal efficacy in reducing the transmission of particles or droplets, those most likely involved in COVID-19 people-to-people transmission ^2, 4^. Among all laboratory tested masks, fitted N95, used by health professionals who take care of COVID-19 patients with other necessary PPEs, perform best. Three-layer surgical masks, used by professionals in the hospital and clinical settings, also showed very good results. Some mask alternatives, such as neck fleeces or bandanas, offer very little protection ^2^. From a public policy perspective, during a pandemic similar to the COVID-19 pandemic, shortages in supply of surgical face masks and N95 respirators, as well as concerns about side effects and the discomfort of prolonged use from the masks, have led to public use of a variety of solutions which are generally less restrictive and usually of unknown efficacy (Table 2). In our opinion, if contagious patients do not strictly isolate themselves in the designated places and stay away from the rest of community, and if the general public is not informed of the correct types of masks (along with other PPE), proper methods to wear a mask, and knows how to properly social distance ^2, 4^, the community transmission will most likely be inevitable. The similar positivity rates of SARS-CoV-2 antigen and antibody, and incidence rate among 3 groups indicated compromised effectiveness of MFM and/or SAH strategies during pandemic periods^37^.

**Table 2.** Facts behind face mask: cost and effectiveness

|  | Issues | How to resolve |
| --- | --- | --- |
| Health<br>Economic<br>and Cost | Not everyone will be able to afford purchasing one-time-use disposable masks.<br>Worsened by global shortage of commercial supplies because production is based on society demands during non-pandemic.<br>Possible waste of natural resources and environments if universal masking in any situation in all states? | Reusable standardized cloth masks with daily washing because of the cost and availability <sup>44</sup>.<br>Homemade masks and face covering alternatives may not be as effective as commercially available masks. Schools and companies should provide free disposable masks during ILI season if they can provide free toilet papers and hand towels. Financial supports from Tax deduction & governments.<br>Stockpiles at home and FEMA |
| Compliance<br>/ Adherence | Feasibility and Adaptations. Very difficult for healthy people including children younger than 2 years old for a long-time use. Not everyone can wear because of health issues<sup>#</sup> | Culture difference: traditionally, Western societies consider only those infected should wear masks because of potential spreading the virus. In Asia, wearing a mask is not just mainly for protecting people from getting infected, but also minimizing the possibility of potential infection in you from spreading to other people. Education people that masks protect you and community. |
| Effectiveness | Works well if you know how to proper use face masks that meet the standards or approved by the FDA and NIOSH | Education on the proper use, disposal of used face masks, daily washing for cloth mask<sup>44</sup>, and hand hygiene (example: washing hands before touching mask; washing hands before and after disposing of mask, etc.). |
| Adverse<br>effects | **Ignoring social distancing and hand hygiene.**<br>Symptoms: Skin breakdown, acne, impaired cognition, anxiety, headaches, trigger skin sensitivity, eczema, acne, shortness of breath, and **mask mouth** (bad breath, dental problems, Yeast and bacterial infections).<br>Society security and safety issues | Most symptomatic side effects go away once mask is removed, otherwise seek professional care. |

ILI: influenza-like illness; FEMA: The Federal Emergency Management Agency. NIOSH: The National Institute for Occupational Safety and Health. #: reference in CDC3 in supplemental eTable 1.

The fourth and last step of this strategy is to provide protective clinical care and prevent the overwhelming of the healthcare system during the pandemic. Hospitals and other healthcare facilities play a critical role in national and local responses to COVID-19. Currently, only one antiviral treatment, Remdesivir, is approved with debatable benefit-risk assessment on 2020-10-20 by United States Food and Drug Administration ^38, 39^, and there is no vaccine available. The lone healthcare strategies remain supportive care solutions and the ability to manage underlying conditions (protective and personalized care). When mitigation steps are compromised, as our data from COVID-19 pandemic has indicated, the final step is to ensure we have sufficient healthcare capacity, especially intensive care unit (ICU) capacity. COVID-19 patients need rooms with negative pressure (e.g. ICU) to prevent contamination to the outside and need ventilators because of the respiratory issues associated with the disease. How do we define the hospital capacity with society safety threshold and tightening threshold needed to handle a potential outbreak demand during phased reopening ^33^? The CDC does not have clear recommendations yet. In Texas, if COVID-19 patients made up less than 15% of all hospitalizations, an increase in reopening capacity every 14 days was allowed (Texas reopening in eTable 1). The hospital capacity data in Figure 5 indicates that we currently have sufficient healthcare capacity to take care of the COVID-19 patients in the majority of the nation if based on Texas’s 15% suggestion, and in the worst scenario elective surgery can be suspended. In addition, there are 900,000 active licensed physicians including at least 70,000 physicians of emergency room, intensive care, and respiratory specialists ^40, 41^. Moreover, there are 3 million nurses and 129,000 respiratory therapists available in the US (Fig. 5). The mitigation steps might be appropriately tightened or loosened to minimize social and economic disruption following the balance among patient’s privacy, vulnerable populations’ safety, people’s freedom, and law enforcement following hospital capacity changes.

**Fig. 5.**
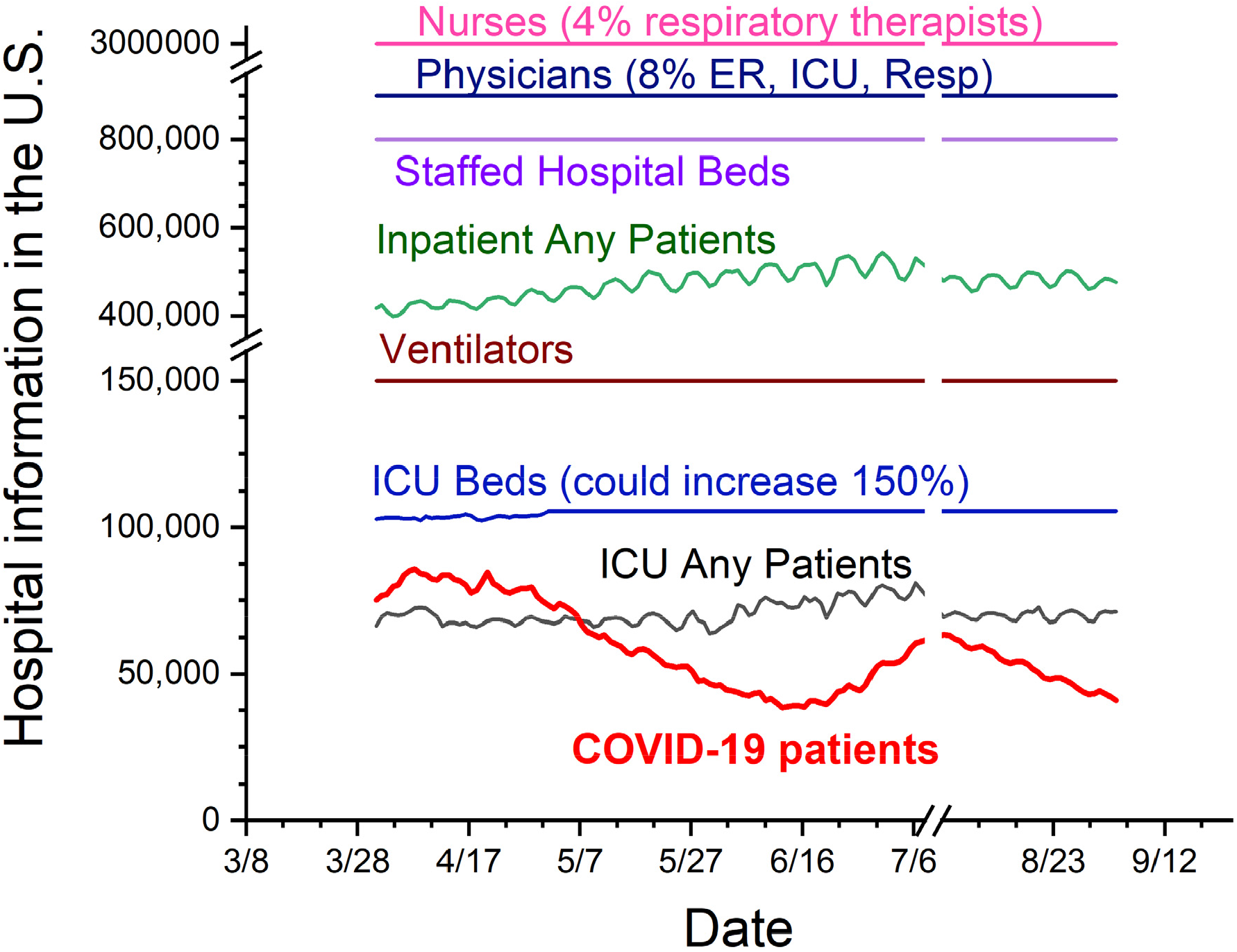
US hospital capacity during COVID-19 pandemic. The periods are from the date of 2020-04-01 to 2020-09-04. The chart includes numbers of physicians, nurses, hospital beds, all hospitalized patients, ventilators, intensive care unit (ICU) beds, all ICU patients, and all COVID-19 patients. From the numbers in this chart, it is possible that individual hospitals might have shortage of beds or respiratory therapists. However, we believed that there are sufficient hospital capacities for COVID-19 patients during the pandemic period.

### Limitations

This study has several limitations. First, these results were limited to the state policy levels. We did not consider data on statewide school closure, nonessential business closure and a county-level MFM orders in no-MFM states, such as those in the state of Florida. Second, we only examined confirmed COVID-19 provisional cases and reported death cases until 2020-08-23. The reported cases and fatality data can lag. There is evidence by the CDC of a higher infection rate in the community than what is reflected in the number of confirmed COVID-19 cases, especially with asymptomatic patients. In addition, we did not examine the percentage of patients who died with do-not-resuscitate status that could be responsible for increased mortality. Third, we did not examine socioeconomic and demographic factors (e.g. age, sex and education level) between states. Socioeconomic conditions or cultural preferences may result in many people living in a crowding space. Fourth, we did not measure enforcement of the mandates, which might affect compliance. Fifth, we also did not investigate the percentage of patients in nursing homes and assisted living facilities. Those with higher proportions of minority residents are more likely to have more COVID-19 cases ^42, 43^. Sixth, because COVID-19 is recognized as an acute disease, when calculating testing percentage, incidence and mortality rate, we presumed the people or patients only count once and that the population in that state will remain constant, even though people pass away and babies are born during the pandemic periods.

## Conclusion

This study provided direct evidence of potential decreases of infection and fatality with (short-term) SAH order, decrease in testing positivity rates, and decrease in fatality rate to save lives when normalized by population density through strategies of SAH + MFM order during the US COVID-19 pandemic. However, overall, many results in this study were inconsistent with the intent of public health strategies of short-term SAH and long-term MFM orders in lowering transmission and fatality. As states begin to reopen, we are considering the possibility of a uphold wave of the COVID-19 or COVID-19 combined with regular flu. From the policy-making level, if we cannot strictly isolate contagious source patients in separate isolated places and cannot implement effective massive contact tracing, we presume that following the CDC recommendations with sufficient testing could be appropriate in helping mitigate the COVID-19 disaster and limiting collateral social-economic damage with close monitoring of healthcare capacity. With the world facing an unprecedented threat, we must learn the lessons of this pandemic now, and ensure that our response is based on vulnerable patients, people’s freedom, community risk, and hospital capacity to make the world a safer place in the future.

## Methods

### Study data

Data on COVID-19 cases were collected from CDC provisional counts of United States COVID-19 cases and deaths by states over time (CDC 1 and CDC 2. Supplemental eTable 1, eTable 3, eTable 4). On 2020-08-23, there were more than 5-million positive cases and close to 170,000 fatalities among 330 million vulnerable populations from the 50 states and the District of Columbia (DC) in the USA. Other data sources, such as state health departments and the Johns Hopkins Coronavirus Resource Center were examined as reference and listed in Supplemental eTable 1 and eTable 2. The hospital capacity and health professional data were collected from American Hospital Association and Association of American Medical Colleges ^9, 40^. Data on state and territorial mandatory SAH and MFM orders for the general public were obtained from government websites containing executive or administrative orders or from press releases (supplemental eTable 1) ^9^. Between 2020-01-21 (first positive case in the US) and 2020-08-23, governors of 35 states and the mayor of Washington DC signed orders mandating all individuals who can medically tolerate the wearing of a face mask to do so in public settings (e.g. public transportation, parks and grocery stores). This practice applies both indoors and outdoors where maintaining six feet of social distancing might not always be possible. If there is no state-wide mandate (e.g. only for store employees, or only a couple counties or cities in the states), we will recognize these states as no-MFM (n=16 states). New Hampshire was counted as a non-MFM state because face coverings are only required for all persons who attend scheduled gatherings for social, spiritual, and recreational activities of 100 people or more. Between 2020-01-21 and 2020-08-23, 43 states and Washington DC signed orders mandating citizens to stay-at-home, or shelter-in-place (see eTable 1 and eTable 2 for references on MFM and SAH state list) ^9, 10^.

### Experimental Approach and Experimental Outcomes

The research design is a US population-based cross-sectional study. This study was designed to evaluate the efficacy of SAH and MFM at the policy-making level during the pandemic. The overall experimental protocol is illustrated in Fig. 1. We primarily organized all the states and DC into SAH + MFM states, SAH + no-MFM states, and no-SAH + no-MFM states. The two primary outcomes examined are incidence and fatality. To examine incidence, incidence rate, positivity rate and average daily cases with or without normalization to population density were investigated. To examine fatality, mortality rate (per 100,000 residents), CFR and average daily death with or without normalization to population density were investigated. Because COVID-19 is recognized as an acute disease, incidence rate or infection rate refers to the occurrence of new cases of COVID-19 in a state’s vulnerable population over a specified pandemic period. *The pandemic period* is from the date of the first positive case in each state to the date of 2020-08-23. To examine severity of disease, CFR was investigated over the *fatality periods* during the pandemic periods from the date of the first death in each state to 2020-08-23. The state-level COVID-19 testing rates, SARS-CoV-2 polymerase chain reaction antigen and antibody positivity rates were also examined.

### Statistical Analysis

We employed an event study, which is generally similar to a difference in-differences design, to examine whether statewide mandated SAH and/or the use of face masks in public affect the spread of COVID-19 based on the state variations noted earlier. This design allowed us to estimate these effects in the context of a natural experiment, comparing the changes in COVID-19 spread among the states with mandates to changes in COVID-19 spread in the states that did not pass these mandates, over a period of time.

Statistical tests were performed using SAS software (SAS Institute Inc., Cary, NC) and Origin 2020b (OriginLab Corporation, Northampton, MA). Nonparametric outcomes, such as two-peak occurrence, were compared between groups using the Chi-square Test (or Fisher Exact Test). Statistical comparisons of incidence rate and CFR were performed with one-way or two-way analysis of variance (ANOVA) as appropriate. Post hoc analyses were carried out to identify specific differences using Tukey’s HSD (honestly significant difference) for multiple comparisons. In all statistical tests, differences were considered statistically significant at 2-sided p<0.05. Data was expressed as the mean ± SEM.

## Data Availability

All data are public available and with information in the supplemental.

## Authors’ contributions

SW and XW conceived the study and collected the data. SW and XW searched the literature. SW and XW wrote the manuscript. SW and XW analyzed the data. All authors were involved in data interpretation and made meaningful contributions to the final submitted manuscript.

## Competing Interest Statement

The authors declare no competing interests.

## Funding/Support

not applicable

## Data sharing

All data are public available and with information in supplemental eTable 1 to eTable 4.

**Correspondence and requests for materials should be addressed to Xin Wu**

## Notes

### Competing Interest Statement

The authors have declared no competing interest.

### Summary of Updates

(1) Figure 1 updated with analysis of 3 groups of the states separately. That provided direct evidence of potential decreases of infection and fatality with (short-term) SAH order. (2) Figure 5 updated with physicians and nurses data. (3) Mortality data also included in the results. Some updated in discussion section. (4) Supplemental files updated.

