## Supplemental data for "Stay-at-home and face mask policies intentions inconsistent with incidence and fatality during US COVID-19 pandemic"

eTable 1. Public reports reference other than papers and books page 2

eTable 2. Date of stay at home and mandatory face mask orders Page 3

eTable 3. COVID-19 cases until 2020-08-23 pages 4-14

eTable 4. COVID-19 death until 2020-08-23 Pages 15-25

**Supplemental 1**

Supplemental eTable 1: Public reports reference other than papers and books

| |
|---|
| **Case and fatality data:**<br>CDC1: Implementation of mitigation strategies for community COVID-19 transmission. Atlanta, GA: US. Department of Health and Human Services, CDC; 2020. https://www.cdc.gov/coronavirus/2019-ncov/ community/community-mitigation.html<br>CDC2: https://data.cdc.gov/Case-Surveillance/United-States-COVID-19-Cases-and-Deaths-by-State-o/9mfq-cb36<br>CDC3: Considerations for Wearing Masks Help Slow the Spread of COVID-19 (who should/should not wear a mask) https://www.cdc.gov/coronavirus/2019-ncov/prevent-getting-sick/cloth-face-cover-guidance.html.<br>HealthData.gov-link: https://healthdata.gov/dataset/covid-19-state-and-county-policy-orders<br>Johns Hopkins Coronavirus Resource Center: https://coronavirus.jhu.edu/us-map<br>The Covid-19 Tracking Project: https://covidtracking.com/data#TX<br>Our World in Data: https://ourworldindata.org/ |
| **Hospital and health professional data:**<br>AHA (hospital data) : https://www.aha.org/statistics/fast-facts-us-hospitals<br>Nurse workforce in the US: https://www.nursingworld.org/practice-policy/workforce/<br>Respiratory Therapists: https://www.bls.gov/ooh/healthcare/respiratory-therapists.htm<br>AAMC physician data: https://www.aamc.org/data-reports/workforce/interactive-data/active-physicians-us-doctor-medicine-us-md-degree-specialty-2015 |
| **Stay at home and face masking order:**<br>USA Today: Coronavirus reopening. Map of COVID-19 case trends, reopening status and mobility, 20200829: https://www.usatoday.com/storytelling/coronavirus-reopening-america-map/<br>See Which States and Cities Have Told Residents to Stay at Home - The New York Times: https://www.nytimes.com/interactive/2020/us/coronavirus-stay-at-home-order.html<br>Face mask: https://www.littler.com/publication-press/publication/facing-your-face-mask-duties-list-statewide-orders<br>Face mask: https://www.aarp.org/health/healthy-living/info-2020/states-mask-mandates-coronavirus.html<br>Face masks: https://masks4all.co/what-states-require-masks/ |
| **Population density** based on data from the 2013 estimate of population by the United States Census Bureau: https://simple.wikipedia.org/wiki/List_of_U.S._states_by_population_density |
| **Texas reopening:** Gov. Greg Abbott loosens coronavirus restrictions for restaurants and other businesses in most regions of Texas: https://www.texastribune.org/2020/09/17/greg-abbott-texas-coronavirus/ |
| Three Metrics To Gauge Our Progress, Coronavirus (COVID-19) Updates. Texas Medical Center **(TMC)**. https://www.tmc.edu/coronavirus-updates/three-metrics-to-gauge-our-progress-2/ |

**eTable 2. Data of date of stay at home and mandatory face mask orders**

| States | population in 2019 | population density 2013 (/mile2) | current date | date of mask order | stay at home starting date | stay at home ending date | COVID-19 antigen testing number until 8/23/2020 | Date of the first peak of the new daily positive (7 day average) | date of most recent peak of the new daily positive (7 day average) |
|---|---|---|---|---|---|---|---|---|---|
| .Alabama | 4,903,185 | 95.40 | 8/23/2020 | 7/15/2020 | 4/4/2020 | 4/30/2020 | 916,703 | 7/24/2020 | 7/24/2020 |
| .Alaska | 731,545 | 1.30 | 8/23/2020 | | 3/11/2020 | 4/21/2020 | 327,440 | 8/1/2020 | 8/1/2020 |
| .Arizona | 7,278,717 | 58.30 | 8/23/2020 | | 3/30/2020 | 5/15/2020 | 1,149,287 | 6/1/2020 | 6/1/2020 |
| .Arkansas | 3,017,804 | 57.00 | 8/23/2020 | 7/20/2020 | | | 670,826 | 7/24/2020 | 7/24/2020 |
| .California | 39,512,223 | 246.10 | 8/23/2020 | 6/18/2020 | 3/19/2020 | 5/4/2020 | 10,541,031 | 7/29/2020 | 7/29/2020 |
| .Colorado (2 peaks) | 5,758,736 | 50.80 | 8/23/2020 | 7/17/2020 | 3/26/2020 | 5/8/2020 | 930,152 | 4/25/2020 | 7/26/2020 |
| .Connecticut | 3,565,287 | 742.60 | 8/23/2020 | 4/20/2020 | 3/23/2020 | 5/20/2020 | 1,020,328 | 4/21/2020 | 4/21/2020 |
| .Delaware (2 peaks) | 973,764 | 475.10 | 8/23/2020 | 4/28/2020 | 3/24/2020 | 5/15/2020 | 221,845 | 4/27/2020 | 8/14/2020 |
| .District of Columbia (2 peaks) | 705,749 | 10588.00 | 8/23/2020 | 5/16/2020 | 4/1/2020 | 5/29/2020 | 266,046 | 5/2/2020 | 7/22/2020 |
| .Florida | 21,477,737 | 364.60 | 8/23/2020 | | 3/20/2020 | 4/30/2020 | 6,016,043 | 7/12/2020 | 7/12/2020 |
| .Georgia | 10,617,423 | 173.70 | 8/23/2020 | | 4/3/2020 | 4/30/2020 | 2,171,884 | 7/24/2020 | 7/24/2020 |
| .Hawaii | 1,415,872 | 218.60 | 8/23/2020 | 4/20/2020 | 3/25/2020 | 5/31/2020 | 228,381 | 8/14/2020 | 8/14/2020 |
| .Idaho | 1,787,065 | 19.50 | 8/23/2020 | | 3/25/2020 | 4/30/2020 | 235,255 | 7/16/2020 | 7/16/2020 |
| .Illinois (2 peaks) | 12,671,821 | 232.00 | 8/23/2020 | 5/1/2020 | 3/21/2020 | 5/30/2020 | 3,704,036 | 5/13/2020 | 8/21/2020 |
| .Indiana (2 peaks) | 6,732,219 | 183.40 | 8/23/2020 | 7/27/2020 | 3/25/2020 | 5/1/2020 | 1,273,212 | 4/26/2020 | 8/23/2020 |
| .Iowa | 3,155,070 | 55.30 | 8/23/2020 | | | | 595,765 | 8/23/2020 | 8/23/2020 |
| .Kansas | 2,913,314 | 35.40 | 8/23/2020 | 7/3/2020 | 3/30/2020 | 5/3/2020 | 366,315 | 8/23/2020 | 8/23/2020 |
| .Kentucky | 4,467,673 | 111.30 | 8/23/2020 | 5/11/2020 | 3/26/2020 | 5/11/2020 | 1,763,838 | 8/13/2020 | 8/13/2020 |
| .Louisiana (2 peaks) | 4,648,794 | 107.10 | 8/23/2020 | 7/13/2020 | 3/23/2020 | 5/14/2020 | 1,763,838 | 4/3/2020 | 8/3/2020 |
| .Maine | 1,344,212 | 43.10 | 8/23/2020 | 5/1/2020 | 4/2/2020 | 5/31/2020 | 231,256 | 5/21/2020 | 5/21/2020 |
| .Maryland (2 peaks) | 6,045,680 | 610.80 | 8/23/2020 | 4/18/2020 | 3/30/2020 | 5/15/2020 | 1,772,656 | 5/20/2020 | 8/1/2020 |
| .Massachusetts | 6,892,503 | 858.00 | 8/23/2020 | 5/6/2020 | 4/24/2020 | 5/18/2020 | 2,064,529 | 4/24/2020 | 4/24/2020 |
| .Michigan (2 peaks) | 9,986,857 | 175.00 | 8/23/2020 | 7/10/2020 | 3/24/2020 | 6/5/2020 | 2,564,334 | 4/2/2020 | 8/1/2020 |
| .Minnesota (2 peaks) | 5,639,632 | 68.10 | 8/23/2020 | 7/25/2020 | 3/27/2020 | 5/4/2020 | 1,374,536 | 5/23/2020 | 7/26/2020 |
| .Mississippi | 2,976,149 | 63.70 | 8/23/2020 | 8/4/2020 | 4/3/2020 | 5/11/2020 | 530,974 | 7/29/2020 | 7/29/2020 |
| .Missouri | 6,137,428 | 87.90 | 8/23/2020 | | 4/6/2020 | 5/3/2020 | 1,218,097 | 7/29/2020 | 7/29/2020 |
| .Montana | 1,068,778 | 7.00 | 8/23/2020 | 7/16/2020 | 3/26/2020 | 4/24/2020 | 210,749 | 7/30/2020 | 7/30/2020 |
| .Nebraska (2 peaks) | 1,934,408 | 24.30 | 8/23/2020 | | | | 338,836 | 4/30/2020 | 7/31/2020 |
| .Nevada | 3,080,156 | 25.40 | 8/23/2020 | 6/24/2020 | 3/31/2020 | 5/15/2020 | 809,683 | 7/17/2020 | 7/17/2020 |
| .New Hampshire | 1,359,711 | 147.80 | 8/23/2020 | | 3/27/2020 | 6/15/2020 | 301,316 | 5/2/2020 | 5/2/2020 |
| .New Jersey | 8,882,190 | 1210.10 | 8/23/2020 | 4/8/2020 | 3/21/2020 | 6/15/2020 | 2,648,740 | 4/4/2020 | 4/4/2020 |
| .New Mexico | 2,096,829 | 17.20 | 8/23/2020 | 5/16/2020 | 3/24/2020 | 5/15/2020 | 704,955 | 7/28/2020 | 7/28/2020 |
| .New York | 19,453,561 | 417.00 | 8/23/2020 | 4/17/2020 | 3/22/2020 | 5/15/2020 | 7,621,159 | 4/9/2020 | 4/9/2020 |
| .North Carolina (2 peaks) | 10,488,084 | 202.60 | 8/23/2020 | 6/26/2020 | 3/30/2020 | 5/8/2020 | 2,078,472 | 7/23/2020 | 8/21/2020 |
| .North Dakota | 762,062 | 10.50 | 8/23/2020 | | | | 432,725 | 8/23/2020 | 8/23/2020 |
| .Ohio | 11,689,100 | 283.20 | 8/23/2020 | 7/23/2020 | 3/23/2020 | 5/30/2020 | 1,977,822 | 7/30/2020 | 7/30/2020 |
| .Oklahoma | 3,956,971 | 56.10 | 8/23/2020 | | | | 821,289 | 7/27/2020 | 7/27/2020 |
| .Oregon | 4,217,737 | 40.90 | 8/23/2020 | 7/1/2020 | 3/23/2020 | 5/15/2020 | 510,056 | 7/24/2020 | 7/24/2020 |
| .Pennsylvania (2 peaks) | 12,801,989 | 285.50 | 8/23/2020 | 4/19/2020 | 3/30/2020 | 5/15/2020 | 1,549,379 | 4/9/2020 | 7/28/2020 |
| .Rhode Island (2 peaks) | 1,059,361 | 1017.10 | 8/23/2020 | 5/8/2020 | 3/28/2020 | 5/8/2020 | 470,162 | 4/24/2020 | 8/6/2020 |
| .South Carolina | 5,148,714 | 158.80 | 8/23/2020 | | 4/7/2020 | 5/12/2020 | 891,006 | 7/11/2020 | 7/11/2020 |
| .South Dakota (2 peaks) | 884,659 | 11.50 | 8/23/2020 | | | | 176,843 | 5/9/2020 | 8/23/2020 |
| .Tennessee | 6,829,174 | 157.50 | 8/23/2020 | | 4/2/2020 | 4/30/2020 | 2,042,256 | 7/27/2020 | 7/27/2020 |
| .Texas | 28,995,881 | 101.20 | 8/23/2020 | 7/3/2020 | 4/2/2020 | 4/30/2020 | 4,668,028 | 7/16/2020 | 7/16/2020 |
| .Utah | 3,205,958 | 35.30 | 8/23/2020 | | | | 766,985 | 7/17/2020 | 7/17/2020 |
| .Vermont | 623,989 | 68.00 | 8/23/2020 | 8/1/2020 | 3/24/2020 | 5/15/2020 | 120,043 | 4/4/2020 | 4/4/2020 |
| .Virginia (2 peaks) | 8,535,519 | 209.20 | 8/23/2020 | 5/29/2020 | 3/30/2020 | 6/10/2020 | 1,466,033 | 5/26/2020 | 8/8/2020 |
| .Washington | 7,614,893 | 104.90 | 8/23/2020 | 6/26/2020 | 3/23/2020 | 5/4/2020 | 1,379,036 | 7/13/2020 | 7/13/2020 |
| .West Virginia | 1,792,147 | 77.10 | 8/23/2020 | 6/6/2020 | 3/24/2020 | 5/4/2020 | 396,018 | 7/24/2020 | 7/24/2020 |
| .Wisconsin | 5,822,434 | 106.00 | 8/23/2020 | 8/1/2020 | 3/25/2020 | 5/26/2020 | 1,185,611 | 7/24/2020 | 7/24/2020 |
| .Wyoming | 578,759 | 6.00 | 8/23/2020 | | | | 93,231 | 7/23/2020 | 7/23/2020 |

**eTable 3. COVID-19 cases**

| Date/total cases | 1/21/2020 | 1/22/2020 | 1/23/2020 | 1/24/2020 | 1/25/2020 | 1/26/2020 | 1/27/2020 | 1/28/2020 | 1/29/2020 | 1/30/2020 | 1/31/2020 | 2/1/2020 | 2/2/2020 | 2/3/2020 | 2/4/2020 | 2/5/2020 | 2/6/2020 | 2/7/2020 |
|---|---|---|---|---|---|---|---|---|---|---|---|---|---|---|---|---|---|---|
| AK | 0 | 0 | 0 | 0 | 0 | 0 | 0 | 0 | 0 | 0 | 0 | 0 | 0 | 0 | 0 | 0 | 0 | 0 |
| AL | 0 | 0 | 0 | 0 | 0 | 0 | 0 | 0 | 0 | 0 | 0 | 0 | 0 | 0 | 0 | 0 | 0 | 0 |
| AR | 0 | 0 | 0 | 0 | 0 | 0 | 0 | 0 | 0 | 0 | 0 | 0 | 0 | 0 | 0 | 0 | 0 | 0 |
| AZ | 0 | 0 | 0 | 0 | 0 | 1 | 1 | 1 | 1 | 1 | 1 | 1 | 1 | 1 | 1 | 1 | 1 | 1 |
| CA | 0 | 0 | 0 | 0 | 0 | 2 | 2 | 2 | 2 | 2 | 3 | 3 | 3 | 6 | 6 | 6 | 6 | 6 |
| CO | 0 | 0 | 0 | 0 | 0 | 0 | 0 | 0 | 0 | 0 | 0 | 0 | 0 | 0 | 0 | 0 | 0 | 0 |
| CT | 0 | 0 | 0 | 0 | 0 | 0 | 0 | 0 | 0 | 0 | 0 | 0 | 0 | 0 | 0 | 0 | 0 | 0 |
| DC | 0 | 0 | 0 | 0 | 0 | 0 | 0 | 0 | 0 | 0 | 0 | 0 | 0 | 0 | 0 | 0 | 0 | 0 |
| DE | 0 | 0 | 0 | 0 | 0 | 0 | 0 | 0 | 0 | 0 | 0 | 0 | 0 | 0 | 0 | 0 | 0 | 0 |
| FL | 0 | 0 | 0 | 0 | 0 | 0 | 0 | 0 | 0 | 0 | 0 | 0 | 0 | 0 | 0 | 0 | 0 | 0 |
| GA | 0 | 0 | 0 | 0 | 0 | 0 | 0 | 0 | 0 | 0 | 0 | 0 | 0 | 0 | 0 | 0 | 0 | 0 |
| HI | 0 | 0 | 0 | 0 | 0 | 0 | 0 | 0 | 0 | 0 | 0 | 0 | 0 | 0 | 0 | 0 | 0 | 0 |
| IA | 0 | 0 | 0 | 0 | 0 | 0 | 0 | 0 | 0 | 0 | 0 | 0 | 0 | 0 | 0 | 0 | 0 | 0 |
| ID | 0 | 0 | 0 | 0 | 0 | 0 | 0 | 0 | 0 | 0 | 0 | 0 | 0 | 0 | 0 | 0 | 0 | 0 |
| IL | 0 | 0 | 0 | 1 | 1 | 1 | 1 | 1 | 1 | 1 | 2 | 2 | 2 | 2 | 2 | 2 | 2 | 2 |
| IN | 0 | 0 | 0 | 0 | 0 | 0 | 0 | 0 | 0 | 0 | 0 | 0 | 0 | 0 | 0 | 0 | 0 | 0 |
| KS | 0 | 0 | 0 | 0 | 0 | 0 | 0 | 0 | 0 | 0 | 0 | 0 | 0 | 0 | 0 | 0 | 0 | 0 |
| KY | 0 | 0 | 0 | 0 | 0 | 0 | 0 | 0 | 0 | 0 | 0 | 0 | 0 | 0 | 0 | 0 | 0 | 0 |
| LA | 0 | 0 | 0 | 0 | 0 | 0 | 0 | 0 | 0 | 0 | 0 | 0 | 0 | 0 | 0 | 0 | 0 | 0 |
| MA | 0 | 0 | 0 | 0 | 0 | 0 | 0 | 0 | 0 | 0 | 0 | 1 | 1 | 1 | 1 | 1 | 1 | 1 |
| MD | 0 | 0 | 0 | 0 | 0 | 0 | 0 | 0 | 0 | 0 | 0 | 0 | 0 | 0 | 0 | 0 | 0 | 0 |
| ME | 0 | 0 | 0 | 0 | 0 | 0 | 0 | 0 | 0 | 0 | 0 | 0 | 0 | 0 | 0 | 0 | 0 | 0 |
| MI | 0 | 0 | 0 | 0 | 0 | 0 | 0 | 0 | 0 | 0 | 0 | 0 | 0 | 0 | 0 | 0 | 0 | 0 |
| MN | 0 | 0 | 0 | 0 | 0 | 0 | 0 | 0 | 0 | 0 | 0 | 0 | 0 | 0 | 0 | 0 | 0 | 0 |
| MO | 0 | 0 | 0 | 0 | 0 | 0 | 0 | 0 | 0 | 0 | 0 | 0 | 0 | 0 | 0 | 0 | 0 | 0 |
| MS | 0 | 0 | 0 | 0 | 0 | 0 | 0 | 0 | 0 | 0 | 0 | 0 | 0 | 0 | 0 | 0 | 0 | 0 |
| MT | 0 | 0 | 0 | 0 | 0 | 0 | 0 | 0 | 0 | 0 | 0 | 0 | 0 | 0 | 0 | 0 | 0 | 0 |
| NC | 0 | 0 | 0 | 0 | 0 | 0 | 0 | 0 | 0 | 0 | 0 | 0 | 0 | 0 | 0 | 0 | 0 | 0 |
| ND | 0 | 0 | 0 | 0 | 0 | 0 | 0 | 0 | 0 | 0 | 0 | 0 | 0 | 0 | 0 | 0 | 0 | 0 |
| NE | 0 | 0 | 0 | 0 | 0 | 0 | 0 | 0 | 0 | 0 | 0 | 0 | 0 | 0 | 0 | 0 | 0 | 0 |
| NH | 0 | 0 | 0 | 0 | 0 | 0 | 0 | 0 | 0 | 0 | 0 | 0 | 0 | 0 | 0 | 0 | 0 | 0 |
| NJ | 0 | 0 | 0 | 0 | 0 | 0 | 0 | 0 | 0 | 0 | 0 | 0 | 0 | 0 | 0 | 0 | 0 | 0 |
| NM | 0 | 0 | 0 | 0 | 0 | 0 | 0 | 0 | 0 | 0 | 0 | 0 | 0 | 0 | 0 | 0 | 0 | 0 |
| NV | 0 | 0 | 0 | 0 | 0 | 0 | 0 | 0 | 0 | 0 | 0 | 0 | 0 | 0 | 0 | 0 | 0 | 0 |
| NY | 0 | 0 | 0 | 0 | 0 | 0 | 0 | 0 | 0 | 0 | 0 | 0 | 0 | 0 | 0 | 0 | 0 | 0 |
| OH | 0 | 0 | 0 | 0 | 0 | 0 | 0 | 0 | 0 | 0 | 0 | 0 | 0 | 0 | 0 | 0 | 0 | 0 |
| OK | 0 | 0 | 0 | 0 | 0 | 0 | 0 | 0 | 0 | 0 | 0 | 0 | 0 | 0 | 0 | 0 | 0 | 0 |
| OR | 0 | 0 | 0 | 0 | 0 | 0 | 0 | 0 | 0 | 0 | 0 | 0 | 0 | 0 | 0 | 0 | 0 | 0 |
| PA | 0 | 0 | 0 | 0 | 0 | 0 | 0 | 0 | 0 | 0 | 0 | 0 | 0 | 0 | 0 | 0 | 0 | 0 |
| RI | 0 | 0 | 0 | 0 | 0 | 0 | 0 | 0 | 0 | 0 | 0 | 0 | 0 | 0 | 0 | 0 | 0 | 0 |
| SC | 0 | 0 | 0 | 0 | 0 | 0 | 0 | 0 | 0 | 0 | 0 | 0 | 0 | 0 | 0 | 0 | 0 | 0 |
| SD | 0 | 0 | 0 | 0 | 0 | 0 | 0 | 0 | 0 | 0 | 0 | 0 | 0 | 0 | 0 | 0 | 0 | 0 |
| TN | 0 | 0 | 0 | 0 | 0 | 0 | 0 | 0 | 0 | 0 | 0 | 0 | 0 | 0 | 0 | 0 | 0 | 0 |
| TX | 0 | 0 | 0 | 0 | 0 | 0 | 0 | 0 | 0 | 0 | 0 | 0 | 0 | 0 | 0 | 0 | 0 | 0 |
| UT | 0 | 0 | 0 | 0 | 0 | 0 | 0 | 0 | 0 | 0 | 0 | 0 | 0 | 0 | 0 | 0 | 0 | 0 |
| VA | 0 | 0 | 0 | 0 | 0 | 0 | 0 | 0 | 0 | 0 | 0 | 0 | 0 | 0 | 0 | 0 | 0 | 0 |
| VT | 0 | 0 | 0 | 0 | 0 | 0 | 0 | 0 | 0 | 0 | 0 | 0 | 0 | 0 | 0 | 0 | 0 | 0 |
| WA | 1 | 1 | 1 | 1 | 1 | 1 | 1 | 1 | 1 | 1 | 1 | 1 | 1 | 1 | 1 | 1 | 1 | 1 |
| WI | 0 | 0 | 0 | 0 | 0 | 0 | 0 | 0 | 0 | 0 | 0 | 0 | 0 | 0 | 0 | 0 | 0 | 0 |
| WV | 0 | 0 | 0 | 0 | 0 | 0 | 0 | 0 | 0 | 0 | 0 | 0 | 0 | 0 | 0 | 0 | 0 | 0 |
| WY | 0 | 0 | 0 | 0 | 0 | 0 | 0 | 0 | 0 | 0 | 0 | 0 | 0 | 0 | 0 | 0 | 0 | 0 |

| 2/8/2020 | 2/9/2020 | 2/10/2020 | 2/11/2020 | 2/12/2020 | 2/13/2020 | 2/14/2020 | 2/15/2020 | 2/16/2020 | 2/17/2020 | 2/18/2020 | 2/19/2020 | 2/20/2020 | 2/21/2020 | 2/22/2020 | 2/23/2020 | 2/24/2020 | 2/25/2020 | 2/26/2020 | 2/27/2020 |
|---|---|---|---|---|---|---|---|---|---|---|---|---|---|---|---|---|---|---|---|
| 0 | 0 | 0 | 0 | 0 | 0 | 0 | 0 | 0 | 0 | 0 | 0 | 0 | 0 | 0 | 0 | 0 | 0 | 0 | 0 |
| 0 | 0 | 0 | 0 | 0 | 0 | 0 | 0 | 0 | 0 | 0 | 0 | 0 | 0 | 0 | 0 | 0 | 0 | 0 | 0 |
| 0 | 0 | 0 | 0 | 0 | 0 | 0 | 0 | 0 | 0 | 0 | 0 | 0 | 0 | 0 | 0 | 0 | 0 | 0 | 0 |
| 1 | 1 | 1 | 1 | 1 | 1 | 1 | 1 | 1 | 1 | 1 | 1 | 1 | 1 | 1 | 1 | 1 | 1 | 1 | 1 |
| 6 | 6 | 6 | 7 | 7 | 8 | 8 | 8 | 8 | 8 | 8 | 8 | 8 | 10 | 10 | 10 | 10 | 10 | 10 | 11 |
| 0 | 0 | 0 | 0 | 0 | 0 | 0 | 0 | 0 | 0 | 0 | 0 | 0 | 0 | 0 | 0 | 0 | 0 | 0 | 0 |
| 0 | 0 | 0 | 0 | 0 | 0 | 0 | 0 | 0 | 0 | 0 | 0 | 0 | 0 | 0 | 0 | 0 | 0 | 0 | 0 |
| 0 | 0 | 0 | 0 | 0 | 0 | 0 | 0 | 0 | 0 | 0 | 0 | 0 | 0 | 0 | 0 | 0 | 0 | 0 | 0 |
| 0 | 0 | 0 | 0 | 0 | 0 | 0 | 0 | 0 | 0 | 0 | 0 | 0 | 0 | 0 | 0 | 0 | 0 | 0 | 0 |
| 0 | 0 | 0 | 0 | 0 | 0 | 0 | 0 | 0 | 0 | 0 | 0 | 0 | 0 | 0 | 0 | 0 | 0 | 0 | 0 |
| 0 | 0 | 0 | 0 | 0 | 0 | 0 | 0 | 0 | 0 | 0 | 0 | 0 | 0 | 0 | 0 | 0 | 0 | 0 | 0 |
| 0 | 0 | 0 | 0 | 0 | 0 | 0 | 0 | 0 | 0 | 0 | 0 | 0 | 0 | 0 | 0 | 0 | 0 | 0 | 0 |
| 0 | 0 | 0 | 0 | 0 | 0 | 0 | 0 | 0 | 0 | 0 | 0 | 0 | 0 | 0 | 0 | 0 | 0 | 0 | 0 |
| 0 | 0 | 0 | 0 | 0 | 0 | 0 | 0 | 0 | 0 | 0 | 0 | 0 | 0 | 0 | 0 | 0 | 0 | 0 | 0 |
| 2 | 2 | 2 | 2 | 2 | 2 | 2 | 2 | 2 | 2 | 2 | 2 | 2 | 2 | 2 | 2 | 2 | 2 | 2 | 2 |
| 0 | 0 | 0 | 0 | 0 | 0 | 0 | 0 | 0 | 0 | 0 | 0 | 0 | 0 | 0 | 0 | 0 | 0 | 0 | 0 |
| 0 | 0 | 0 | 0 | 0 | 0 | 0 | 0 | 0 | 0 | 0 | 0 | 0 | 0 | 0 | 0 | 0 | 0 | 0 | 0 |
| 0 | 0 | 0 | 0 | 0 | 0 | 0 | 0 | 0 | 0 | 0 | 0 | 0 | 0 | 0 | 0 | 0 | 0 | 0 | 0 |
| 0 | 0 | 0 | 0 | 0 | 0 | 0 | 0 | 0 | 0 | 0 | 0 | 0 | 0 | 0 | 0 | 0 | 0 | 0 | 0 |
| 1 | 1 | 1 | 1 | 1 | 1 | 1 | 1 | 1 | 1 | 1 | 1 | 1 | 1 | 1 | 1 | 1 | 1 | 1 | 1 |
| 0 | 0 | 0 | 0 | 0 | 0 | 0 | 0 | 0 | 0 | 0 | 0 | 0 | 0 | 0 | 0 | 0 | 0 | 0 | 0 |
| 0 | 0 | 0 | 0 | 0 | 0 | 0 | 0 | 0 | 0 | 0 | 0 | 0 | 0 | 0 | 0 | 0 | 0 | 0 | 0 |
| 0 | 0 | 0 | 0 | 0 | 0 | 0 | 0 | 0 | 0 | 0 | 0 | 0 | 0 | 0 | 0 | 0 | 0 | 0 | 0 |
| 0 | 0 | 0 | 0 | 0 | 0 | 0 | 0 | 0 | 0 | 0 | 0 | 0 | 0 | 0 | 0 | 0 | 0 | 0 | 0 |
| 0 | 0 | 0 | 0 | 0 | 0 | 0 | 0 | 0 | 0 | 0 | 0 | 0 | 0 | 0 | 0 | 0 | 0 | 0 | 0 |
| 0 | 0 | 0 | 0 | 0 | 0 | 0 | 0 | 0 | 0 | 0 | 0 | 0 | 0 | 0 | 0 | 0 | 0 | 0 | 0 |
| 0 | 0 | 0 | 0 | 0 | 0 | 0 | 0 | 0 | 0 | 0 | 0 | 0 | 0 | 0 | 0 | 0 | 0 | 0 | 0 |
| 0 | 0 | 0 | 0 | 0 | 0 | 0 | 0 | 0 | 0 | 0 | 0 | 0 | 0 | 0 | 0 | 0 | 0 | 0 | 0 |
| 0 | 0 | 0 | 0 | 0 | 0 | 0 | 0 | 0 | 0 | 0 | 0 | 0 | 0 | 0 | 0 | 0 | 0 | 0 | 0 |
| 0 | 0 | 0 | 0 | 0 | 0 | 0 | 0 | 0 | 0 | 0 | 0 | 0 | 0 | 0 | 0 | 0 | 0 | 0 | 0 |
| 0 | 0 | 0 | 0 | 0 | 0 | 0 | 0 | 0 | 0 | 0 | 0 | 0 | 0 | 0 | 0 | 0 | 0 | 0 | 0 |
| 0 | 0 | 0 | 0 | 0 | 0 | 0 | 0 | 0 | 0 | 0 | 0 | 0 | 0 | 0 | 0 | 0 | 0 | 0 | 0 |
| 0 | 0 | 0 | 0 | 0 | 0 | 0 | 0 | 0 | 0 | 0 | 0 | 0 | 0 | 0 | 0 | 0 | 0 | 0 | 0 |
| 0 | 0 | 0 | 0 | 0 | 0 | 0 | 0 | 0 | 0 | 0 | 0 | 0 | 0 | 0 | 0 | 0 | 0 | 0 | 0 |
| 0 | 0 | 0 | 0 | 0 | 0 | 0 | 0 | 0 | 0 | 0 | 0 | 0 | 0 | 0 | 0 | 0 | 0 | 0 | 0 |
| 0 | 0 | 0 | 0 | 0 | 0 | 0 | 0 | 0 | 0 | 0 | 0 | 0 | 0 | 0 | 0 | 0 | 0 | 0 | 0 |
| 0 | 0 | 0 | 0 | 0 | 0 | 0 | 0 | 0 | 0 | 0 | 0 | 0 | 0 | 0 | 0 | 0 | 0 | 0 | 0 |
| 0 | 0 | 0 | 0 | 0 | 0 | 0 | 0 | 0 | 0 | 0 | 0 | 0 | 0 | 0 | 0 | 0 | 0 | 0 | 0 |
| 0 | 0 | 0 | 0 | 0 | 0 | 0 | 0 | 0 | 0 | 0 | 0 | 0 | 0 | 0 | 0 | 0 | 0 | 0 | 0 |
| 0 | 0 | 0 | 0 | 0 | 0 | 0 | 0 | 0 | 0 | 0 | 0 | 0 | 0 | 0 | 0 | 0 | 0 | 0 | 0 |
| 0 | 0 | 0 | 0 | 0 | 0 | 0 | 0 | 0 | 0 | 0 | 0 | 0 | 0 | 0 | 0 | 0 | 0 | 0 | 0 |
| 0 | 0 | 0 | 0 | 0 | 0 | 0 | 0 | 0 | 0 | 0 | 0 | 0 | 0 | 0 | 0 | 0 | 0 | 0 | 0 |
| 0 | 0 | 0 | 0 | 0 | 0 | 0 | 0 | 0 | 0 | 0 | 0 | 0 | 0 | 0 | 0 | 0 | 0 | 0 | 0 |
| 0 | 0 | 0 | 0 | 0 | 0 | 0 | 0 | 0 | 0 | 0 | 0 | 0 | 0 | 0 | 0 | 0 | 0 | 0 | 0 |
| 0 | 0 | 0 | 0 | 0 | 0 | 0 | 0 | 0 | 0 | 0 | 0 | 0 | 0 | 0 | 0 | 0 | 0 | 0 | 0 |
| 0 | 0 | 0 | 0 | 0 | 0 | 0 | 0 | 0 | 0 | 0 | 0 | 0 | 0 | 0 | 0 | 0 | 0 | 0 | 0 |
| 0 | 0 | 0 | 0 | 0 | 0 | 0 | 0 | 0 | 0 | 0 | 0 | 0 | 0 | 0 | 0 | 0 | 0 | 0 | 0 |
| 1 | 1 | 1 | 1 | 1 | 1 | 1 | 1 | 1 | 1 | 1 | 1 | 1 | 1 | 1 | 1 | 1 | 1 | 1 | 1 |
| 0 | 0 | 0 | 0 | 0 | 0 | 0 | 0 | 0 | 0 | 0 | 0 | 0 | 0 | 0 | 0 | 0 | 0 | 0 | 0 |
| 0 | 0 | 0 | 0 | 0 | 0 | 0 | 0 | 0 | 0 | 0 | 0 | 0 | 0 | 0 | 0 | 0 | 0 | 0 | 0 |
| 0 | 0 | 0 | 0 | 0 | 0 | 0 | 0 | 0 | 0 | 0 | 0 | 0 | 0 | 0 | 0 | 0 | 0 | 0 | 0 |

| 2/28/2020 | 2/29/2020 | 3/1/2020 | 3/2/2020 | 3/3/2020 | 3/4/2020 | 3/5/2020 | 3/6/2020 | 3/7/2020 | 3/8/2020 | 3/9/2020 | 3/10/2020 | 3/11/2020 | 3/12/2020 | 3/13/2020 | 3/14/2020 | 3/15/2020 | 3/16/2020 | 3/17/2020 | 3/18/2020 | 3/19/2020 |
|---|---|---|---|---|---|---|---|---|---|---|---|---|---|---|---|---|---|---|---|---|
| 0 | 0 | 0 | 0 | 0 | 0 | 0 | 0 | 0 | 0 | 0 | 0 | 0 | 0 | 1 | 1 | 1 | 1 | 3 | 6 | 9 |
| 0 | 0 | 0 | 0 | 0 | 0 | 0 | 0 | 0 | 0 | 0 | 0 | 0 | 0 | 1 | 1 | 22 | 29 | 39 | 51 | 78 |
| 0 | 0 | 0 | 0 | 0 | 0 | 0 | 0 | 0 | 0 | 0 | 0 | 1 | 6 | 9 | 12 | 16 | 17 | 24 | 37 | 64 |
| 1 | 1 | 1 | 1 | 2 | 2 | 3 | 3 | 5 | 5 | 6 | 8 | 9 | 9 | 10 | 12 | 13 | 18 | 18 | 27 | 44 |
| 11 | 12 | 12 | 21 | 29 | 36 | 45 | 56 | 56 | 110 | 135 | 152 | 175 | 224 | 264 | 311 | 369 | 447 | 596 | 652 | 982 |
| 0 | 0 | 0 | 0 | 0 | 0 | 1 | 2 | 8 | 8 | 12 | 17 | 34 | 49 | 49 | 49 | 144 | 160 | 183 | 216 | 277 |
| 0 | 0 | 0 | 0 | 0 | 0 | 0 | 0 | 0 | 1 | 1 | 2 | 3 | 7 | 7 | 7 | 26 | 41 | 68 | 96 | 159 |
| 0 | 0 | 0 | 0 | 0 | 0 | 0 | 0 | 1 | 1 | 2 | 2 | 2 | 2 | 3 | 3 | 17 | 22 | 31 | 39 | 71 |
| 0 | 0 | 0 | 0 | 0 | 0 | 0 | 0 | 0 | 0 | 0 | 0 | 1 | 4 | 5 | 8 | 8 | 8 | 16 | 26 | 30 |
| 0 | 0 | 0 | 1 | 2 | 2 | 3 | 3 | 6 | 6 | 19 | 19 | 26 | 31 | 31 | 55 | 136 | 142 | 195 | 299 | 393 |
| 0 | 0 | 0 | 0 | 2 | 2 | 2 | 3 | 6 | 7 | 18 | 23 | 31 | 45 | 45 | 45 | 119 | 143 | 178 | 262 | 397 |
| 0 | 0 | 0 | 0 | 0 | 0 | 0 | 0 | 1 | 1 | 1 | 1 | 1 | 1 | 2 | 2 | 7 | 10 | 10 | 10 | 20 |
| 0 | 0 | 0 | 0 | 0 | 0 | 0 | 0 | 0 | 3 | 8 | 13 | 14 | 16 | 16 | 16 | 22 | 23 | 29 | 38 | 44 |
| 0 | 0 | 0 | 0 | 0 | 0 | 0 | 0 | 0 | 0 | 0 | 0 | 0 | 0 | 0 | 1 | 5 | 5 | 5 | 10 | 23 |
| 2 | 2 | 3 | 4 | 4 | 4 | 5 | 6 | 6 | 7 | 11 | 19 | 27 | 32 | 32 | 32 | 93 | 105 | 160 | 288 | 422 |
| 0 | 0 | 0 | 0 | 0 | 0 | 0 | 1 | 1 | 2 | 3 | 5 | 9 | 12 | 12 | 12 | 19 | 24 | 30 | 39 | 56 |
| 0 | 0 | 0 | 0 | 0 | 0 | 0 | 0 | 0 | 1 | 1 | 1 | 1 | 4 | 6 | 7 | 8 | 11 | 16 | 21 | 35 |
| 0 | 0 | 0 | 0 | 0 | 0 | 0 | 0 | 1 | 1 | 1 | 4 | 4 | 11 | 11 | 15 | 20 | 22 | 26 | 35 | 43 |
| 0 | 0 | 0 | 0 | 0 | 0 | 0 | 0 | 0 | 0 | 1 | 1 | 8 | 14 | 33 | 33 | 101 | 137 | 190 | 261 | 392 |
| 1 | 1 | 1 | 1 | 2 | 2 | 2 | 7 | 9 | 28 | 28 | 91 | 95 | 108 | 109 | 109 | 164 | 197 | 218 | 256 | 328 |
| 0 | 0 | 0 | 0 | 0 | 0 | 3 | 3 | 3 | 3 | 9 | 9 | 12 | 12 | 12 | 12 | 37 | 57 | 85 | 107 | 149 |
| 0 | 0 | 0 | 0 | 0 | 0 | 0 | 0 | 0 | 0 | 0 | 0 | 0 | 1 | 1 | 5 | 14 | 19 | 32 | 44 | 52 |
| 0 | 0 | 0 | 0 | 0 | 0 | 0 | 0 | 0 | 0 | 0 | 2 | 2 | 3 | 13 | 25 | 53 | 54 | 65 | 80 | 334 |
| 0 | 0 | 0 | 0 | 0 | 0 | 0 | 1 | 1 | 2 | 2 | 4 | 5 | 10 | 14 | 14 | 54 | 60 | 77 | 89 | 115 |
| 0 | 0 | 0 | 0 | 0 | 0 | 0 | 0 | 1 | 1 | 1 | 1 | 1 | 1 | 2 | 2 | 5 | 8 | 13 | 24 | 28 |
| 0 | 0 | 0 | 0 | 0 | 0 | 0 | 0 | 0 | 0 | 0 | 0 | 0 | 1 | 1 | 6 | 10 | 12 | 34 | 50 | 80 |
| 0 | 0 | 0 | 0 | 0 | 0 | 0 | 0 | 0 | 0 | 0 | 0 | 1 | 1 | 1 | 1 | 7 | 7 | 11 | 12 | 15 |
| 0 | 0 | 0 | 0 | 1 | 1 | 1 | 2 | 2 | 2 | 7 | 7 | 8 | 15 | 15 | 17 | 33 | 40 | 62 | 97 | 137 |
| 0 | 0 | 0 | 0 | 0 | 0 | 0 | 0 | 0 | 0 | 0 | 0 | 0 | 1 | 1 | 1 | 1 | 1 | 5 | 7 | 19 |
| 0 | 0 | 0 | 0 | 0 | 0 | 0 | 1 | 2 | 2 | 4 | 4 | 5 | 10 | 10 | 10 | 18 | 18 | 21 | 27 | 27 |
| 0 | 0 | 0 | 1 | 2 | 2 | 2 | 2 | 2 | 4 | 4 | 5 | 5 | 6 | 6 | 6 | 13 | 17 | 26 | 39 | 44 |
| 0 | 0 | 0 | 0 | 0 | 0 | 2 | 4 | 6 | 6 | 8 | 15 | 23 | 29 | 30 | 44 | 98 | 178 | 267 | 427 | 742 |
| 0 | 0 | 0 | 0 | 0 | 0 | 0 | 0 | 0 | 0 | 0 | 0 | 3 | 5 | 5 | 7 | 17 | 21 | 23 | 28 | 35 |
| 0 | 0 | 0 | 0 | 0 | 0 | 1 | 1 | 2 | 3 | 4 | 5 | 7 | 11 | 11 | 11 | 16 | 45 | 55 | 63 | 95 |
| 0 | 0 | 0 | 1 | 1 | 2 | 14 | 23 | 30 | 40 | 142 | 173 | 217 | 325 | 362 | 461 | 669 | 831 | 2601 | 4597 | 6834 |
| 0 | 0 | 0 | 0 | 0 | 0 | 0 | 0 | 0 | 0 | 0 | 3 | 3 | 5 | 5 | 5 | 37 | 50 | 67 | 88 | 119 |
| 0 | 0 | 0 | 0 | 0 | 0 | 0 | 0 | 1 | 1 | 1 | 2 | 2 | 3 | 5 | 5 | 9 | 10 | 19 | 31 | 49 |
| 0 | 1 | 1 | 3 | 3 | 3 | 3 | 3 | 7 | 14 | 14 | 15 | 19 | 24 | 24 | 24 | 39 | 47 | 65 | 75 | 88 |
| 0 | 0 | 0 | 0 | 0 | 0 | 0 | 2 | 4 | 6 | 10 | 12 | 16 | 23 | 29 | 47 | 63 | 76 | 96 | 133 | 185 |
| 0 | 0 | 1 | 2 | 2 | 2 | 2 | 2 | 3 | 3 | 3 | 5 | 5 | 5 | 5 | 9 | 21 | 23 | 23 | 33 | 44 |
| 0 | 0 | 0 | 0 | 0 | 0 | 0 | 0 | 2 | 6 | 7 | 9 | 10 | 12 | 12 | 12 | 28 | 33 | 47 | 60 | 81 |
| 0 | 0 | 0 | 0 | 0 | 0 | 0 | 0 | 0 | 0 | 0 | 5 | 8 | 8 | 9 | 9 | 9 | 10 | 11 | 11 | 14 |
| 0 | 0 | 0 | 0 | 0 | 0 | 1 | 1 | 1 | 1 | 1 | 7 | 9 | 18 | 26 | 26 | 39 | 52 | 73 | 98 | 154 |
| 0 | 0 | 0 | 0 | 0 | 0 | 3 | 5 | 5 | 6 | 13 | 15 | 23 | 24 | 36 | 43 | 51 | 57 | 64 | 83 | 143 |
| 0 | 0 | 0 | 0 | 0 | 0 | 0 | 0 | 0 | 1 | 1 | 2 | 3 | 4 | 6 | 6 | 21 | 29 | 29 | 40 | 68 |
| 0 | 0 | 0 | 0 | 0 | 0 | 0 | 0 | 0 | 2 | 5 | 8 | 15 | 29 | 29 | 29 | 51 | 67 | 68 | 94 | 114 |
| 0 | 0 | 0 | 0 | 0 | 0 | 0 | 0 | 0 | 1 | 1 | 1 | 1 | 2 | 2 | 3 | 4 | 8 | 10 | 12 | 16 |
| 1 | 7 | 11 | 18 | 27 | 39 | 70 | 79 | 102 | 136 | 162 | 267 | 366 | 457 | 568 | 642 | 708 | 769 | 930 | 1,187 | 1,376 |
| 0 | 0 | 0 | 0 | 1 | 1 | 1 | 1 | 1 | 1 | 1 | 3 | 5 | 8 | 9 | 19 | 27 | 53 | 88 | 106 | 155 |
| 0 | 0 | 0 | 0 | 0 | 0 | 0 | 0 | 0 | 0 | 0 | 0 | 0 | 0 | 0 | 0 | 0 | 0 | 1 | 2 | 5 |
| 0 | 0 | 0 | 0 | 0 | 0 | 0 | 0 | 0 | 0 | 0 | 0 | 0 | 1 | 1 | 2 | 3 | 3 | 11 | 16 | 18 |

| 3/20/2020 | 3/21/2020 | 3/22/2020 | 3/23/2020 | 3/24/2020 | 3/25/2020 | 3/26/2020 | 3/27/2020 | 3/28/2020 | 3/29/2020 | 3/30/2020 | 3/31/2020 | 4/1/2020 | 4/2/2020 | 4/3/2020 | 4/4/2020 | 4/5/2020 | 4/6/2020 | 4/7/2020 | 4/8/2020 | 4/9/2020 |
|---|---|---|---|---|---|---|---|---|---|---|---|---|---|---|---|---|---|---|---|---|
| 12 | 14 | 22 | 36 | 42 | 59 | 69 | 85 | 102 | 114 | 119 | 133 | 143 | 147 | 157 | 171 | 185 | 191 | 213 | 226 | 235 |
| 106 | 131 | 157 | 196 | 242 | 386 | 531 | 639 | 720 | 830 | 947 | 999 | 1,106 | 1,270 | 1,535 | 1,633 | 1,841 | 2,006 | 2,197 | 2,499 | 2,838 |
| 100 | 118 | 164 | 197 | 230 | 306 | 349 | 386 | 409 | 443 | 504 | 560 | 614 | 679 | 738 | 743 | 853 | 915 | 993 | 1,071 | 1,127 |
| 63 | 104 | 152 | 253 | 326 | 401 | 508 | 665 | 773 | 919 | 1,157 | 1,289 | 1,413 | 1,598 | 1,769 | 2,019 | 2,269 | 2,456 | 2,575 | 2,726 | 3,018 |
| 1,304 | 1,534 | 1,709 | 1,931 | 2,511 | 2,982 | 3,777 | 4,730 | 5,259 | 5,739 | 6,909 | 8,131 | 8,155 | 9,191 | 12,024 | 12,026 | 13,438 | 14,336 | 15,865 | 16,957 | 18,309 |
| 363 | 475 | 591 | 720 | 921 | 1,086 | 1,430 | 1,734 | 2,061 | 2,307 | 2,627 | 2,966 | 3,342 | 3,728 | 4,173 | 4,565 | 4,950 | 5,172 | 5,429 | 5,655 | 6,202 |
| 194 | 223 | 327 | 415 | 618 | 875 | 1,012 | 1,291 | 1,524 | 1,993 | 2,571 | 3,128 | 3,557 | 3,824 | 4,915 | 5,276 | 5,675 | 6,906 | 7,781 | 8,781 | 9,784 |
| 77 | 98 | 116 | 137 | 183 | 231 | 267 | 304 | 342 | 401 | 495 | 495 | 586 | 653 | 757 | 902 | 998 | 1,097 | 1,211 | 1,440 | 1,523 |
| 39 | 45 | 56 | 87 | 104 | 119 | 143 | 165 | 214 | 232 | 264 | 319 | 368 | 393 | 450 | 593 | 673 | 783 | 928 | 1,116 | 1,209 |
| 510 | 706 | 937 | 1,141 | 1,373 | 1,861 | 2,352 | 3,054 | 3,877 | 4,768 | 5,489 | 6,490 | 7,495 | 8,694 | 9,925 | 11,173 | 11,961 | 13,214 | 14,302 | 15,234 | 16,323 |
| 485 | 555 | 757 | 1,015 | 1,119 | 1,441 | 1,714 | 2,198 | 2,446 | 2,703 | 3,612 | 4,585 | 4,748 | 5,486 | 5,967 | 6,383 | 6,752 | 8,634 | 9,713 | 10,446 | 11,318 |
| 37 | 48 | 48 | 66 | 70 | 76 | 86 | 98 | 126 | 141 | 163 | 185 | 205 | 225 | 270 | 310 | 324 | 332 | 362 | 377 | 386 |
| 45 | 68 | 90 | 105 | 124 | 145 | 179 | 235 | 298 | 336 | 424 | 497 | 549 | 614 | 699 | 786 | 868 | 946 | 1,048 | 1,145 | 1,270 |
| 31 | 42 | 47 | 51 | 71 | 123 | 189 | 230 | 261 | 310 | 415 | 525 | 669 | 891 | 1,013 | 1,077 | 1,101 | 1,170 | 1,210 | 1,232 | 1,353 |
| 585 | 753 | 1,049 | 1,285 | 1,535 | 1,865 | 2,538 | 3,026 | 3,491 | 4,596 | 5,057 | 5,994 | 6,980 | 7,695 | 8,904 | 10,357 | 11,256 | 12,262 | 13,549 | 15,078 | 16,422 |
| 79 | 126 | 201 | 259 | 365 | 477 | 645 | 981 | 1,232 | 1,514 | 1,786 | 2,159 | 2,565 | 3,039 | 3,457 | 3,953 | 4,411 | 4,944 | 5,507 | 5,943 | 6,351 |
| 44 | 55 | 64 | 82 | 98 | 98 | 168 | 202 | 261 | 319 | 368 | 428 | 482 | 552 | 620 | 698 | 813 | 845 | 900 | 1,076 | 1,106 |
| 48 | 87 | 103 | 124 | 163 | 198 | 248 | 302 | 394 | 439 | 480 | 591 | 680 | 770 | 831 | 917 | 955 | 1,008 | 1,149 | 1,346 | 1,452 |
| 537 | 763 | 1,035 | 1,172 | 1,388 | 1,946 | 2,305 | 2,746 | 3,315 | 3,540 | 4,025 | 5,237 | 6,424 | 9,150 | 10,297 | 12,496 | 13,010 | 14,867 | 16,284 | 17,030 | 18,283 |
| 413 | 525 | 646 | 777 | 1,159 | 1,838 | 2,417 | 3,240 | 4,257 | 4,955 | 5,752 | 6,620 | 7,738 | 8,966 | 10,402 | 11,736 | 12,500 | 13,837 | 15,202 | 16,567 | 18,941 |
| 149 | 190 | 288 | 288 | 349 | 580 | 774 | 774 | 992 | 1,239 | 1,660 | 1,660 | 1,985 | 2,331 | 2,758 | 3,125 | 4,045 | 4,371 | 5,529 | 6,185 | 6,968 |
| 56 | 70 | 93 | 109 | 118 | 147 | 155 | 168 | 211 | 253 | 303 | 303 | 344 | 376 | 432 | 456 | 470 | 499 | 519 | 537 | 560 |
| 549 | 787 | 1,035 | 1,328 | 1,791 | 2,294 | 2,856 | 3,657 | 4,650 | 5,486 | 6,498 | 7,615 | 9,334 | 10,791 | 12,670 | 14,225 | 15,718 | 17,221 | 18,970 | 20,346 | 21,504 |
| 115 | 137 | 236 | 262 | 287 | 346 | 399 | 399 | 441 | 576 | 576 | 689 | 742 | 789 | 789 | 865 | 986 | 1,069 | 1,154 | 1,242 | 1,336 |
| 73 | 90 | 106 | 183 | 255 | 356 | 502 | 670 | 838 | 903 | 1,031 | 1,327 | 1,581 | 1,834 | 2,113 | 2,291 | 2,367 | 2,722 | 3,037 | 3,327 | 3,539 |
| 80 | 140 | 249 | 320 | 377 | 485 | 579 | 579 | 663 | 847 | 937 | 1,073 | 1,177 | 1,358 | 1,358 | 1,455 | 1,738 | 1,915 | 2,003 | 2,260 | 2,469 |
| 21 | 27 | 34 | 37 | 48 | 65 | 90 | 121 | 147 | 173 | 184 | 203 | 228 | 244 | 262 | 281 | 300 | 320 | 332 | 352 | 365 |
| 164 | 227 | 255 | 398 | 504 | 636 | 764 | 764 | 935 | 1,307 | 1,498 | 1,584 | 1,857 | 2,093 | 2,093 | 2,402 | 2,870 | 2,870 | 3,221 | 3,651 | 3,651 |
| 26 | 28 | 30 | 32 | 37 | 45 | 58 | 68 | 94 | 98 | 109 | 126 | 147 | 159 | 173 | 186 | 207 | 225 | 237 | 251 | 269 |
| 38 | 42 | 50 | 52 | 53 | 64 | 73 | 89 | 108 | 120 | 145 | 177 | 214 | 255 | 285 | 323 | 367 | 412 | 478 | 523 | 577 |
| 55 | 65 | 78 | 101 | 108 | 137 | 158 | 187 | 214 | 258 | 314 | 367 | 415 | 479 | 540 | 621 | 669 | 715 | 747 | 788 | 819 |
| 890 | 1,327 | 1,914 | 2,844 | 3,675 | 4,402 | 6,876 | 8,825 | 11,124 | 13,386 | 16,636 | 18,696 | 22,255 | 25,590 | 29,895 | 34,124 | 37,505 | 41,090 | 44,416 | 47,437 | 51,027 |
| 43 | 57 | 65 | 83 | 100 | 112 | 136 | 191 | 208 | 237 | 281 | 315 | 363 | 403 | 495 | 543 | 624 | 686 | 794 | 865 | 989 |
| 109 | 154 | 190 | 245 | 278 | 321 | 535 | 621 | 738 | 920 | 1,044 | 1,113 | 1,279 | 1,458 | 1,514 | 1,742 | 1,836 | 1,953 | 2,087 | 2,318 | 2,456 |
| 7845 | 10356 | 15168 | 21689 | 26358 | 32966 | 38977 | 45934 | 53308 | 59219 | 67131 | 74427 | 84735 | 90279 | 101993 | 111248 | 119435 | 141100 | 138773 | 149316 | 157073 |
| 169 | 247 | 351 | 442 | 564 | 704 | 867 | 1,137 | 1,406 | 1,653 | 1,933 | 2,199 | 2,547 | 2,902 | 3,312 | 3,739 | 4,043 | 4,450 | 4,782 | 5,148 | 5,512 |
| 49 | 53 | 67 | 81 | 106 | 223 | 248 | 322 | 377 | 481 | 481 | 565 | 719 | 879 | 988 | 1,159 | 1,250 | 1,327 | 1,472 | 1,524 | 1,684 |
| 114 | 137 | 161 | 191 | 209 | 266 | 316 | 414 | 479 | 548 | 606 | 690 | 736 | 826 | 899 | 999 | 1,068 | 1,132 | 1,181 | 1,239 | 1,321 |
| 269 | 371 | 479 | 644 | 851 | 1,128 | 1,688 | 2,219 | 2,752 | 3,394 | 4,087 | 4,843 | 5,805 | 7,016 | 8,421 | 10,017 | 11,510 | 12,980 | 14,559 | 16,239 | 18,228 |
| 54 | 66 | 97 | 106 | 124 | 132 | 188 | 203 | 239 | 294 | 479 | 520 | 566 | 681 | 711 | 806 | 922 | 1,170 | 1,414 | 1,450 | 1,939 |
| 125 | 173 | 195 | 298 | 342 | 424 | 456 | 539 | 660 | 774 | 925 | 1,083 | 1,293 | 1,554 | 1,700 | 1,917 | 2,049 | 2,232 | 2,417 | 2,552 | 2,792 |
| 14 | 14 | 21 | 28 | 30 | 41 | 46 | 58 | 68 | 90 | 101 | 108 | 129 | 165 | 187 | 212 | 240 | 288 | 320 | 393 | 447 |
| 228 | 371 | 505 | 615 | 667 | 784 | 957 | 1,203 | 1,373 | 1,537 | 1,834 | 2,239 | 2,240 | 2,845 | 2,880 | 3,103 | 3,633 | 3,803 | 4,139 | 4,362 | 4,635 |
| 401 | 507 | 507 | 507 | 715 | 974 | 1,396 | 1,731 | 2,052 | 2,552 | 2,877 | 3,266 | 3,997 | 4,669 | 5,330 | 6,110 | 6,812 | 7,276 | 8,262 | 9,353 | 10,230 |
| 102 | 125 | 169 | 280 | 298 | 346 | 402 | 466 | 588 | 789 | 806 | 934 | 1,012 | 1,165 | 1,246 | 1,428 | 1,605 | 1,716 | 1,804 | 1,916 | 2,061 |
| 114 | 152 | 254 | 290 | 391 | 460 | 604 | 604 | 739 | 1,020 | 1,020 | 1,484 | 1,706 | 2,012 | 2,012 | 2,407 | 2,878 | 3,333 | 3,645 | 4,042 | 4,509 |
| 29 | 49 | 52 | 75 | 95 | 123 | 156 | 184 | 211 | 235 | 256 | 293 | 321 | 338 | 389 | 461 | 512 | 543 | 575 | 605 | 628 |
| 1,524 | 1,793 | 1,996 | 2,221 | 2,469 | 2,580 | 3,207 | 3,723 | 4,310 | 4,506 | 4,896 | 4,896 | 5,319 | 5,683 | 6,966 | 7,591 | 6,973 | 7,318 | 8,682 | 9,097 | 9,608 |
| 206 | 281 | 381 | 416 | 457 | 585 | 707 | 842 | 989 | 989 | 1,221 | 1,351 | 1,550 | 1,730 | 1,916 | 2,112 | 2,267 | 2,440 | 2,578 | 2,756 | 2,885 |
| 8 | 12 | 16 | 20 | 39 | 51 | 76 | 96 | 113 | 124 | 145 | 162 | 191 | 217 | 237 | 282 | 324 | 345 | 412 | 412 | 523 |
| 22 | 24 | 24 | 26 | 30 | 44 | 55 | 73 | 84 | 87 | 95 | 120 | 137 | 150 | 166 | 187 | 200 | 212 | 221 | 230 | 239 |

| 4/10/2020 | 4/11/2020 | 4/12/2020 | 4/13/2020 | 4/14/2020 | 4/15/2020 | 4/16/2020 | 4/17/2020 | 4/18/2020 | 4/19/2020 | 4/20/2020 | 4/21/2020 | 4/22/2020 | 4/23/2020 | 4/24/2020 | 4/25/2020 | 4/26/2020 | 4/27/2020 | 4/28/2020 | 4/29/2020 |
|---|---|---|---|---|---|---|---|---|---|---|---|---|---|---|---|---|---|---|---|
| 246 | 257 | 272 | 277 | 285 | 293 | 300 | 309 | 314 | 319 | 321 | 329 | 335 | 337 | 339 | 339 | 341 | 345 | 351 | 355 |
| 3,008 | 3,262 | 3,583 | 3,803 | 3,953 | 4,241 | 4,404 | 4,572 | 4,723 | 4,946 | 5,424 | 5,668 | 5,831 | 6,071 | 6,026 | 6,213 | 6,627 | 6,539 | 6,947 | 7,105 |
| 1,202 | 1,234 | 1,307 | 1,410 | 1,498 | 1,583 | 1,635 | 1,702 | 1,777 | 1,781 | 1,964 | 2,262 | 2,281 | 2,465 | 2,810 | 2,909 | 2,941 | 3,055 | 3,121 | 3,192 |
| 3,112 | 3,393 | 3,539 | 3,702 | 3,806 | 3,962 | 4,234 | 4,507 | 4,719 | 4,929 | 5,064 | 5,251 | 5,459 | 5,769 | 6,045 | 6,280 | 6,526 | 6,716 | 6,948 | 7,202 |
| 19,472 | 20,615 | 21,794 | 22,348 | 23,338 | 24,424 | 26,182 | 27,528 | 28,963 | 28,963 | 30,978 | 33,261 | 35,396 | 37,369 | 39,254 | 41,137 | 42,164 | 43,464 | 45,031 | 46,500 |
| 6,365 | 6,893 | 7,303 | 7,691 | 7,941 | 8,280 | 8,582 | 9,047 | 9,433 | 9,634 | 10,019 | 10,368 | 10,825 | 11,182 | 12,184 | 12,868 | 13,350 | 13,798 | 14,238 | 14,675 |
| 10,131 | 11,510 | 12,035 | 13,381 | 13,989 | 14,755 | 15,884 | 16,809 | 17,550 | 17,962 | 19,815 | 20,360 | 22,469 | 23,100 | 23,921 | 24,582 | 25,269 | 25,997 | 26,312 | 26,767 |
| 1,660 | 1,778 | 1,875 | 1,955 | 2,058 | 2,197 | 2,350 | 2,476 | 2,666 | 2,793 | 2,927 | 3,098 | 3,206 | 3,361 | 3,528 | 3,699 | 3,841 | 3,892 | 3,994 | 4,106 |
| 1,326 | 1,479 | 1,625 | 1,761 | 1,926 | 2,014 | 2,075 | 2,323 | 2,538 | 2,745 | 2,745 | 3,200 | 3,308 | 3,442 | 3,442 | 3,576 | 4,162 | 4,575 | 4,655 | 4,734 |
| 17,448 | 18,445 | 19,337 | 20,394 | 20,984 | 21,865 | 22,674 | 24,066 | 24,797 | 25,598 | 26,329 | 27,127 | 27,791 | 28,843 | 29,707 | 29,996 | 30,680 | 31,290 | 31,986 | 32,318 |
| 11,318 | 12,261 | 13,012 | 13,621 | 14,766 | 15,409 | 16,451 | 16,658 | 17,014 | 18,781 | 19,630 | 20,607 | 20,769 | 21,681 | 22,183 | 22,225 | 23,410 | 23,801 | 24,609 | 25,324 |
| 409 | 452 | 465 | 504 | 517 | 517 | 541 | 553 | 574 | 580 | 584 | 547 | 537 | 540 | 601 | 604 | 550 | 607 | 553 | 557 |
| 1,388 | 1,510 | 1,587 | 1,710 | 1,899 | 1,995 | 2,141 | 2,332 | 2,513 | 2,902 | 3,159 | 3,641 | 3,748 | 3,924 | 4,445 | 5,092 | 5,475 | 5,868 | 6,376 | 6,843 |
| 1,396 | 1,407 | 1,458 | 1,486 | 1,486 | 1,587 | 1,609 | 1,655 | 1,668 | 1,672 | 1,736 | 1,766 | 1,802 | 1,836 | 1,870 | 1,887 | 1,897 | 1,917 | 1,952 | 1,984 |
| 17,887 | 19,180 | 20,852 | 22,025 | 23,247 | 24,593 | 25,733 | 27,575 | 29,160 | 30,357 | 31,508 | 33,059 | 35,108 | 36,934 | 39,658 | 41,777 | 43,903 | 45,883 | 48,102 | 50,355 |
| 6,907 | 7,435 | 7,928 | 8,236 | 8,527 | 8,955 | 9,542 | 10,154 | 10,641 | 11,210 | 11,686 | 12,097 | 12,438 | 13,039 | 13,680 | 14,395 | 15,012 | 15,961 | 16,588 | 17,182 |
| 1,241 | 1,279 | 1,377 | 1,376 | 1,477 | 1,494 | 1,588 | 1,760 | 1,811 | 1,849 | 1,986 | 2,025 | 2,211 | 2,482 | 2,777 | 3,056 | 3,174 | 3,328 | 3,491 | 3,738 |
| 1,693 | 1,840 | 1,963 | 2,048 | 2,210 | 2,291 | 2,429 | 2,522 | 2,707 | 2,960 | 3,050 | 3,192 | 3,373 | 3,481 | 3,779 | 3,905 | 4,074 | 4,146 | 4,375 | 4,539 |
| 19,253 | 20,014 | 20,595 | 21,016 | 21,518 | 21,951 | 22,532 | 23,118 | 23,580 | 23,928 | 24,523 | 24,854 | 25,317 | 25,798 | 26,140 | 26,512 | 26,832 | 27,111 | 27,329 | 27,703 |
| 20,845 | 23,918 | 25,475 | 26,867 | 28,163 | 29,918 | 32,181 | 34,402 | 36,372 | 38,077 | 39,643 | 41,199 | 42,944 | 46,023 | 50,969 | 53,348 | 54,938 | 56,462 | 58,302 | 60,265 |
| 6,968 | 7,694 | 8,936 | 9,472 | 10,032 | 10,784 | 11,572 | 11,572 | 12,308 | 13,684 | 14,193 | 14,775 | 15,737 | 16,616 | 16,616 | 17,766 | 19,487 | 20,113 | 20,849 | 21,742 |
| 583 | 616 | 633 | 698 | 734 | 770 | 796 | 827 | 847 | 867 | 875 | 888 | 907 | 937 | 965 | 990 | 1,023 | 1,040 | 1,056 | 1,095 |
| 22,562 | 23,853 | 24,638 | 25,635 | 27,001 | 28,059 | 29,263 | 29,952 | 30,717 | 31,424 | 32,000 | 32,967 | 33,966 | 35,291 | 36,627 | 37,184 | 37,778 | 38,210 | 39,262 | 40,399 |
| 1,336 | 1,514 | 1,621 | 1,695 | 1,809 | 1,862 | 2,013 | 2,174 | 2,213 | 2,470 | 2,567 | 2,721 | 2,721 | 3,185 | 3,185 | 3,446 | 3,602 | 4,181 | 4,645 | 5,136 |
| 3,799 | 4,024 | 4,160 | 4,388 | 4,686 | 4,895 | 5,111 | 5,283 | 5,517 | 5,667 | 5,807 | 5,941 | 6,137 | 6,321 | 6,625 | 6,826 | 6,997 | 7,171 | 7,303 | 7,425 |
| 2,469 | 2,642 | 2,942 | 3,087 | 3,360 | 3,624 | 3,793 | 3,793 | 3,974 | 4,512 | 4,716 | 4,716 | 5,153 | 5,434 | 5,434 | 5,718 | 6,094 | 6,342 | 6,569 | 6,569 |
| 365 | 377 | 394 | 398 | 404 | 415 | 422 | 422 | 426 | 433 | 437 | 439 | 442 | 444 | 444 | 445 | 449 | 451 | 451 | 451 |
| 3,908 | 4,312 | 4,520 | 4,816 | 5,024 | 5,123 | 5,465 | 5,859 | 6,140 | 6,493 | 6,764 | 6,951 | 7,220 | 7,608 | 8,052 | 8,623 | 8,830 | 9,142 | 9,568 | 9,948 |
| 278 | 293 | 308 | 331 | 341 | 365 | 393 | 439 | 528 | 585 | 627 | 644 | 679 | 709 | 748 | 803 | 867 | 942 | 991 | 1,033 |
| 648 | 704 | 814 | 871 | 901 | 952 | 1,066 | 1,138 | 1,287 | 1,474 | 1,648 | 1,722 | 1,813 | 2,124 | 2,421 | 2,732 | 3,028 | 3,358 | 3,374 | 3,784 |
| 885 | 929 | 985 | 1,020 | 1,091 | 1,139 | 1,211 | 1,287 | 1,342 | 1,392 | 1,447 | 1,491 | 1,588 | 1,670 | 1,720 | 1,787 | 1,864 | 1,938 | 2,010 | 2,054 |
| 54,588 | 58,151 | 61,850 | 64,584 | 68,824 | 71,030 | 75,317 | 78,467 | 81,436 | 85,301 | 88,806 | 92,387 | 95,865 | 99,989 | 102,196 | 105,523 | 109,038 | 111,188 | 113,856 | 116,264 |
| 1,091 | 1,174 | 1,245 | 1,345 | 1,407 | 1,484 | 1,597 | 1,711 | 1,798 | 1,845 | 1,971 | 2,072 | 2,210 | 2,379 | 2,521 | 2,660 | 2,726 | 2,823 | 2,974 | 3,213 |
| 2,584 | 2,700 | 2,836 | 2,971 | 3,088 | 3,211 | 3,321 | 3,524 | 3,626 | 3,728 | 3,830 | 3,937 | 4,081 | 4,208 | 4,398 | 4,539 | 4,602 | 4,690 | 4,812 | 4,898 |
| 170884 | 180458 | 188694 | 195081 | 201834 | 211550 | 216703 | 224438 | 233570 | 238138 | 245580 | 250800 | 253219 | 263744 | 267256 | 277606 | 282991 | 287607 | 290481 | 294715 |
| 5,878 | 6,250 | 6,518 | 6,975 | 7,280 | 7,791 | 8,414 | 9,107 | 10,222 | 11,602 | 12,919 | 13,725 | 14,117 | 14,694 | 15,169 | 15,587 | 15,963 | 16,325 | 16,769 | 17,303 |
| 1,794 | 1,868 | 1,970 | 2,069 | 2,209 | 2,263 | 2,359 | 2,465 | 2,570 | 2,599 | 2,680 | 2,807 | 2,894 | 3,017 | 3,121 | 3,193 | 3,253 | 3,280 | 3,410 | 3,473 |
| 1,371 | 1,447 | 1,527 | 1,584 | 1,633 | 1,663 | 1,736 | 1,785 | 1,844 | 1,910 | 1,956 | 2,002 | 2,059 | 2,127 | 2,177 | 2,253 | 2,311 | 2,354 | 2,385 | 2,446 |
| 19,980 | 21,656 | 22,833 | 24,199 | 25,345 | 26,490 | 27,735 | 29,442 | 31,070 | 32,284 | 33,232 | 34,528 | 35,684 | 37,053 | 38,652 | 40,049 | 41,165 | 42,050 | 43,264 | 44,366 |
| 2,015 | 2,349 | 2,665 | 2,976 | 3,369 | 3,529 | 3,969 | 4,177 | 4,491 | 4,706 | 5,090 | 5,716 | 6,012 | 6,256 | 6,699 | 7,129 | 7,439 | 7,708 | 7,926 | 8,247 |
| 3,065 | 3,207 | 3,319 | 3,439 | 3,553 | 3,656 | 3,931 | 4,086 | 4,246 | 4,377 | 4,439 | 4,608 | 4,761 | 4,917 | 5,070 | 5,253 | 5,490 | 5,613 | 5,735 | 5,881 |
| 536 | 626 | 730 | 868 | 1,168 | 1,168 | 1,311 | 1,411 | 1,542 | 1,635 | 1,685 | 1,754 | 1,858 | 1,956 | 2,040 | 2,147 | 2,212 | 2,245 | 2,313 | 2,373 |
| 4,863 | 4,863 | 5,308 | 5,610 | 5,823 | 5,836 | 6,262 | 6,330 | 6,763 | 6,796 | 7,239 | 7,394 | 7,572 | 8,281 | 8,742 | 8,882 | 9,698 | 9,952 | 10,087 | 10,132 |
| 11,671 | 12,561 | 13,484 | 13,906 | 14,624 | 15,492 | 16,455 | 17,371 | 18,260 | 18,923 | 19,458 | 20,196 | 21,069 | 21,944 | 22,806 | 23,773 | 24,631 | 25,297 | 26,171 | 27,054 |
| 2,102 | 2,206 | 2,338 | 2,392 | 2,453 | 2,637 | 2,760 | 2,805 | 2,946 | 3,151 | 3,259 | 3,369 | 3,540 | 3,722 | 3,782 | 3,948 | 4,214 | 4,307 | 4,423 | 4,605 |
| 4,509 | 5,077 | 5,747 | 6,171 | 6,500 | 6,889 | 7,491 | 8,053 | 8,053 | 8,990 | 9,630 | 9,630 | 10,998 | 11,594 | 12,366 | 12,366 | 13,535 | 14,339 | 14,961 | 15,846 |
| 679 | 711 | 727 | 748 | 750 | 759 | 768 | 779 | 803 | 812 | 816 | 818 | 823 | 825 | 827 | 843 | 851 | 855 | 862 | 862 |
| 9,887 | 10,224 | 10,411 | 10,538 | 10,694 | 10,783 | 11,152 | 11,445 | 11,802 | 11,790 | 12,085 | 12,282 | 12,494 | 12,753 | 12,977 | 13,319 | 13,521 | 13,686 | 13,842 | 14,070 |
| 3,068 | 3,221 | 3,341 | 3,428 | 3,555 | 3,721 | 3,875 | 4,045 | 4,199 | 4,346 | 4,499 | 4,620 | 4,845 | 5,052 | 5,356 | 5,687 | 5,911 | 6,081 | 6,289 | 6,520 |
| 574 | 591 | 611 | 626 | 694 | 718 | 739 | 775 | 825 | 863 | 902 | 914 | 963 | 981 | 1,010 | 1,025 | 1,053 | 1,077 | 1,095 | 1,109 |
| 253 | 261 | 270 | 373 | 494 | 393 | 401 | 412 | 423 | 426 | 428 | 441 | 447 | 453 | 473 | 491 | 502 | 520 | 536 | 544 |

| 4/30/2020 | 5/1/2020 | 5/2/2020 | 5/3/2020 | 5/4/2020 | 5/5/2020 | 5/6/2020 | 5/7/2020 | 5/8/2020 | 5/9/2020 | 5/10/2020 | 5/11/2020 | 5/12/2020 | 5/13/2020 | 5/14/2020 | 5/15/2020 | 5/16/2020 | 5/17/2020 | 5/18/2020 | 5/19/2020 | 5/20/2020 |
|---|---|---|---|---|---|---|---|---|---|---|---|---|---|---|---|---|---|---|---|---|
| 355 | 364 | 365 | 368 | 370 | 371 | 372 | 374 | 377 | 378 | 379 | 381 | 383 | 383 | 387 | 388 | 392 | 396 | 399 | 399 | 402 |
| 7,306 | 7,294 | 7,611 | 7,888 | 8,312 | 8,679 | 8,691 | 9,243 | 9,639 | 9,923 | 10,145 | 10,413 | 10,665 | 10,936 | 11,338 | 11,642 | 11,981 | 11,771 | 12,587 | 12,376 | 13,300 |
| 3,255 | 3,310 | 3,372 | 3,431 | 3,469 | 3,496 | 3,611 | 3,665 | 3,747 | 3,984 | 3,984 | 4,043 | 4,164 | 4,236 | 4,348 | 4,463 | 4,578 | 4,759 | 4,813 | 4,923 | 5,003 |
| 7,648 | 7,962 | 8,364 | 8,640 | 8,919 | 9,305 | 9,707 | 9,945 | 10,526 | 10,960 | 11,119 | 11,380 | 11,736 | 12,176 | 12,674 | 13,169 | 13,631 | 13,937 | 14,170 | 14,566 | 14,897 |
| 48,917 | 50,442 | 52,197 | 53,616 | 54,937 | 56,212 | 58,815 | 60,614 | 62,512 | 64,561 | 66,680 | 67,939 | 69,382 | 71,141 | 73,164 | 74,936 | 76,793 | 78,839 | 80,430 | 81,795 | 84,057 |
| 15,182 | 15,668 | 16,120 | 16,534 | 16,878 | 17,317 | 17,738 | 18,318 | 18,793 | 19,316 | 19,632 | 19,735 | 20,091 | 20,401 | 20,762 | 21,131 | 21,511 | 21,797 | 22,095 | 22,399 | 22,752 |
| 27,700 | 28,764 | 29,287 | 29,312 | 29,973 | 30,621 | 30,995 | 31,784 | 32,411 | 32,984 | 33,554 | 33,765 | 34,333 | 34,855 | 35,464 | 36,085 | 36,703 | 37,419 | 38,116 | 38,430 | 39,017 |
| 4,323 | 4,658 | 4,797 | 5,016 | 5,170 | 5,322 | 5,461 | 5,654 | 5,899 | 6,102 | 6,272 | 6,389 | 6,485 | 6,584 | 6,736 | 6,871 | 7,042 | 7,123 | 7,270 | 7,434 | 7,551 |
| 4,734 | 4,918 | 5,038 | 5,208 | 5,288 | 5,371 | 5,778 | 6,111 | 6,111 | 6,447 | 6,565 | 6,741 | 6,952 | 7,223 | 7,373 | 7,373 | 7,547 | 7,869 | 8,037 | 8,194 | 8,386 |
| 33,690 | 34,728 | 35,463 | 35,969 | 36,897 | 37,439 | 38,002 | 38,828 | 39,199 | 40,001 | 39,888 | 40,982 | 41,923 | 42,402 | 43,210 | 42,940 | 44,811 | 45,588 | 46,442 | 46,944 | 47,471 |
| 26,095 | 27,733 | 28,338 | 28,621 | 29,196 | 29,602 | 30,750 | 31,383 | 32,609 | 32,590 | 32,980 | 33,995 | 34,731 | 35,335 | 35,858 | 36,680 | 37,147 | 37,642 | 38,081 | 38,753 | 39,702 |
| 562 | 619 | 620 | 557 | 558 | 570 | 626 | 576 | 576 | 579 | 580 | 582 | 583 | 587 | 586 | 587 | 588 | 589 | 589 | 591 | 593 |
| 7,145 | 7,884 | 8,641 | 9,169 | 9,703 | 10,111 | 10,404 | 11,059 | 11,457 | 11,671 | 11,959 | 12,373 | 12,912 | 13,289 | 13,675 | 14,049 | 14,328 | 14,651 | 15,082 | 15,349 | 15,595 |
| 2,015 | 2,035 | 2,061 | 2,061 | 2,106 | 2,127 | 2,158 | 2,178 | 2,205 | 2,230 | 2,230 | 2,260 | 2,293 | 2,324 | 2,351 | 2,389 | 2,419 | 2,419 | 2,455 | 2,476 | 2,506 |
| 52,918 | 56,055 | 58,505 | 61,449 | 63,840 | 65,962 | 68,232 | 70,873 | 73,760 | 76,085 | 77,741 | 79,007 | 83,021 | 84,698 | 87,937 | 90,369 | 92,457 | 94,191 | 96,485 | 98,030 | 100,418 |
| 17,835 | 18,630 | 19,295 | 19,933 | 20,507 | 21,033 | 21,870 | 22,503 | 23,146 | 23,732 | 24,126 | 24,627 | 25,127 | 25,473 | 26,053 | 26,655 | 27,280 | 27,778 | 28,255 | 28,705 | 29,274 |
| 4,238 | 4,449 | 4,746 | 5,030 | 5,245 | 5,458 | 5,734 | 6,144 | 6,501 | 6,751 | 6,984 | 7,116 | 7,116 | 7,468 | 7,468 | 7,886 | 7,886 | 7,886 | 8,340 | 8,340 | 8,539 |
| 4,708 | 4,879 | 4,879 | 5,130 | 5,245 | 5,822 | 5,934 | 6,129 | 6,288 | 6,440 | 6,440 | 6,677 | 6,853 | 7,080 | 7,225 | 7,444 | 7,688 | 7,688 | 7,935 | 8,069 | 8,167 |
| 28,044 | 28,711 | 29,140 | 29,383 | 29,746 | 30,069 | 30,399 | 30,725 | 30,928 | 31,490 | 31,673 | 31,881 | 32,116 | 32,728 | 33,555 | 33,903 | 34,183 | 34,498 | 34,832 | 35,161 | 35,439 |
| 62,205 | 64,311 | 66,263 | 68,087 | 69,087 | 70,271 | 72,025 | 73,721 | 75,333 | 76,743 | 77,793 | 78,462 | 79,332 | 80,497 | 82,182 | 83,421 | 84,933 | 86,010 | 87,052 | 87,925 | 88,970 |
| 23,472 | 23,472 | 24,473 | 26,408 | 27,117 | 28,163 | 28,163 | 30,485 | 31,534 | 32,587 | 33,373 | 34,061 | 34,812 | 35,903 | 36,986 | 37,968 | 38,804 | 38,804 | 41,546 | 42,323 | 43,531 |
| 1,123 | 1,123 | 1,152 | 1,205 | 1,226 | 1,254 | 1,254 | 1,374 | 1,374 | 1,434 | 1,457 | 1,462 | 1,515 | 1,515 | 1,603 | 1,648 | 1,687 | 1,713 | 1,741 | 1,819 | 1,877 |
| 41,379 | 42,356 | 43,207 | 43,754 | 43,950 | 44,397 | 45,054 | 45,646 | 46,326 | 46,756 | 47,138 | 47,552 | 48,021 | 48,391 | 49,582 | 50,079 | 50,504 | 51,142 | 51,915 | 52,350 | 53,009 |
| 5,136 | 5,730 | 6,228 | 7,238 | 7,234 | 7,851 | 8,579 | 10,088 | 10,088 | 11,271 | 11,271 | 11,799 | 12,917 | 12,917 | 14,240 | 14,240 | 15,668 | 16,372 | 17,029 | 17,670 | 17,670 |
| 7,562 | 7,835 | 8,154 | 8,386 | 8,754 | 8,916 | 9,102 | 9,341 | 9,489 | 9,666 | 9,844 | 9,918 | 10,006 | 10,142 | 10,316 | 10,456 | 10,675 | 10,789 | 10,945 | 11,080 | 11,232 |
| 7,212 | 7,212 | 7,441 | 7,877 | 8,207 | 8,422 | 8,424 | 9,090 | 9,378 | 9,501 | 9,674 | 9,908 | 10,093 | 10,483 | 10,801 | 11,123 | 11,296 | 11,432 | 11,704 | 11,967 | 12,222 |
| 453 | 453 | 455 | 457 | 457 | 456 | 456 | 459 | 458 | 458 | 458 | 459 | 459 | 462 | 466 | 466 | 468 | 470 | 470 | 478 | 478 |
| 10,509 | 10,923 | 11,509 | 11,664 | 11,848 | 12,256 | 12,758 | 13,397 | 13,868 | 14,360 | 14,764 | 15,045 | 15,346 | 15,816 | 16,507 | 17,129 | 17,982 | 18,512 | 19,023 | 19,700 | 20,122 |
| 1,067 | 1,107 | 1,153 | 1,191 | 1,225 | 1,266 | 1,323 | 1,371 | 1,425 | 1,464 | 1,491 | 1,518 | 1,571 | 1,647 | 1,712 | 1,761 | 1,848 | 1,900 | 1,931 | 1,994 | 2,095 |
| 4,281 | 4,838 | 5,326 | 5,659 | 6,083 | 6,438 | 6,771 | 7,190 | 7,831 | 8,234 | 8,315 | 8,572 | 8,692 | 9,075 | 9,416 | 9,772 | 10,220 | 10,348 | 10,625 | 10,846 | 11,122 |
| 2,146 | 2,310 | 2,429 | 2,518 | 2,588 | 2,636 | 2,740 | 2,843 | 2,947 | 3,011 | 3,071 | 3,160 | 3,239 | 3,299 | 3,382 | 3,464 | 3,556 | 3,596 | 3,652 | 3,721 | 3,868 |
| 118,652 | 121,190 | 123,717 | 126,744 | 128,269 | 130,593 | 131,890 | 133,635 | 135,454 | 137,085 | 138,532 | 139,945 | 140,743 | 141,560 | 142,704 | 143,905 | 145,089 | 146,334 | 148,039 | 149,013 | 150,399 |
| 3,411 | 3,513 | 3,732 | 3,850 | 4,031 | 4,138 | 4,291 | 4,493 | 4,673 | 4,778 | 4,863 | 5,069 | 5,212 | 5,364 | 5,503 | 5,662 | 5,847 | 5,938 | 6,096 | 6,192 | 6,317 |
| 5,007 | 5,227 | 5,311 | 5,426 | 5,500 | 5,603 | 5,663 | 5,766 | 5,884 | 6,028 | 6,109 | 6,163 | 6,322 | 6,405 | 6,499 | 6,629 | 6,662 | 6,872 | 6,921 | 7,046 | 7,182 |
| 299106 | 303129 | 307506 | 312618 | 313836 | 316041 | 318915 | 321962 | 324710 | 328599 | 334640 | 336017 | 336681 | 338617 | 340799 | 343304 | 351872 | 347936 | 349214 | 356179 | 357431 |
| 18,027 | 18,743 | 19,335 | 19,914 | 20,474 | 20,969 | 21,576 | 22,131 | 23,016 | 23,697 | 24,081 | 24,777 | 25,250 | 25,721 | 26,357 | 26,954 | 27,474 | 27,923 | 28,272 | 28,952 | 29,436 |
| 3,618 | 3,748 | 3,851 | 3,972 | 4,044 | 4,054 | 4,201 | 4,210 | 4,210 | 4,335 | 4,589 | 4,439 | 4,439 | 4,741 | 4,861 | 4,971 | 4,861 | 5,310 | 5,247 | 5,407 | 5,407 |
| 2,510 | 2,579 | 2,635 | 2,680 | 2,759 | 2,839 | 2,916 | 2,989 | 3,068 | 3,160 | 3,228 | 3,286 | 3,358 | 3,416 | 3,479 | 3,541 | 3,612 | 3,623 | 3,687 | 3,726 | 3,801 |
| 45,763 | 46,971 | 48,305 | 49,267 | 50,092 | 50,957 | 51,845 | 52,915 | 54,238 | 55,316 | 56,611 | 57,154 | 57,991 | 58,698 | 59,636 | 60,622 | 61,611 | 62,234 | 63,666 | 63,936 | 64,412 |
| 8,621 | 8,962 | 9,289 | 9,477 | 9,652 | 9,933 | 10,205 | 10,530 | 10,779 | 10,989 | 11,274 | 11,450 | 11,614 | 11,835 | 12,016 | 12,219 | 12,434 | 12,674 | 12,795 | 12,951 | 13,356 |
| 6,095 | 6,258 | 6,489 | 6,626 | 6,757 | 6,841 | 6,936 | 7,142 | 7,367 | 7,531 | 7,653 | 7,792 | 7,927 | 8,030 | 8,189 | 8,407 | 8,661 | 8,816 | 8,942 | 9,056 | 9,175 |
| 2,449 | 2,525 | 2,588 | 2,631 | 2,668 | 2,721 | 2,779 | 2,905 | 3,144 | 3,393 | 3,517 | 3,614 | 3,663 | 3,732 | 3,792 | 3,887 | 3,959 | 3,987 | 4,027 | 4,085 | 4,177 |
| 10,735 | 11,891 | 12,661 | 13,243 | 13,571 | 13,690 | 13,938 | 14,166 | 14,512 | 14,843 | 15,060 | 15,622 | 16,190 | 16,451 | 16,784 | 17,052 | 17,374 | 17,495 | 18,121 | 18,491 | 18,646 |
| 28,087 | 29,229 | 30,552 | 31,548 | 32,332 | 33,369 | 34,422 | 35,390 | 36,609 | 37,860 | 38,869 | 39,869 | 41,048 | 42,403 | 43,851 | 45,198 | 46,999 | 47,784 | 48,693 | 49,912 | 51,323 |
| 4,753 | 4,828 | 4,981 | 5,261 | 5,406 | 5,530 | 5,595 | 5,794 | 6,041 | 6,234 | 6,317 | 6,395 | 6,548 | 6,675 | 6,837 | 7,012 | 7,166 | 7,305 | 7,459 | 7,595 | 7,808 |
| 16,901 | 16,901 | 17,731 | 19,492 | 19,492 | 20,256 | 20,256 | 22,342 | 22,342 | 23,196 | 25,070 | 25,800 | 26,746 | 27,813 | 28,672 | 29,683 | 30,388 | 31,140 | 32,145 | 32,908 | 34,137 |
| 866 | 879 | 886 | 897 | 902 | 906 | 908 | 916 | 918 | 921 | 927 | 926 | 927 | 929 | 933 | 933 | 932 | 940 | 940 | 944 | 945 |
| 14,327 | 14,637 | 15,003 | 15,185 | 15,462 | 15,594 | 15,905 | 16,231 | 16,388 | 16,674 | 16,891 | 17,122 | 17,330 | 17,512 | 17,773 | 17,951 | 18,287 | 18,433 | 18,611 | 18,811 | 18,971 |
| 6,854 | 7,314 | 7,660 | 7,964 | 8,236 | 8,566 | 8,901 | 9,215 | 9,590 | 9,939 | 10,219 | 10,418 | 10,611 | 10,902 | 11,275 | 11,685 | 12,187 | 12,543 | 12,687 | 12,885 | 13,431 |
| 1,125 | 1,151 | 1,184 | 1,195 | 1,224 | 1,242 | 1,276 | 1,297 | 1,323 | 1,347 | 1,362 | 1,369 | 1,378 | 1,404 | 1,434 | 1,447 | 1,470 | 1,490 | 1,502 | 1,514 | 1,567 |
| 559 | 566 | 579 | 586 | 596 | 604 | 631 | 635 | 644 | 653 | 662 | 669 | 675 | 688 | 701 | 716 | 741 | 754 | 766 | 776 | 787 |

| 5/21/2020 | 5/22/2020 | 5/23/2020 | 5/24/2020 | 5/25/2020 | 5/26/2020 | 5/27/2020 | 5/28/2020 | 5/29/2020 | 5/30/2020 | 5/31/2020 | 6/1/2020 | 6/2/2020 | 6/3/2020 | 6/4/2020 | 6/5/2020 | 6/6/2020 | 6/7/2020 | 6/8/2020 | 6/9/2020 | 6/10/2020 |
|---|---|---|---|---|---|---|---|---|---|---|---|---|---|---|---|---|---|---|---|---|
| 402 | 404 | 408 | 408 | 409 | 411 | 412 | 425 | 430 | 434 | 460 | 467 | 487 | 505 | 513 | 524 | 536 | 544 | 563 | 573 | 593 |
| 13,647 | 13,941 | 14,388 | 14,741 | 15,257 | 15,805 | 16,262 | 16,729 | 17,322 | 17,649 | 18,245 | 18,438 | 18,771 | 18,851 | 19,082 | 19,396 | 20,054 | 20,500 | 21,430 | 21,422 | 22,000 |
| 5,458 | 5,612 | 5,775 | 5,922 | 6,029 | 6,180 | 6,277 | 6,538 | 6,777 | 7,013 | 7,253 | 7,443 | 7,818 | 8,067 | 8,425 | 8,651 | 9,101 | 9,426 | 9,740 | 10,080 | 10,368 |
| 15,315 | 15,608 | 16,039 | 16,339 | 16,561 | 16,783 | 17,262 | 17,763 | 18,465 | 19,255 | 19,936 | 20,123 | 21,250 | 22,223 | 22,753 | 24,332 | 25,451 | 26,889 | 27,678 | 28,296 | 29,852 |
| 86,197 | 88,444 | 90,631 | 92,710 | 94,558 | 96,733 | 98,980 | 101,697 | 103,886 | 106,878 | 110,583 | 113,006 | 115,310 | 117,687 | 119,807 | 122,901 | 126,016 | 128,812 | 131,319 | 133,489 | 136,191 |
| 23,121 | 23,455 | 23,937 | 24,137 | 24,226 | 24,503 | 24,995 | 25,121 | 25,613 | 26,098 | 26,378 | 26,577 | 26,788 | 27,060 | 27,360 | 27,615 | 27,848 | 28,001 | 28,183 | 28,347 | 28,499 |
| 39,208 | 39,640 | 40,022 | 40,468 | 40,873 | 41,303 | 41,288 | 41,559 | 41,762 | 42,022 | 42,201 | 42,740 | 42,979 | 43,091 | 43,239 | 43,460 | 43,818 | 43,968 | 44,092 | 44,179 | 44,347 |
| 7,788 | 7,893 | 7,966 | 8,110 | 8,225 | 8,334 | 8,406 | 8,492 | 8,538 | 8,717 | 8,801 | 8,857 | 8,886 | 9,016 | 9,120 | 9,199 | 9,269 | 9,332 | 9,389 | 9,474 | 9,537 |
| 8,529 | 8,690 | 8,690 | 8,809 | 9,067 | 9,096 | 9,171 | 9,236 | 9,422 | 9,498 | 9,606 | 9,685 | 9,712 | 9,746 | 9,773 | 9,845 | 9,942 | 9,972 | 10,020 | 10,056 | 10,107 |
| 48,675 | 49,451 | 50,127 | 50,867 | 51,746 | 52,255 | 52,634 | 51,918 | 53,114 | 54,029 | 54,764 | 55,415 | 56,001 | 57,293 | 58,701 | 59,993 | 61,246 | 62,416 | 63,378 | 64,448 | 65,779 |
| 40,460 | 41,482 | 42,145 | 42,846 | 43,369 | 43,794 | 44,447 | 45,226 | 45,711 | 46,301 | 47,009 | 47,656 | 48,207 | 48,894 | 49,847 | 50,621 | 51,309 | 51,898 | 52,497 | 53,249 | 53,980 |
| 596 | 594 | 595 | 595 | 595 | 595 | 596 | 601 | 603 | 606 | 607 | 607 | 610 | 616 | 619 | 625 | 634 | 636 | 637 | 644 | 647 |
| 16,170 | 16,504 | 16,898 | 17,250 | 17,577 | 17,703 | 18,357 | 18,585 | 18,956 | 19,244 | 19,552 | 19,698 | 20,015 | 20,108 | 20,804 | 21,146 | 21,478 | 21,588 | 21,988 | 22,237 | 22,603 |
| 2,534 | 2,595 | 2,626 | 2,626 | 2,684 | 2,699 | 2,731 | 2,769 | 2,803 | 2,839 | 2,839 | 2,906 | 2,933 | 2,990 | 3,054 | 3,111 | 3,139 | 3,139 | 3,189 | 3,220 | 3,260 |
| 102,686 | 105,444 | 107,796 | 110,304 | 112,017 | 113,195 | 114,306 | 115,833 | 117,455 | 118,917 | 120,260 | 121,234 | 122,848 | 123,830 | 124,759 | 125,915 | 126,890 | 127,757 | 129,139 | 129,936 | 130,561 |
| 29,936 | 30,409 | 30,901 | 31,376 | 31,715 | 32,078 | 32,437 | 33,068 | 33,558 | 34,211 | 34,574 | 34,830 | 35,237 | 35,712 | 36,096 | 36,578 | 36,997 | 37,397 | 37,623 | 38,033 | 38,337 |
| 8,539 | 8,958 | 8,958 | 8,958 | 9,218 | 9,218 | 9,337 | 9,337 | 9,719 | 9,719 | 9,719 | 10,011 | 10,011 | 10,170 | 10,170 | 10,393 | 10,393 | 10,393 | 10,650 | 10,650 | 10,812 |
| 8,286 | 8,426 | 8,571 | 8,571 | 8,571 | 8,951 | 9,077 | 9,184 | 9,464 | 9,704 | 9,704 | 10,046 | 10,185 | 10,410 | 10,705 | 10,977 | 11,287 | 11,287 | 11,476 | 11,708 | 11,883 |
| 36,627 | 37,048 | 37,163 | 37,292 | 37,914 | 38,159 | 38,602 | 38,907 | 38,907 | 39,682 | 39,916 | 40,452 | 40,857 | 41,133 | 41,562 | 41,989 | 42,486 | 42,816 | 43,050 | 43,612 | 44,030 |
| 90,084 | 90,899 | 91,662 | 92,675 | 93,271 | 93,693 | 94,220 | 94,895 | 95,512 | 96,301 | 96,965 | 100,805 | 101,163 | 101,592 | 102,063 | 102,557 | 103,132 | 103,436 | 103,626 | 103,889 | 104,156 |
| 44,424 | 45,495 | 46,313 | 47,152 | 47,687 | 48,423 | 49,709 | 50,988 | 52,015 | 52,778 | 53,327 | 54,175 | 54,982 | 55,858 | 56,770 | 57,482 | 57,973 | 58,404 | 58,904 | 59,465 | 60,197 |
| 1,948 | 1,948 | 2,013 | 2,074 | 2,109 | 2,109 | 2,189 | 2,226 | 2,282 | 2,325 | 2,349 | 2,377 | 2,418 | 2,418 | 2,446 | 2,482 | 2,570 | 2,570 | 2,588 | 2,606 | 2,667 |
| 53,510 | 53,913 | 54,365 | 54,679 | 54,881 | 55,104 | 55,608 | 56,014 | 56,621 | 56,884 | 57,397 | 57,532 | 57,731 | 58,035 | 58,241 | 63,539 | 63,983 | 64,413 | 64,701 | 64,998 | 65,182 |
| 19,005 | 19,845 | 19,845 | 20,573 | 21,960 | 22,464 | 22,464 | 23,531 | 23,531 | 24,850 | 25,208 | 25,208 | 25,508 | 25,870 | 26,273 | 27,501 | 27,886 | 28,224 | 28,523 | 28,869 | 29,316 |
| 11,340 | 11,558 | 11,752 | 11,988 | 12,167 | 12,291 | 12,492 | 12,673 | 12,795 | 12,962 | 13,147 | 13,327 | 13,575 | 13,767 | 14,057 | 14,253 | 14,442 | 14,553 | 14,734 | 14,913 | 15,187 |
| 12,624 | 13,005 | 13,252 | 13,458 | 13,731 | 14,044 | 14,372 | 14,790 | 15,230 | 15,501 | 15,752 | 16,021 | 16,322 | 16,560 | 16,560 | 16,769 | 17,270 | 17,768 | 17,768 | 18,483 | 18,483 |
| 479 | 479 | 479 | 479 | 479 | 479 | 485 | 493 | 505 | 505 | 515 | 523 | 525 | 539 | 539 | 540 | 540 | 548 | 548 | 554 | 561 |
| 20,860 | 21,618 | 22,725 | 23,222 | 23,964 | 24,140 | 24,628 | 25,412 | 26,488 | 27,673 | 28,589 | 29,263 | 29,889 | 30,777 | 31,966 | 33,255 | 34,625 | 35,555 | 36,484 | 37,160 | 38,171 |
| 2,229 | 2,317 | 2,365 | 2,418 | 2,457 | 2,422 | 2,439 | 2,481 | 2,520 | 2,554 | 2,577 | 2,625 | 2,646 | 2,679 | 2,706 | 2,745 | 2,816 | 2,861 | 2,880 | 2,901 | 2,941 |
| 11,425 | 11,662 | 11,989 | 12,134 | 12,355 | 12,619 | 12,976 | 13,261 | 13,654 | 13,905 | 14,101 | 14,345 | 14,611 | 14,866 | 15,117 | 15,379 | 15,543 | 15,634 | 15,752 | 15,883 | 16,025 |
| 3,935 | 4,014 | 4,089 | 4,149 | 4,197 | 4,231 | 4,286 | 4,386 | 4,492 | 4,545 | 4,651 | 4,685 | 4,749 | 4,795 | 4,876 | 4,953 | 5,019 | 5,043 | 5,079 | 5,132 | 5,178 |
| 151,472 | 152,719 | 153,104 | 154,154 | 155,092 | 155,764 | 156,628 | 157,185 | 158,844 | 159,608 | 160,445 | 160,918 | 161,545 | 162,068 | 162,530 | 163,336 | 163,893 | 164,164 | 164,497 | 164,796 | 165,346 |
| 6,472 | 6,625 | 6,795 | 6,943 | 7,026 | 7,130 | 7,252 | 7,364 | 7,493 | 7,624 | 7,689 | 7,800 | 8,024 | 8,140 | 8,353 | 8,672 | 8,800 | 8,940 | 9,062 | 9,105 | 9,250 |
| 7,271 | 7,401 | 7,696 | 7,786 | 7,888 | 7,998 | 8,113 | 8,225 | 8,350 | 8,495 | 8,610 | 8,688 | 8,830 | 8,951 | 9,090 | 9,285 | 9,460 | 9,649 | 9,805 | 10,030 | 10,164 |
| 353623 | 355621 | 362859 | 358844 | 365405 | 366638 | 367994 | 369460 | 365323 | 366802 | 373022 | 368777 | 375224 | 371019 | 372046 | 373097 | 379322 | 380089 | 380737 | 381505 | 382136 |
| 30,167 | 30,794 | 31,408 | 31,911 | 32,477 | 33,006 | 33,439 | 33,915 | 34,566 | 35,034 | 35,513 | 35,984 | 36,413 | 36,792 | 37,282 | 37,758 | 38,111 | 38,476 | 38,837 | 39,162 | 39,575 |
| 5,503 | 5,503 | 5,543 | 5,691 | 5,860 | 5,974 | 6,051 | 6,104 | 6,238 | 6,280 | 6,280 | 6,347 | 6,427 | 6,582 | 6,707 | 6,820 | 6,921 | 6,921 | 7,072 | 7,163 | 7,218 |
| 3,817 | 3,864 | 3,888 | 3,927 | 3,949 | 3,967 | 4,038 | 4,086 | 4,131 | 4,185 | 4,243 | 4,302 | 4,335 | 4,399 | 4,474 | 4,570 | 4,662 | 4,808 | 4,922 | 4,988 | 5,060 |
| 66,258 | 66,983 | 67,713 | 68,186 | 68,637 | 69,417 | 70,042 | 70,735 | 71,415 | 71,926 | 72,282 | 72,894 | 73,405 | 73,942 | 74,385 | 75,086 | 75,592 | 75,943 | 76,436 | 76,846 | 77,313 |
| 13,571 | 13,736 | 13,952 | 14,065 | 14,065 | 14,210 | 14,353 | 14,494 | 14,635 | 14,819 | 14,928 | 14,991 | 15,112 | 15,219 | 15,325 | 15,441 | 15,441 | 15,441 | 15,642 | 15,691 | 15,756 |
| 9,379 | 9,638 | 9,895 | 10,096 | 10,178 | 10,416 | 10,623 | 10,788 | 11,131 | 11,394 | 11,861 | 12,148 | 12,415 | 12,651 | 13,005 | 13,453 | 13,916 | 14,286 | 14,800 | 15,228 | 15,759 |
| 4,250 | 4,356 | 4,468 | 4,563 | 4,586 | 4,653 | 4,710 | 4,793 | 4,866 | 4,960 | 4,993 | 5,034 | 5,067 | 5,162 | 5,247 | 5,277 | 5,367 | 5,438 | 5,471 | 5,523 | 5,604 |
| 19,073 | 19,510 | 19,909 | 20,269 | 20,731 | 21,089 | 21,441 | 21,822 | 22,236 | 22,720 | 23,159 | 23,709 | 24,543 | 24,990 | 25,289 | 25,690 | 26,248 | 26,561 | 27,129 | 27,759 | 28,061 |
| 52,268 | 53,449 | 54,509 | 55,348 | 55,971 | 56,560 | 57,921 | 59,776 | 61,006 | 62,338 | 64,287 | 64,880 | 66,568 | 68,271 | 69,920 | 71,613 | 73,553 | 74,978 | 75,616 | 77,253 | 79,757 |
| 7,981 | 8,179 | 8,331 | 8,500 | 8,584 | 8,658 | 8,815 | 9,137 | 9,429 | 9,688 | 9,944 | 10,101 | 10,353 | 10,737 | 11,015 | 11,252 | 11,939 | 12,117 | 12,429 | 12,704 | 13,048 |
| 34,950 | 35,749 | 36,244 | 37,727 | 39,342 | 40,249 | 41,401 | 42,533 | 43,611 | 44,607 | 45,398 | 46,239 | 46,905 | 47,856 | 48,532 | 49,397 | 50,681 | 51,251 | 51,738 | 52,177 | 52,647 |
| 950 | 952 | 954 | 956 | 962 | 967 | 971 | 974 | 975 | 977 | 981 | 983 | 988 | 990 | 1,025 | 1,027 | 1,045 | 1,063 | 1,075 | 1,084 | 1,094 |
| 19,117 | 19,265 | 19,585 | 19,828 | 20,065 | 20,181 | 20,406 | 20,764 | 21,071 | 21,349 | 21,702 | 21,977 | 22,157 | 22,484 | 22,729 | 22,993 | 23,422 | 23,729 | 24,041 | 24,354 | 24,642 |
| 13,885 | 14,396 | 14,877 | 15,277 | 15,584 | 15,863 | 16,462 | 16,974 | 17,707 | 18,230 | 18,403 | 18,543 | 18,917 | 19,400 | 19,892 | 20,249 | 20,571 | 20,835 | 21,038 | 21,308 | 21,593 |
| 1,603 | 1,705 | 1,729 | 1,771 | 1,782 | 1,854 | 1,899 | 1,935 | 1,972 | 1,989 | 2,010 | 2,028 | 2,056 | 2,077 | 2,102 | 2,119 | 2,136 | 2,144 | 2,161 | 2,179 | 2,193 |
| 801 | 803 | 813 | 838 | 843 | 850 | 860 | 876 | 891 | 898 | 903 | 910 | 912 | 915 | 921 | 933 | 939 | 947 | 960 | 970 | 980 |

| 6/11/2020 | 6/12/2020 | 6/13/2020 | 6/14/2020 | 6/15/2020 | 6/16/2020 | 6/17/2020 | 6/18/2020 | 6/19/2020 | 6/20/2020 | 6/21/2020 | 6/22/2020 | 6/23/2020 | 6/24/2020 | 6/25/2020 | 6/26/2020 | 6/27/2020 | 6/28/2020 | 6/29/2020 | 6/30/2020 |
|---|---|---|---|---|---|---|---|---|---|---|---|---|---|---|---|---|---|---|---|
| 610 | 625 | 654 | 661 | 664 | 676 | 696 | 708 | 723 | 743 | 755 | 761 | 778 | 792 | 816 | 836 | 854 | 883 | 904 | 940 |
| 22,845 | 23,710 | 25,368 | 26,287 | 26,713 | 26,912 | 27,327 | 28,214 | 29,017 | 29,549 | 30,021 | 30,477 | 31,234 | 32,086 | 33,232 | 34,220 | 35,105 | 36,350 | 37,203 | 38,064 |
| 10,816 | 11,547 | 12,095 | 12,501 | 12,917 | 13,191 | 13,606 | 13,928 | 14,631 | 15,142 | 15,561 | 16,083 | 16,678 | 17,375 | 18,062 | 18,740 | 19,310 | 19,818 | 20,257 | 20,777 |
| 31,264 | 32,918 | 34,458 | 35,691 | 36,705 | 39,097 | 40,924 | 43,443 | 46,689 | 49,798 | 52,390 | 54,586 | 58,179 | 59,974 | 63,030 | 66,458 | 70,051 | 73,908 | 74,533 | 79,215 |
| 139,281 | 141,983 | 145,643 | 148,855 | 151,452 | 153,560 | 157,015 | 161,099 | 165,416 | 169,309 | 173,824 | 178,054 | 183,073 | 190,222 | 195,571 | 200,461 | 206,433 | 211,243 | 216,550 | 222,917 |
| 28,647 | 28,822 | 29,017 | 29,130 | 29,299 | 29,442 | 29,673 | 29,901 | 30,187 | 30,349 | 30,539 | 30,705 | 30,893 | 31,155 | 31,479 | 31,796 | 32,022 | 32,307 | 32,511 | 32,715 |
| 44,461 | 44,689 | 44,994 | 45,088 | 45,235 | 45,349 | 45,429 | 45,440 | 45,557 | 45,715 | 45,755 | 45,782 | 45,899 | 45,913 | 45,994 | 46,059 | 46,206 | 46,303 | 46,362 | 46,514 |
| 9,589 | 9,654 | 9,709 | 9,767 | 9,799 | 9,818 | 9,847 | 9,903 | 9,952 | 9,984 | 10,020 | 10,058 | 10,094 | 10,128 | 10,159 | 10,185 | 10,216 | 10,248 | 10,292 | 10,327 |
| 10,173 | 10,229 | 10,264 | 10,340 | 10,403 | 10,444 | 10,500 | 10,611 | 10,611 | 10,775 | 10,822 | 10,847 | 10,889 | 10,980 | 11,017 | 11,107 | 11,253 | 11,416 | 11,376 | 11,510 |
| 67,456 | 69,341 | 71,589 | 73,650 | 75,388 | 78,128 | 80,676 | 83,854 | 87,643 | 91,670 | 95,139 | 98,047 | 101,303 | 106,743 | 111,724 | 120,574 | 130,092 | 138,567 | 143,805 | 149,781 |
| 54,973 | 55,783 | 56,801 | 57,681 | 58,414 | 59,078 | 60,030 | 60,912 | 62,009 | 63,809 | 64,701 | 65,928 | 67,675 | 69,381 | 71,095 | 72,995 | 74,985 | 77,210 | 79,417 | 81,291 |
| 654 | 663 | 680 | 688 | 696 | 674 | 678 | 718 | 746 | 760 | 769 | 771 | 775 | 766 | 782 | 796 | 802 | 828 | 829 | 842 |
| 22,973 | 23,348 | 23,717 | 23,926 | 24,077 | 24,179 | 24,461 | 24,854 | 25,275 | 25,496 | 25,957 | 26,051 | 26,372 | 26,705 | 27,257 | 27,685 | 28,012 | 28,489 | 28,782 | 28,947 |
| 3,302 | 3,353 | 3,399 | 3,399 | 3,462 | 3,540 | 3,632 | 3,743 | 3,871 | 4,006 | 4,006 | 4,254 | 4,402 | 4,645 | 4,865 | 5,148 | 5,319 | 5,319 | 5,752 | 6,117 |
| 131,327 | 132,059 | 132,732 | 133,404 | 133,877 | 134,500 | 135,046 | 135,639 | 136,470 | 137,104 | 137,762 | 138,224 | 138,825 | 139,540 | 140,434 | 141,344 | 142,130 | 142,776 | 143,514 | 144,238 |
| 38,748 | 39,146 | 39,543 | 39,909 | 40,430 | 40,786 | 41,013 | 41,438 | 41,746 | 42,061 | 42,423 | 42,633 | 42,871 | 43,140 | 43,655 | 44,140 | 44,575 | 44,930 | 45,228 | 45,594 |
| 10,812 | 11,047 | 11,047 | 11,047 | 11,419 | 11,419 | 11,681 | 11,681 | 12,059 | 12,059 | 12,059 | 12,465 | 12,465 | 12,970 | 12,970 | 13,538 | 13,538 | 13,538 | 14,443 | 14,443 |
| 11,945 | 12,166 | 12,445 | 12,445 | 12,647 | 12,829 | 12,995 | 13,197 | 13,454 | 13,630 | 13,750 | 13,839 | 14,141 | 14,363 | 14,617 | 14,859 | 15,167 | 15,232 | 15,347 | 15,624 |
| 44,472 | 44,995 | 46,283 | 46,619 | 47,172 | 47,706 | 48,634 | 48,634 | 48,515 | 49,385 | 49,778 | 50,239 | 51,595 | 52,477 | 53,415 | 54,769 | 54,769 | 56,236 | 57,081 | 58,095 |
| 104,667 | 105,059 | 105,395 | 105,603 | 105,690 | 105,885 | 106,151 | 106,422 | 106,650 | 106,936 | 107,061 | 107,210 | 107,439 | 107,611 | 107,837 | 108,070 | 108,443 | 108,667 | 108,768 | 108,882 |
| 60,613 | 61,305 | 61,701 | 62,032 | 62,409 | 62,969 | 63,229 | 63,548 | 63,956 | 64,306 | 64,603 | 65,007 | 65,337 | 65,777 | 66,115 | 66,450 | 66,777 | 67,254 | 67,559 | 67,918 |
| 2,667 | 2,721 | 2,757 | 2,810 | 2,819 | 2,819 | 2,836 | 2,878 | 2,938 | 2,938 | 2,957 | 2,971 | 2,994 | 3,071 | 3,070 | 3,102 | 3,191 | 3,221 | 3,252 | 3,294 |
| 65,449 | 65,672 | 65,836 | 66,054 | 66,085 | 66,269 | 66,497 | 66,798 | 67,097 | 67,545 | 67,711 | 67,957 | 68,197 | 68,555 | 68,989 | 69,329 | 69,679 | 69,946 | 70,223 | 70,728 |
| 29,316 | 29,795 | 30,172 | 30,693 | 30,882 | 31,296 | 31,296 | 31,675 | 32,467 | 32,920 | 33,227 | 33,469 | 33,763 | 33,763 | 34,616 | 34,616 | 35,549 | 35,546 | 35,861 | 36,716 |
| 15,390 | 15,585 | 15,810 | 15,810 | 16,189 | 16,414 | 16,625 | 16,908 | 17,201 | 17,590 | 18,005 | 18,143 | 18,577 | 18,868 | 19,421 | 19,914 | 20,261 | 20,575 | 21,043 | 21,551 |
| 18,483 | 19,348 | 19,348 | 19,799 | 20,152 | 20,641 | 20,641 | 20,641 | 20,641 | 20,641 | 20,641 | 22,287 | 22,898 | 23,424 | 24,516 | 25,531 | 25,892 | 26,567 | 27,247 | 27,900 |
| 573 | 588 | 588 | 609 | 612 | 630 | 630 | 666 | 698 | 716 | 717 | 734 | 766 | 803 | 829 | 852 | 852 | 919 | 967 | 1,018 |
| 39,481 | 41,249 | 42,676 | 44,119 | 45,102 | 45,853 | 46,855 | 48,188 | 49,840 | 51,389 | 52,801 | 53,605 | 54,453 | 56,174 | 57,183 | 58,818 | 60,537 | 62,142 | 63,484 | 64,670 |
| 2,980 | 3,016 | 3,058 | 3,080 | 3,101 | 3,124 | 3,166 | 3,193 | 3,226 | 3,251 | 3,288 | 3,313 | 3,320 | 3,362 | 3,393 | 3,421 | 3,458 | 3,495 | 3,539 | 3,576 |
| 16,315 | 16,513 | 16,633 | 16,725 | 16,851 | 17,031 | 17,226 | 17,415 | 17,591 | 17,707 | 17,810 | 17,957 | 18,092 | 18,221 | 18,346 | 18,524 | 18,775 | 18,899 | 19,042 | 19,177 |
| 5,209 | 5,251 | 5,299 | 5,318 | 5,345 | 5,364 | 5,436 | 5,450 | 5,450 | 5,518 | 5,544 | 5,558 | 5,571 | 5,598 | 5,638 | 5,638 | 5,671 | 5,747 | 5,760 | 5,782 |
| 165,816 | 166,164 | 166,603 | 166,881 | 167,103 | 167,426 | 167,703 | 168,107 | 168,496 | 168,834 | 169,142 | 169,415 | 169,734 | 169,892 | 170,196 | 170,584 | 170,873 | 171,182 | 171,272 | 171,667 |
| 9,367 | 9,526 | 9,621 | 9,723 | 9,845 | 9,933 | 10,065 | 10,153 | 10,260 | 10,430 | 10,565 | 10,694 | 10,838 | 10,990 | 11,192 | 11,408 | 11,619 | 11,809 | 11,982 | 12,147 |
| 10,417 | 10,678 | 10,964 | 11,173 | 11,297 | 11,658 | 11,860 | 12,094 | 12,515 | 13,160 | 13,434 | 13,764 | 14,228 | 14,592 | 15,085 | 15,472 | 16,576 | 17,397 | 18,131 | 18,684 |
| 382913 | 378606 | 379459 | 380052 | 380734 | 386489 | 386990 | 387645 | 388330 | 389213 | 389737 | 390326 | 390894 | 391510 | 392292 | 393041 | 393728 | 394250 | 394667 | 395294 |
| 40,004 | 40,424 | 40,848 | 41,148 | 41,576 | 42,010 | 42,422 | 43,122 | 43,731 | 44,262 | 44,808 | 45,537 | 46,127 | 46,759 | 47,651 | 48,638 | 49,455 | 50,309 | 51,046 | 51,789 |
| 7,378 | 7,496 | 7,640 | 8,231 | 8,251 | 8,437 | 8,665 | 8,925 | 9,378 | 9,730 | 10,062 | 10,545 | 10,759 | 10,759 | 11,948 | 11,975 | 12,367 | 12,675 | 12,977 | 13,217 |
| 5,237 | 5,377 | 5,535 | 5,636 | 5,820 | 6,098 | 6,218 | 6,366 | 6,572 | 6,750 | 6,937 | 7,083 | 7,274 | 7,444 | 7,568 | 7,818 | 8,094 | 8,341 | 8,485 | 8,656 |
| 77,999 | 78,462 | 78,462 | 79,121 | 79,483 | 79,818 | 80,236 | 80,762 | 81,266 | 81,730 | 82,186 | 82,186 | 83,191 | 83,770 | 84,370 | 84,991 | 85,496 | 85,988 | 86,606 | 87,242 |
| 15,862 | 15,947 | 15,947 | 15,947 | 16,093 | 16,164 | 16,213 | 16,269 | 16,337 | 16,337 | 16,337 | 16,459 | 16,533 | 16,606 | 16,640 | 16,661 | 16,661 | 16,661 | 16,764 | 16,813 |
| 16,441 | 17,170 | 17,955 | 18,795 | 19,378 | 19,990 | 20,556 | 21,548 | 22,631 | 23,786 | 24,693 | 25,701 | 26,613 | 27,897 | 29,022 | 30,335 | 31,939 | 33,320 | 34,644 | 36,399 |
| 5,665 | 5,742 | 5,833 | 5,898 | 5,928 | 5,966 | 6,050 | 6,109 | 6,158 | 6,225 | 6,297 | 6,326 | 6,353 | 6,419 | 6,479 | 6,535 | 6,626 | 6,681 | 6,716 | 6,764 |
| 28,538 | 29,126 | 29,541 | 30,432 | 31,160 | 31,830 | 32,143 | 32,829 | 34,017 | 34,446 | 35,102 | 35,553 | 36,303 | 37,235 | 38,034 | 39,444 | 40,172 | 40,172 | 42,297 | 43,509 |
| 81,583 | 83,680 | 86,011 | 87,854 | 89,108 | 93,206 | 96,335 | 99,851 | 103,305 | 107,735 | 111,601 | 114,881 | 120,370 | 125,921 | 131,917 | 137,624 | 143,371 | 148,723 | 153,011 | 159,986 |
| 13,429 | 13,789 | 14,184 | 14,469 | 14,826 | 15,141 | 15,539 | 16,199 | 16,826 | 17,400 | 17,825 | 18,135 | 18,660 | 19,185 | 19,816 | 20,481 | 20,986 | 21,450 | 22,263 | 22,746 |
| 53,211 | 53,869 | 54,506 | 54,886 | 55,331 | 55,775 | 56,238 | 56,793 | 57,443 | 57,994 | 58,465 | 58,994 | 59,514 | 59,946 | 60,570 | 61,247 | 61,736 | 62,189 | 62,787 | 62,787 |
| 1,109 | 1,119 | 1,125 | 1,127 | 1,128 | 1,130 | 1,129 | 1,135 | 1,144 | 1,147 | 1,159 | 1,163 | 1,164 | 1,184 | 1,191 | 1,198 | 1,200 | 1,202 | 1,208 | 1,208 |
| 24,779 | 25,171 | 25,538 | 25,834 | 26,158 | 26,531 | 26,784 | 27,192 | 27,601 | 28,225 | 28,680 | 28,870 | 29,386 | 29,869 | 30,367 | 30,855 | 31,404 | 31,752 | 32,253 | 32,824 |
| 21,926 | 24,722 | 25,031 | 25,295 | 25,480 | 25,789 | 26,079 | 26,542 | 26,852 | 27,294 | 27,587 | 27,838 | 28,127 | 28,593 | 29,105 | 29,668 | 30,227 | 30,707 | 31,033 | 31,662 |
| 2,217 | 2,249 | 2,274 | 2,290 | 2,322 | 2,341 | 2,376 | 2,418 | 2,468 | 2,500 | 2,543 | 2,571 | 2,593 | 2,629 | 2,694 | 2,730 | 2,782 | 2,832 | 2,870 | 2,905 |
| 1,009 | 1,027 | 1,050 | 1,060 | 1,079 | 1,089 | 1,114 | 1,144 | 1,173 | 1,179 | 1,197 | 1,230 | 1,254 | 1,282 | 1,326 | 1,368 | 1,392 | 1,417 | 1,450 | 1,487 |

| 7/1/2020 | 7/2/2020 | 7/3/2020 | 7/4/2020 | 7/5/2020 | 7/6/2020 | 7/7/2020 | 7/8/2020 | 7/9/2020 | 7/10/2020 | 7/11/2020 | 7/12/2020 | 7/13/2020 | 7/14/2020 | 7/15/2020 | 7/16/2020 | 7/17/2020 | 7/18/2020 | 7/19/2020 | 7/20/2020 | 7/21/2020 |
|---|---|---|---|---|---|---|---|---|---|---|---|---|---|---|---|---|---|---|---|---|
| 978 | 1,017 | 1,063 | 1,111 | 1,138 | 1,166 | 1,184 | 1,226 | 1,272 | 1,323 | 1,385 | 1,479 | 1,539 | 1,579 | 1,631 | 1,693 | 1,733 | 1,795 | 1,874 | 1,949 | 2,041 |
| 38,981 | 40,111 | 41,865 | 42,862 | 44,909 | 44,909 | 45,821 | 46,991 | 49,205 | 50,546 | 51,947 | 53,587 | 55,590 | 57,308 | 59,113 | 61,143 | 63,153 | 65,234 | 67,011 | 68,950 | 70,426 |
| 21,197 | 22,075 | 22,622 | 23,209 | 23,814 | 24,253 | 24,512 | 25,246 | 26,052 | 26,803 | 27,864 | 28,367 | 28,939 | 29,733 | 30,297 | 31,114 | 31,762 | 32,533 | 33,228 | 33,927 | 34,665 |
| 84,092 | 87,425 | 91,858 | 94,553 | 98,089 | 101,441 | 105,094 | 108,614 | 112,671 | 116,892 | 119,930 | 122,467 | 123,824 | 128,097 | 131,354 | 134,613 | 138,523 | 141,265 | 143,624 | 145,183 | 148,683 |
| 232,657 | 240,195 | 248,235 | 254,745 | 260,155 | 271,684 | 277,774 | 289,468 | 296,499 | 304,297 | 312,344 | 320,804 | 329,162 | 336,508 | 347,634 | 356,178 | 366,164 | 375,363 | 384,692 | 391,538 | 400,769 |
| 33,029 | 33,352 | 33,612 | 33,866 | 34,065 | 34,257 | 34,664 | 35,116 | 35,525 | 36,191 | 36,591 | 36,913 | 37,242 | 37,686 | 38,155 | 38,726 | 39,344 | 39,788 | 40,142 | 40,566 | 41,059 |
| 46,572 | 46,646 | 46,717 | 46,717 | 46,717 | 46,976 | 47,033 | 47,108 | 47,209 | 47,287 | 47,287 | 47,287 | 47,510 | 47,530 | 47,636 | 47,750 | 47,893 | 47,893 | 47,893 | 48,055 | 48,096 |
| 10,365 | 10,390 | 10,435 | 10,447 | 10,482 | 10,515 | 10,569 | 10,642 | 10,679 | 10,743 | 10,801 | 10,847 | 10,906 | 10,946 | 11,026 | 11,076 | 11,115 | 11,194 | 11,261 | 11,339 | 11,427 |
| 11,739 | 11,731 | 11,923 | 12,128 | 12,348 | 12,469 | 12,525 | 12,610 | 12,751 | 12,857 | 12,978 | 12,879 | 12,969 | 13,050 | 13,114 | 13,337 | 13,429 | 13,429 | 13,519 | 13,746 | 13,792 |
| 156,288 | 166,303 | 175,718 | 187,090 | 197,076 | 203,376 | 210,594 | 220,476 | 229,367 | 240,710 | 250,984 | 266,119 | 278,667 | 287,789 | 297,876 | 311,640 | 323,002 | 333,201 | 345,612 | 355,899 | 365,244 |
| 84,237 | 87,709 | 90,493 | 93,319 | 95,516 | 97,064 | 100,470 | 103,890 | 106,727 | 111,211 | 114,401 | 116,926 | 120,569 | 123,963 | 127,834 | 131,275 | 135,183 | 139,872 | 143,123 | 145,575 | 148,988 |
| 851 | 870 | 894 | 915 | 939 | 946 | 980 | 1,001 | 1,036 | 1,058 | 1,097 | 1,115 | 1,138 | 1,158 | 1,190 | 1,209 | 1,232 | 1,245 | 1,273 | 1,287 | 1,310 |
| 29,458 | 30,209 | 30,430 | 31,061 | 31,353 | 31,657 | 32,029 | 32,509 | 33,243 | 33,896 | 34,647 | 35,171 | 35,529 | 35,850 | 36,294 | 37,132 | 37,722 | 38,041 | 38,723 | 39,224 | 39,473 |
| 6,370 | 6,593 | 6,994 | 7,370 | 7,733 | 8,052 | 8,539 | 8,969 | 9,428 | 9,928 | 10,505 | 10,902 | 11,402 | 11,718 | 12,445 | 13,133 | 13,752 | 14,302 | 14,873 | 15,266 | 15,822 |
| 145,066 | 145,066 | 146,872 | 147,734 | 148,373 | 148,987 | 149,574 | 150,554 | 151,572 | 152,899 | 154,094 | 155,048 | 155,931 | 156,638 | 157,825 | 159,082 | 160,509 | 161,785 | 162,750 | 163,923 | 164,878 |
| 45,952 | 46,387 | 46,915 | 47,432 | 48,008 | 48,331 | 48,626 | 49,063 | 49,575 | 50,300 | 51,079 | 51,612 | 52,037 | 52,685 | 53,370 | 54,080 | 54,813 | 55,654 | 56,571 | 57,206 | 57,916 |
| 14,990 | 14,990 | 15,919 | 15,919 | 15,919 | 16,901 | 16,901 | 17,618 | 17,618 | 18,611 | 18,611 | 18,611 | 20,058 | 20,058 | 20,933 | 20,933 | 21,965 | 21,965 | 21,965 | 23,334 | 23,334 |
| 15,842 | 16,079 | 16,376 | 16,376 | 16,376 | 17,152 | 17,519 | 17,919 | 18,245 | 18,670 | 19,121 | 19,389 | 19,653 | 20,223 | 20,677 | 21,083 | 21,605 | 22,184 | 23,161 | 23,414 | 24,060 |
| 60,178 | 61,561 | 63,289 | 63,289 | 65,226 | 66,327 | 68,263 | 70,151 | 71,994 | 74,636 | 76,803 | 78,122 | 79,827 | 82,042 | 84,131 | 86,411 | 88,590 | 88,590 | 91,706 | 94,892 | 96,583 |
| 109,143 | 109,338 | 109,628 | 109,838 | 109,974 | 110,137 | 110,338 | 110,602 | 110,897 | 111,110 | 111,398 | 111,597 | 111,827 | 112,130 | 112,347 | 112,581 | 112,879 | 113,238 | 113,534 | 113,789 | 114,033 |
| 68,423 | 68,961 | 69,341 | 69,632 | 69,904 | 70,396 | 70,861 | 71,447 | 71,910 | 72,467 | 73,109 | 73,527 | 74,260 | 75,016 | 75,664 | 76,371 | 77,206 | 78,131 | 78,685 | 79,545 | 80,172 |
| 3,328 | 3,328 | 3,397 | 3,397 | 3,423 | 3,440 | 3,460 | 3,486 | 3,499 | 3,499 | 3,539 | 3,558 | 3,566 | 3,578 | 3,598 | 3,636 | 3,644 | 3,646 | 3,711 | 3,711 | 3,722 |
| 71,089 | 71,678 | 71,678 | 72,581 | 72,941 | 73,269 | 73,900 | 74,551 | 75,063 | 75,685 | 76,370 | 76,776 | 77,198 | 77,864 | 78,913 | 79,839 | 80,593 | 81,338 | 81,868 | 82,395 | 83,059 |
| 37,210 | 37,624 | 37,624 | 38,136 | 38,569 | 39,133 | 39,133 | 40,136 | 40,767 | 41,571 | 41,571 | 42,772 | 43,170 | 43,170 | 44,347 | 45,013 | 45,470 | 46,204 | 46,204 | 47,457 | 47,961 |
| 21,927 | 22,283 | 22,830 | 23,215 | 23,436 | 23,856 | 24,629 | 25,204 | 25,999 | 26,661 | 27,133 | 27,443 | 27,890 | 28,826 | 29,714 | 30,422 | 31,290 | 32,248 | 33,094 | 33,624 | 34,762 |
| 28,770 | 28,770 | 30,674 | 30,900 | 31,257 | 31,257 | 32,214 | 33,591 | 34,622 | 34,622 | 36,287 | 36,680 | 37,542 | 37,542 | 39,798 | 40,828 | 40,829 | 42,639 | 43,889 | 45,524 | 47,071 |
| 1,083 | 1,123 | 1,128 | 1,167 | 1,249 | 1,249 | 1,371 | 1,466 | 1,569 | 1,593 | 1,677 | 1,841 | 1,843 | 1,952 | 2,218 | 2,342 | 2,366 | 2,471 | 2,533 | 2,621 | 2,712 |
| 66,514 | 64,670 | 70,241 | 70,241 | 72,983 | 74,529 | 75,875 | 77,310 | 79,349 | 81,331 | 83,793 | 85,701 | 87,528 | 89,484 | 91,266 | 93,426 | 95,477 | 97,958 | 99,778 | 101,046 | 102,861 |
| 3,615 | 3,657 | 3,722 | 3,779 | 3,816 | 3,849 | 3,898 | 3,971 | 4,070 | 4,154 | 4,243 | 4,334 | 4,442 | 4,565 | 4,668 | 4,792 | 4,907 | 5,019 | 5,126 | 5,207 | 5,367 |
| 19,310 | 19,452 | 19,660 | 19,827 | 19,929 | 20,046 | 20,201 | 20,425 | 20,623 | 20,777 | 20,998 | 21,172 | 21,399 | 21,717 | 21,979 | 22,134 | 22,361 | 22,481 | 22,583 | 22,847 | 23,190 |
| 5,802 | 5,822 | 5,822 | 5,857 | 5,897 | 5,914 | 5,932 | 5,952 | 5,973 | 5,991 | 6,024 | 6,054 | 6,068 | 6,091 | 6,113 | 6,139 | 6,165 | 6,188 | 6,203 | 6,249 | 6,262 |
| 171,928 | 172,356 | 172,742 | 173,033 | 173,402 | 173,611 | 173,878 | 174,039 | 174,270 | 174,628 | 174,959 | 175,298 | 175,522 | 175,915 | 176,278 | 176,501 | 176,551 | 176,814 | 176,783 | 176,963 | 177,256 |
| 12,276 | 12,520 | 12,776 | 13,063 | 13,256 | 13,507 | 13,727 | 14,017 | 14,251 | 14,549 | 14,773 | 15,028 | 15,291 | 15,514 | 15,841 | 16,138 | 16,456 | 16,736 | 16,971 | 17,215 | 17,517 |
| 19,328 | 19,964 | 20,946 | 21,803 | 22,646 | 23,137 | 24,020 | 24,587 | 25,139 | 26,144 | 26,862 | 27,894 | 28,744 | 29,832 | 30,673 | 32,120 | 33,503 | 34,689 | 35,977 | 36,925 | 37,741 |
| 395982 | 396869 | 397675 | 398327 | 398828 | 399411 | 399925 | 400719 | 401345 | 402110 | 402861 | 403474 | 404051 | 405017 | 405876 | 406594 | 407334 | 407972 | 408495 | 408730 | 409845 |
| 52,865 | 54,166 | 55,257 | 56,183 | 57,151 | 57,956 | 58,904 | 60,181 | 61,331 | 62,856 | 64,214 | 65,592 | 66,853 | 67,995 | 69,311 | 70,601 | 72,280 | 73,822 | 74,932 | 76,168 | 77,215 |
| 13,853 | 14,217 | 14,217 | 15,645 | 15,776 | 16,063 | 16,501 | 17,357 | 18,036 | 18,680 | 19,779 | 19,934 | 20,401 | 20,922 | 21,832 | 22,907 | 23,535 | 25,056 | 25,265 | 25,581 | 26,326 |
| 8,931 | 9,294 | 9,636 | 9,930 | 10,230 | 10,395 | 10,605 | 10,819 | 11,188 | 11,454 | 11,851 | 12,170 | 12,438 | 12,805 | 13,081 | 13,509 | 13,802 | 14,149 | 14,579 | 14,847 | 15,139 |
| 88,074 | 88,741 | 89,375 | 89,854 | 90,304 | 91,299 | 92,148 | 92,867 | 93,876 | 94,689 | 95,266 | 95,742 | 96,671 | 97,665 | 98,446 | 99,478 | 100,241 | 101,027 | 101,738 | 102,765 | 103,396 |
| 16,853 | 16,941 | 16,991 | 16,991 | 16,991 | 16,991 | 17,154 | 17,204 | 17,243 | 17,312 | 17,312 | 17,312 | 17,487 | 17,487 | 17,640 | 17,711 | 17,793 | 17,793 | 17,793 | 17,904 | 17,986 |
| 37,919 | 39,701 | 41,532 | 43,386 | 44,847 | 46,380 | 47,352 | 48,909 | 50,691 | 52,419 | 54,699 | 56,648 | 58,168 | 60,389 | 62,245 | 64,083 | 66,060 | 67,612 | 69,986 | 71,445 | 73,337 |
| 6,826 | 6,893 | 6,978 | 7,028 | 7,063 | 7,105 | 7,163 | 7,242 | 7,336 | 7,401 | 7,454 | 7,499 | 7,524 | 7,572 | 7,652 | 7,694 | 7,789 | 7,862 | 7,906 | 7,943 | 8,019 |
| 45,315 | 46,890 | 48,712 | 50,140 | 51,431 | 52,155 | 53,514 | 55,986 | 57,591 | 59,546 | 61,006 | 61,960 | 65,274 | 66,788 | 69,061 | 71,540 | 73,819 | 76,336 | 78,115 | 79,754 | 81,944 |
| 168,062 | 175,977 | 183,532 | 191,790 | 195,239 | 200,557 | 210,585 | 220,564 | 230,346 | 240,111 | 250,462 | 258,658 | 264,313 | 275,058 | 282,365 | 292,656 | 307,572 | 317,730 | 325,030 | 332,434 | 341,739 |
| 23,386 | 23,921 | 24,571 | 25,002 | 25,500 | 26,004 | 26,706 | 27,480 | 28,054 | 28,963 | 29,605 | 30,177 | 30,722 | 31,196 | 31,664 | 32,587 | 33,392 | 34,045 | 34,682 | 35,084 | 35,648 |
| 63,203 | 63,735 | 64,393 | 65,748 | 66,102 | 66,740 | 67,375 | 67,988 | 68,931 | 69,782 | 70,670 | 71,642 | 72,443 | 73,527 | 74,431 | 75,433 | 76,373 | 77,430 | 78,375 | 79,371 | 80,393 |
| 1,210 | 1,227 | 1,238 | 1,238 | 1,251 | 1,251 | 1,254 | 1,256 | 1,272 | 1,283 | 1,283 | 1,296 | 1,301 | 1,305 | 1,318 | 1,325 | 1,334 | 1,338 | 1,350 | 1,360 | 1,366 |
| 33,435 | 34,151 | 34,778 | 35,247 | 35,898 | 36,985 | 37,420 | 37,941 | 38,581 | 39,218 | 39,218 | 40,656 | 41,757 | 42,304 | 43,046 | 44,313 | 45,067 | 46,026 | 46,946 | 47,743 | 48,575 |
| 32,225 | 32,809 | 33,431 | 34,207 | 34,740 | 35,230 | 35,765 | 36,410 | 37,210 | 38,099 | 39,080 | 39,877 | 40,382 | 41,349 | 42,197 | 43,139 | 44,068 | 45,099 | 45,948 | 46,675 | 47,836 |
| 2,979 | 3,053 | 3,126 | 3,205 | 3,335 | 3,442 | 3,505 | 3,707 | 3,826 | 3,983 | 4,146 | 4,244 | 4,313 | 4,407 | 4,557 | 4,657 | 4,783 | 4,922 | 5,042 | 5,142 | 5,199 |
| 1,514 | 1,550 | 1,582 | 1,606 | 1,634 | 1,675 | 1,711 | 1,740 | 1,774 | 1,790 | 1,839 | 1,862 | 1,903 | 1,951 | 1,985 | 2,026 | 2,069 | 2,108 | 2,126 | 2,187 | 2,238 |

| 7/22/2020 | 7/23/2020 | 7/24/2020 | 7/25/2020 | 7/26/2020 | 7/27/2020 | 7/28/2020 | 7/29/2020 | 7/30/2020 | 7/31/2020 | 8/1/2020 | 8/2/2020 | 8/3/2020 | 8/4/2020 | 8/5/2020 | 8/6/2020 | 8/7/2020 | 8/8/2020 | 8/9/2020 | 8/10/2020 | 8/11/2020 |
|---|---|---|---|---|---|---|---|---|---|---|---|---|---|---|---|---|---|---|---|---|
| 2,132 | 2,192 | 2,249 | 2,338 | 2,524 | 2,622 | 2,729 | 2,797 | 2,878 | 2,990 | 3,136 | 3,280 | 3,341 | 3,394 | 3,449 | 3,484 | 3,536 | 3,613 | 3,711 | 3,775 | 3,821 |
| 71,886 | 74,344 | 76,136 | 78,130 | 79,294 | 81,270 | 82,530 | 83,782 | 85,906 | 87,867 | 89,349 | 91,444 | 92,661 | 93,847 | 94,819 | 96,592 | 98,484 | 100,353 | 101,334 | 103,020 | 103,851 |
| 35,246 | 36,259 | 37,249 | 37,981 | 38,623 | 39,447 | 40,181 | 40,968 | 41,759 | 42,511 | 43,173 | 43,810 | 44,597 | 45,381 | 46,293 | 47,028 | 48,039 | 48,811 | 49,383 | 50,028 | 50,411 |
| 150,609 | 152,944 | 156,301 | 160,041 | 162,014 | 163,827 | 165,934 | 168,273 | 170,798 | 174,010 | 177,022 | 178,467 | 179,497 | 180,505 | 182,203 | 183,647 | 185,053 | 186,107 | 186,923 | 187,523 | 188,737 |
| 413,576 | 425,616 | 435,334 | 445,400 | 453,659 | 460,550 | 466,550 | 475,305 | 485,502 | 493,588 | 500,130 | 509,162 | 514,901 | 519,427 | 524,722 | 529,980 | 538,416 | 545,787 | 554,160 | 561,911 | 574,411 |
| 41,698 | 42,314 | 42,980 | 43,789 | 44,336 | 44,565 | 45,314 | 45,796 | 46,204 | 46,809 | 47,267 | 47,716 | 47,968 | 48,394 | 48,988 | 49,436 | 49,893 | 50,324 | 50,660 | 51,039 | 51,441 |
| 48,223 | 48,232 | 48,776 | 48,776 | 48,776 | 48,983 | 49,077 | 49,540 | 49,670 | 49,810 | 49,810 | 49,810 | 50,062 | 50,110 | 50,225 | 50,245 | 50,320 | 50,320 | 50,320 | 50,567 | 50,684 |
| 11,529 | 11,571 | 11,649 | 11,717 | 11,780 | 11,858 | 11,945 | 11,999 | 12,057 | 12,126 | 12,205 | 12,274 | 12,313 | 12,398 | 12,443 | 12,518 | 12,589 | 12,653 | 12,753 | 12,807 | 12,896 |
| 13,924 | 13,924 | 14,202 | 14,175 | 14,406 | 14,476 | 14,602 | 14,689 | 14,689 | 14,877 | 14,877 | 14,949 | 15,055 | 15,296 | 15,365 | 15,365 | 15,502 | 15,575 | 15,634 | 15,634 | 15,699 |
| 374,920 | 385,091 | 397,470 | 409,585 | 418,844 | 427,698 | 436,867 | 446,251 | 456,105 | 465,030 | 474,621 | 481,668 | 486,426 | 491,773 | 497,181 | 504,768 | 512,421 | 520,846 | 527,036 | 531,188 | 536,981 |
| 152,302 | 156,588 | 161,401 | 165,188 | 167,953 | 170,843 | 175,052 | 178,323 | 182,286 | 186,352 | 190,012 | 193,177 | 195,435 | 197,948 | 201,713 | 204,895 | 209,004 | 213,427 | 216,596 | 219,025 | 222,588 |
| 1,339 | 1,389 | 1,449 | 1,516 | 1,576 | 1,603 | 1,647 | 1,750 | 1,871 | 1,989 | 2,067 | 2,110 | 2,311 | 2,442 | 2,603 | 2,741 | 2,919 | 3,138 | 3,270 | 3,408 | 3,523 |
| 40,000 | 40,635 | 41,271 | 41,671 | 42,199 | 42,554 | 42,782 | 43,280 | 44,039 | 44,582 | 44,981 | 45,492 | 45,845 | 46,045 | 46,659 | 47,362 | 47,865 | 48,286 | 48,789 | 49,074 | 49,329 |
| 16,322 | 16,736 | 17,264 | 17,827 | 18,177 | 18,694 | 19,222 | 19,679 | 20,246 | 20,721 | 21,114 | 21,344 | 21,675 | 22,234 | 22,707 | 23,399 | 23,922 | 24,495 | 24,671 | 25,100 | 25,595 |
| 166,476 | 168,100 | 169,699 | 171,125 | 172,666 | 173,897 | 174,973 | 176,366 | 178,138 | 180,118 | 181,757 | 183,224 | 184,522 | 185,993 | 187,752 | 189,705 | 191,808 | 193,998 | 195,380 | 196,699 | 198,248 |
| 58,673 | 59,602 | 60,598 | 61,520 | 62,372 | 62,907 | 63,678 | 64,299 | 65,253 | 66,154 | 67,122 | 67,857 | 68,433 | 69,255 | 69,975 | 71,015 | 72,254 | 73,287 | 74,328 | 74,992 | 75,862 |
| 24,104 | 24,104 | 25,109 | 25,109 | 25,109 | 26,172 | 26,172 | 26,870 | 26,870 | 27,812 | 27,812 | 27,812 | 28,876 | 28,876 | 29,717 | 29,717 | 30,638 | 30,638 | 30,638 | 31,730 | 31,730 |
| 24,540 | 25,147 | 25,931 | 26,764 | 27,079 | 27,601 | 28,126 | 28,727 | 29,386 | 30,151 | 30,723 | 31,185 | 31,508 | 32,197 | 32,741 | 33,254 | 33,796 | 34,758 | 34,982 | 35,254 | 35,793 |
| 99,354 | 101,650 | 103,734 | 103,734 | 107,574 | 109,917 | 111,038 | 112,773 | 114,481 | 116,280 | 116,280 | 119,747 | 120,846 | 124,461 | 125,943 | 127,246 | 128,746 | 128,746 | 131,399 | 131,961 | 133,125 |
| 114,320 | 114,647 | 114,985 | 115,274 | 115,637 | 115,926 | 116,182 | 116,684 | 117,098 | 117,612 | 118,040 | 118,458 | 118,657 | 119,203 | 119,643 | 119,874 | 120,291 | 120,711 | 121,040 | 121,315 | 121,707 |
| 80,836 | 81,766 | 83,054 | 83,748 | 84,876 | 85,524 | 86,285 | 87,177 | 88,346 | 89,365 | 90,274 | 91,144 | 91,854 | 92,426 | 93,005 | 93,806 | 94,581 | 95,503 | 96,258 | 96,843 | 97,384 |
| 3,737 | 3,758 | 3,757 | 3,814 | 3,832 | 3,838 | 3,866 | 3,888 | 3,910 | 3,937 | 3,958 | 3,970 | 3,975 | 3,992 | 3,997 | 4,015 | 4,026 | 4,042 | 4,051 | 4,050 | 4,069 |
| 83,730 | 84,431 | 85,072 | 85,622 | 86,661 | 87,173 | 87,958 | 88,974 | 89,781 | 90,574 | 91,332 | 91,761 | 92,374 | 93,175 | 93,893 | 94,656 | 95,470 | 96,191 | 96,726 | 97,306 | 98,213 |
| 48,721 | 49,490 | 50,291 | 50,291 | 51,803 | 52,281 | 52,947 | 53,692 | 54,464 | 55,188 | 55,947 | 55,947 | 57,162 | 57,779 | 58,640 | 59,185 | 60,101 | 60,898 | 61,516 | 61,839 | 61,839 |
| 36,063 | 37,700 | 39,352 | 40,709 | 41,927 | 43,050 | 44,823 | 46,750 | 48,834 | 50,323 | 51,258 | 51,840 | 52,887 | 54,080 | 55,321 | 56,383 | 57,379 | 57,379 | 58,927 | 59,954 | 60,935 |
| 48,053 | 49,663 | 51,097 | 52,304 | 52,304 | 52,957 | 55,804 | 57,579 | 57,579 | 59,881 | 59,881 | 61,125 | 62,199 | 62,199 | 63,444 | 64,400 | 66,646 | 66,646 | 67,173 | 67,649 | 68,293 |
| 2,909 | 3,037 | 3,247 | 3,342 | 3,381 | 3,381 | 3,667 | 3,780 | 3,954 | 3,965 | 4,081 | 4,193 | 4,314 | 4,429 | 4,593 | 4,744 | 4,888 | 4,952 | 5,017 | 5,017 | 5,289 |
| 105,001 | 106,893 | 108,995 | 111,092 | 112,713 | 114,338 | 116,087 | 117,850 | 120,194 | 122,148 | 123,878 | 125,219 | 126,532 | 128,161 | 129,288 | 131,267 | 132,812 | 134,766 | 136,218 | 136,844 | 137,895 |
| 5,493 | 5,614 | 5,736 | 5,876 | 5,986 | 6,141 | 6,227 | 6,301 | 6,469 | 6,602 | 6,660 | 6,785 | 6,933 | 7,057 | 7,177 | 7,327 | 7,508 | 7,596 | 7,713 | 7,885 | 7,970 |
| 23,486 | 23,818 | 24,174 | 24,395 | 24,618 | 24,889 | 25,157 | 25,422 | 25,766 | 26,211 | 26,391 | 26,702 | 26,956 | 27,178 | 27,489 | 27,821 | 28,104 | 28,245 | 28,432 | 28,696 | 29,030 |
| 6,295 | 6,318 | 6,375 | 6,415 | 6,436 | 6,441 | 6,500 | 6,513 | 6,544 | 6,583 | 6,613 | 6,634 | 6,660 | 6,693 | 6,719 | 6,742 | 6,779 | 6,818 | 6,831 | 6,840 | 6,861 |
| 177,645 | 177,887 | 178,345 | 178,858 | 179,363 | 179,812 | 180,295 | 180,766 | 180,970 | 181,660 | 182,029 | 182,350 | 182,614 | 182,970 | 183,327 | 183,701 | 184,061 | 184,429 | 184,773 | 185,031 | 185,475 |
| 17,828 | 18,163 | 18,475 | 18,788 | 19,042 | 19,502 | 19,791 | 20,136 | 20,388 | 20,600 | 20,796 | 21,016 | 21,130 | 21,340 | 21,566 | 21,773 | 21,965 | 22,115 | 22,315 | 22,444 | 22,643 |
| 38,877 | 40,140 | 40,140 | 42,037 | 43,067 | 44,055 | 45,151 | 46,021 | 47,170 | 48,312 | 49,306 | 50,437 | 51,422 | 52,423 | 53,068 | 53,919 | 54,757 | 55,644 | 56,456 | 57,198 | 57,745 |
| 410560 | 411140 | 412062 | 412562 | 413091 | 413834 | 414479 | 415209 | 415960 | 416633 | 417273 | 417730 | 418352 | 419131 | 419752 | 420523 | 421137 | 421834 | 422349 | 422824 | 423587 |
| 78,742 | 80,186 | 81,746 | 83,184 | 84,073 | 85,177 | 86,497 | 87,893 | 89,626 | 91,159 | 92,087 | 93,031 | 93,963 | 95,106 | 96,305 | 97,471 | 98,675 | 99,969 | 100,848 | 101,731 | 102,826 |
| 27,541 | 28,221 | 29,348 | 30,360 | 33,582 | 34,981 | 34,990 | 37,060 | 38,291 | 39,105 | 40,418 | 40,910 | 41,199 | 41,423 | 43,414 | 44,325 | 45,272 | 46,159 | 46,652 | 47,067 | 47,908 |
| 15,394 | 15,713 | 16,104 | 16,492 | 16,758 | 17,088 | 17,416 | 17,721 | 18,131 | 18,493 | 18,817 | 19,097 | 19,366 | 19,699 | 19,978 | 20,225 | 20,636 | 21,010 | 21,272 | 21,488 | 21,774 |
| 104,358 | 105,571 | 106,625 | 107,425 | 108,264 | 109,384 | 110,218 | 111,078 | 112,048 | 112,936 | 112,936 | 113,590 | 114,155 | 115,009 | 115,714 | 116,521 | 117,279 | 118,092 | 118,852 | 119,453 | 120,281 |
| 18,062 | 18,148 | 18,224 | 18,224 | 18,224 | 18,515 | 18,725 | 18,800 | 18,950 | 19,022 | 19,022 | 19,022 | 19,246 | 19,390 | 19,481 | 19,611 | 19,738 | 19,738 | 19,738 | 19,934 | 20,053 |
| 75,042 | 76,606 | 78,607 | 80,008 | 81,199 | 82,417 | 84,109 | 85,846 | 87,572 | 89,016 | 90,599 | 91,788 | 92,951 | 94,190 | 95,472 | 96,797 | 98,219 | 99,460 | 100,435 | 101,159 | 102,130 |
| 8,077 | 8,143 | 8,200 | 8,305 | 8,395 | 8,444 | 8,492 | 8,641 | 8,685 | 8,764 | 8,867 | 8,955 | 9,020 | 9,079 | 9,168 | 9,273 | 9,371 | 9,477 | 9,605 | 9,663 | 9,713 |
| 84,417 | 86,987 | 89,078 | 90,796 | 93,936 | 96,489 | 99,044 | 100,822 | 102,871 | 105,959 | 108,184 | 109,627 | 110,636 | 112,441 | 114,098 | 116,350 | 118,782 | 120,585 | 122,712 | 123,914 | 124,915 |
| 351,618 | 361,125 | 369,826 | 375,846 | 381,656 | 385,923 | 394,265 | 403,307 | 412,107 | 420,946 | 430,485 | 430,485 | 442,014 | 451,181 | 459,887 | 467,485 | 474,524 | 481,483 | 486,362 | 490,817 | 500,620 |
| 36,254 | 36,889 | 37,681 | 38,064 | 38,502 | 38,915 | 39,337 | 39,942 | 40,258 | 40,797 | 41,380 | 41,759 | 42,153 | 42,544 | 43,096 | 43,606 | 44,067 | 44,090 | 44,714 | 45,006 | 45,399 |
| 81,237 | 82,364 | 83,609 | 84,567 | 86,072 | 86,994 | 87,993 | 88,904 | 89,888 | 90,801 | 91,782 | 93,106 | 94,251 | 95,049 | 95,867 | 97,882 | 99,189 | 100,086 | 100,750 | 101,746 | 102,521 |
| 1,366 | 1,377 | 1,385 | 1,396 | 1,400 | 1,402 | 1,405 | 1,406 | 1,407 | 1,414 | 1,421 | 1,426 | 1,426 | 1,431 | 1,436 | 1,445 | 1,448 | 1,459 | 1,462 | 1,462 | 1,478 |
| 49,247 | 50,009 | 50,824 | 51,849 | 52,635 | 53,321 | 54,205 | 54,995 | 55,803 | 55,803 | 57,541 | 58,172 | 58,715 | 59,379 | 60,084 | 60,917 | 61,587 | 62,523 | 63,072 | 63,647 | 64,151 |
| 48,583 | 49,669 | 50,727 | 51,715 | 52,680 | 53,281 | 54,064 | 54,988 | 56,079 | 56,934 | 58,058 | 58,990 | 59,401 | 60,171 | 61,110 | 61,985 | 63,028 | 64,213 | 64,835 | 65,356 | 66,123 |
| 5,461 | 5,550 | 5,695 | 5,821 | 5,960 | 6,054 | 6,173 | 6,326 | 6,422 | 6,642 | 6,735 | 6,854 | 6,973 | 7,051 | 7,159 | 7,277 | 7,433 | 7,563 | 7,694 | 7,754 | 7,875 |
| 2,288 | 2,347 | 2,405 | 2,446 | 2,475 | 2,520 | 2,589 | 2,628 | 2,686 | 2,726 | 2,769 | 2,808 | 2,848 | 2,884 | 2,923 | 2,958 | 3,000 | 3,013 | 3,050 | 3,042 | 3,073 |

| 8/12/2020 | 8/13/2020 | 8/14/2020 | 8/15/2020 | 8/16/2020 | 8/17/2020 | 8/18/2020 | 8/19/2020 | 8/20/2020 | 8/21/2020 | 8/22/2020 | 8/23/2020 |
|---|---|---|---|---|---|---|---|---|---|---|---|
| 3,881 | 3,963 | 4,073 | 4,156 | 4,259 | 4,309 | 4,371 | 4,438 | 4,520 | 4,588 | 4,677 | 4,741 |
| 104,786 | 105,557 | 106,309 | 107,580 | 108,433 | 109,004 | 110,361 | 111,478 | 112,449 | 113,632 | 114,532 | 115,992 |
| 51,114 | 51,766 | 52,392 | 51,992 | 52,665 | 53,077 | 53,487 | 54,216 | 54,765 | 55,652 | 56,199 | 56,574 |
| 189,443 | 190,794 | 191,721 | 192,654 | 193,537 | 194,005 | 194,920 | 195,557 | 196,280 | 196,899 | 197,895 | 198,103 |
| 586,056 | 593,141 | 601,075 | 613,689 | 621,562 | 628,031 | 632,667 | 638,831 | 644,751 | 650,336 | 656,892 | 663,669 |
| 51,756 | 52,129 | 52,538 | 52,838 | 53,176 | 53,370 | 53,631 | 53,901 | 54,230 | 54,586 | 54,883 | 55,133 |
| 50,706 | 50,782 | 50,897 | 50,897 | 50,897 | 51,267 | 51,255 | 51,314 | 51,432 | 51,519 | 51,519 | 51,519 |
| 12,959 | 13,024 | 13,118 | 13,159 | 13,220 | 13,273 | 13,325 | 13,354 | 13,409 | 13,469 | 13,534 | 13,590 |
| 15,967 | 15,967 | 16,396 | 16,451 | 16,536 | 16,536 | 16,643 | 16,718 | 16,770 | 16,831 | 16,895 | 16,942 |
| 545,040 | 551,232 | 557,337 | 563,628 | 567,375 | 570,024 | 573,811 | 577,891 | 582,407 | 587,023 | 591,283 | 594,287 |
| 226,153 | 228,668 | 231,895 | 235,168 | 237,030 | 238,861 | 241,677 | 243,982 | 246,741 | 249,630 | 252,222 | 253,949 |
| 3,699 | 4,022 | 4,241 | 4,510 | 4,697 | 4,839 | 4,965 | 5,176 | 5,372 | 5,587 | 5,864 | 6,111 |
| 49,806 | 50,336 | 50,935 | 51,793 | 52,433 | 52,829 | 52,969 | 53,280 | 54,533 | 55,006 | 55,735 | 56,275 |
| 26,133 | 26,631 | 27,173 | 27,477 | 27,660 | 27,942 | 28,326 | 28,696 | 29,120 | 29,369 | 29,662 | 29,853 |
| 199,893 | 201,727 | 204,023 | 205,851 | 207,413 | 209,186 | 210,926 | 213,221 | 215,053 | 217,346 | 219,702 | 221,595 |
| 76,522 | 77,565 | 78,632 | 79,676 | 80,415 | 81,006 | 81,847 | 82,336 | 83,277 | 84,317 | 85,317 | 85,932 |
| 32,547 | 32,547 | 33,885 | 33,885 | 33,885 | 35,167 | 35,167 | 35,890 | 35,890 | 36,856 | 36,856 | 36,856 |
| 36,945 | 37,686 | 38,298 | 38,930 | 39,315 | 39,691 | 40,299 | 40,926 | 41,626 | 42,265 | 43,066 | 43,529 |
| 134,304 | 135,439 | 136,737 | 136,737 | 137,918 | 138,485 | 139,125 | 139,903 | 140,821 | 141,720 | 141,720 | 142,943 |
| 121,936 | 122,255 | 124,081 | 123,199 | 123,593 | 123,842 | 124,063 | 124,415 | 124,728 | 125,159 | 125,268 | 125,268 |
| 98,160 | 98,875 | 99,693 | 100,212 | 100,715 | 101,235 | 101,649 | 102,229 | 102,899 | 103,523 | 104,102 | 104,669 |
| 4,089 | 4,115 | 4,144 | 4,168 | 4,197 | 4,213 | 4,235 | 4,253 | 4,285 | 4,316 | 4,335 | 4,356 |
| 98,689 | 99,856 | 100,724 | 101,782 | 102,259 | 102,749 | 103,403 | 104,091 | 104,618 | 104,618 | 106,044 | 106,808 |
| 62,296 | 63,723 | 64,413 | 65,162 | 65,716 | 65,716 | 66,061 | 67,308 | 68,133 | 68,133 | 69,584 | 70,298 |
| 62,530 | 63,797 | 65,270 | 66,397 | 67,475 | 68,623 | 69,417 | 70,675 | 71,733 | 72,964 | 74,257 | 75,075 |
| 69,374 | 69,986 | 70,930 | 71,755 | 72,136 | 73,207 | 73,207 | 75,449 | 75,449 | 77,268 | 77,894 | 78,405 |
| 5,403 | 5,539 | 5,541 | 5,659 | 5,779 | 5,847 | 5,847 | 5,956 | 6,214 | 6,216 | 6,376 | 6,490 |
| 139,061 | 140,824 | 142,170 | 143,706 | 144,952 | 145,516 | 146,779 | 147,932 | 149,904 | 151,912 | 153,641 | 155,113 |
| 8,171 | 8,322 | 8,444 | 8,587 | 8,647 | 8,782 | 8,968 | 9,242 | 9,474 | 9,736 | 9,876 | 10,000 |
| 29,244 | 29,660 | 29,988 | 30,241 | 30,372 | 30,563 | 30,825 | 31,040 | 31,348 | 31,626 | 31,780 | 31,889 |
| 6,887 | 6,921 | 6,964 | 6,980 | 6,988 | 7,004 | 7,017 | 7,036 | 7,050 | 7,071 | 7,092 | 7,107 |
| 185,938 | 186,594 | 187,164 | 187,442 | 187,455 | 187,767 | 188,098 | 188,427 | 188,527 | 188,817 | 189,236 | 189,494 |
| 22,816 | 22,987 | 23,160 | 23,302 | 23,408 | 23,500 | 23,579 | 23,749 | 23,951 | 24,095 | 24,302 | 24,396 |
| 58,270 | 58,856 | 59,955 | 60,814 | 61,511 | 62,171 | 62,854 | 63,248 | 63,804 | 64,697 | 65,334 | 65,866 |
| 424329 | 424955 | 425315 | 426386 | 426909 | 427270 | 427897 | 428558 | 429087 | 429727 | 430408 | 430885 |
| 104,248 | 105,426 | 106,557 | 107,674 | 108,287 | 109,062 | 109,923 | 110,881 | 112,003 | 113,046 | 114,165 | 114,802 |
| 48,700 | 49,505 | 50,384 | 51,387 | 51,387 | 52,468 | 53,264 | 54,132 | 54,993 | 56,079 | 57,113 | 57,686 |
| 22,022 | 22,300 | 22,613 | 23,018 | 23,744 | 23,451 | 23,676 | 23,870 | 24,165 | 24,421 | 24,710 | 24,937 |
| 121,130 | 122,121 | 122,950 | 123,800 | 124,460 | 124,844 | 125,579 | 126,149 | 126,940 | 127,633 | 128,429 | 129,048 |
| 20,129 | 20,240 | 20,355 | 20,355 | 20,335 | 20,572 | 20,692 | 20,795 | 20,871 | 21,022 | 21,022 | 21,022 |
| 102,974 | 103,909 | 104,841 | 105,882 | 106,497 | 106,953 | 107,672 | 108,411 | 109,320 | 110,378 | 111,295 | 111,988 |
| 9,815 | 9,897 | 10,024 | 10,118 | 10,274 | 10,360 | 10,443 | 10,566 | 10,691 | 10,884 | 11,135 | 11,276 |
| 126,393 | 128,511 | 130,458 | 131,747 | 133,708 | 134,744 | 135,778 | 137,800 | 139,175 | 140,844 | 142,083 | 143,937 |
| 506,820 | 513,575 | 520,593 | 528,838 | 535,042 | 542,950 | 550,232 | 557,256 | 562,559 | 567,580 | 573,139 | 577,537 |
| 45,722 | 46,155 | 46,628 | 46,942 | 47,248 | 47,487 | 47,785 | 48,260 | 48,761 | 49,088 | 49,416 | 49,728 |
| 103,623 | 104,838 | 105,753 | 106,691 | 107,421 | 108,283 | 109,019 | 109,882 | 110,860 | 112,072 | 112,072 | 113,630 |
| 1,478 | 1,484 | 1,501 | 1,509 | 1,515 | 1,527 | 1,530 | 1,533 | 1,537 | 1,541 | 1,553 | 1,557 |
| 64,702 | 65,339 | 66,139 | 66,885 | 67,461 | 67,721 | 68,264 | 68,689 | 69,389 | 69,779 | 70,595 | 71,012 |
| 66,654 | 67,622 | 68,681 | 69,547 | 70,246 | 70,715 | 71,424 | 72,134 | 72,893 | 73,751 | 74,726 | 75,205 |
| 8,008 | 8,151 | 8,274 | 8,457 | 8,564 | 8,632 | 8,731 | 8,801 | 8,982 | 9,066 | 9,185 | 9,272 |
| 3,086 | 3,119 | 3,183 | 3,227 | 3,286 | 3,331 | 3,363 | 3,430 | 3,468 | 3,524 | 3,543 | 3,579 |

**eTable 4. COVID-19 death**

| Date/Total Death | 1/22/2020 | 1/23/2020 | 1/24/2020 | 1/25/2020 | 1/26/2020 | 1/27/2020 | 1/28/2020 | 1/29/2020 | 1/30/2020 | 1/31/2020 | 2/1/2020 | 2/2/2020 | 2/3/2020 | 2/4/2020 | 2/5/2020 | 2/6/2020 | 2/7/2020 | 2/8/2020 | 2/9/2020 | 2/10/2020 |
|---|---|---|---|---|---|---|---|---|---|---|---|---|---|---|---|---|---|---|---|---|
| AK | 0 | 0 | 0 | 0 | 0 | 0 | 0 | 0 | 0 | 0 | 0 | 0 | 0 | 0 | 0 | 0 | 0 | 0 | 0 | 0 |
| AL | 0 | 0 | 0 | 0 | 0 | 0 | 0 | 0 | 0 | 0 | 0 | 0 | 0 | 0 | 0 | 0 | 0 | 0 | 0 | 0 |
| AR | 0 | 0 | 0 | 0 | 0 | 0 | 0 | 0 | 0 | 0 | 0 | 0 | 0 | 0 | 0 | 0 | 0 | 0 | 0 | 0 |
| AZ | 0 | 0 | 0 | 0 | 0 | 0 | 0 | 0 | 0 | 0 | 0 | 0 | 0 | 0 | 0 | 0 | 0 | 0 | 0 | 0 |
| CA | 0 | 0 | 0 | 0 | 0 | 0 | 0 | 0 | 0 | 0 | 0 | 0 | 0 | 0 | 0 | 0 | 0 | 0 | 0 | 0 |
| CO | 0 | 0 | 0 | 0 | 0 | 0 | 0 | 0 | 0 | 0 | 0 | 0 | 0 | 0 | 0 | 0 | 0 | 0 | 0 | 0 |
| CT | 0 | 0 | 0 | 0 | 0 | 0 | 0 | 0 | 0 | 0 | 0 | 0 | 0 | 0 | 0 | 0 | 0 | 0 | 0 | 0 |
| DC | 0 | 0 | 0 | 0 | 0 | 0 | 0 | 0 | 0 | 0 | 0 | 0 | 0 | 0 | 0 | 0 | 0 | 0 | 0 | 0 |
| DE | 0 | 0 | 0 | 0 | 0 | 0 | 0 | 0 | 0 | 0 | 0 | 0 | 0 | 0 | 0 | 0 | 0 | 0 | 0 | 0 |
| FL | 0 | 0 | 0 | 0 | 0 | 0 | 0 | 0 | 0 | 0 | 0 | 0 | 0 | 0 | 0 | 0 | 0 | 0 | 0 | 0 |
| GA | 0 | 0 | 0 | 0 | 0 | 0 | 0 | 0 | 0 | 0 | 0 | 0 | 0 | 0 | 0 | 0 | 0 | 0 | 0 | 0 |
| HI | 0 | 0 | 0 | 0 | 0 | 0 | 0 | 0 | 0 | 0 | 0 | 0 | 0 | 0 | 0 | 0 | 0 | 0 | 0 | 0 |
| IA | 0 | 0 | 0 | 0 | 0 | 0 | 0 | 0 | 0 | 0 | 0 | 0 | 0 | 0 | 0 | 0 | 0 | 0 | 0 | 0 |
| ID | 0 | 0 | 0 | 0 | 0 | 0 | 0 | 0 | 0 | 0 | 0 | 0 | 0 | 0 | 0 | 0 | 0 | 0 | 0 | 0 |
| IL | 0 | 0 | 0 | 0 | 0 | 0 | 0 | 0 | 0 | 0 | 0 | 0 | 0 | 0 | 0 | 0 | 0 | 0 | 0 | 0 |
| IN | 0 | 0 | 0 | 0 | 0 | 0 | 0 | 0 | 0 | 0 | 0 | 0 | 0 | 0 | 0 | 0 | 0 | 0 | 0 | 0 |
| KS | 0 | 0 | 0 | 0 | 0 | 0 | 0 | 0 | 0 | 0 | 0 | 0 | 0 | 0 | 0 | 0 | 0 | 0 | 0 | 0 |
| KY | 0 | 0 | 0 | 0 | 0 | 0 | 0 | 0 | 0 | 0 | 0 | 0 | 0 | 0 | 0 | 0 | 0 | 0 | 0 | 0 |
| LA | 0 | 0 | 0 | 0 | 0 | 0 | 0 | 0 | 0 | 0 | 0 | 0 | 0 | 0 | 0 | 0 | 0 | 0 | 0 | 0 |
| MA | 0 | 0 | 0 | 0 | 0 | 0 | 0 | 0 | 0 | 0 | 0 | 0 | 0 | 0 | 0 | 0 | 0 | 0 | 0 | 0 |
| MD | 0 | 0 | 0 | 0 | 0 | 0 | 0 | 0 | 0 | 0 | 0 | 0 | 0 | 0 | 0 | 0 | 0 | 0 | 0 | 0 |
| ME | 0 | 0 | 0 | 0 | 0 | 0 | 0 | 0 | 0 | 0 | 0 | 0 | 0 | 0 | 0 | 0 | 0 | 0 | 0 | 0 |
| MI | 0 | 0 | 0 | 0 | 0 | 0 | 0 | 0 | 0 | 0 | 0 | 0 | 0 | 0 | 0 | 0 | 0 | 0 | 0 | 0 |
| MN | 0 | 0 | 0 | 0 | 0 | 0 | 0 | 0 | 0 | 0 | 0 | 0 | 0 | 0 | 0 | 0 | 0 | 0 | 0 | 0 |
| MO | 0 | 0 | 0 | 0 | 0 | 0 | 0 | 0 | 0 | 0 | 0 | 0 | 0 | 0 | 0 | 0 | 0 | 0 | 0 | 0 |
| MS | 0 | 0 | 0 | 0 | 0 | 0 | 0 | 0 | 0 | 0 | 0 | 0 | 0 | 0 | 0 | 0 | 0 | 0 | 0 | 0 |
| MT | 0 | 0 | 0 | 0 | 0 | 0 | 0 | 0 | 0 | 0 | 0 | 0 | 0 | 0 | 0 | 0 | 0 | 0 | 0 | 0 |
| NC | 0 | 0 | 0 | 0 | 0 | 0 | 0 | 0 | 0 | 0 | 0 | 0 | 0 | 0 | 0 | 0 | 0 | 0 | 0 | 0 |
| ND | 0 | 0 | 0 | 0 | 0 | 0 | 0 | 0 | 0 | 0 | 0 | 0 | 0 | 0 | 0 | 0 | 0 | 0 | 0 | 0 |
| NE | 0 | 0 | 0 | 0 | 0 | 0 | 0 | 0 | 0 | 0 | 0 | 0 | 0 | 0 | 0 | 0 | 0 | 0 | 0 | 0 |
| NH | 0 | 0 | 0 | 0 | 0 | 0 | 0 | 0 | 0 | 0 | 0 | 0 | 0 | 0 | 0 | 0 | 0 | 0 | 0 | 0 |
| NJ | 0 | 0 | 0 | 0 | 0 | 0 | 0 | 0 | 0 | 0 | 0 | 0 | 0 | 0 | 0 | 0 | 0 | 0 | 0 | 0 |
| NM | 0 | 0 | 0 | 0 | 0 | 0 | 0 | 0 | 0 | 0 | 0 | 0 | 0 | 0 | 0 | 0 | 0 | 0 | 0 | 0 |
| NV | 0 | 0 | 0 | 0 | 0 | 0 | 0 | 0 | 0 | 0 | 0 | 0 | 0 | 0 | 0 | 0 | 0 | 0 | 0 | 0 |
| NY | 0 | 0 | 0 | 0 | 0 | 0 | 0 | 0 | 0 | 0 | 0 | 0 | 0 | 0 | 0 | 0 | 0 | 0 | 0 | 0 |
| OH | 0 | 0 | 0 | 0 | 0 | 0 | 0 | 0 | 0 | 0 | 0 | 0 | 0 | 0 | 0 | 0 | 0 | 0 | 0 | 0 |
| OK | 0 | 0 | 0 | 0 | 0 | 0 | 0 | 0 | 0 | 0 | 0 | 0 | 0 | 0 | 0 | 0 | 0 | 0 | 0 | 0 |
| OR | 0 | 0 | 0 | 0 | 0 | 0 | 0 | 0 | 0 | 0 | 0 | 0 | 0 | 0 | 0 | 0 | 0 | 0 | 0 | 0 |
| PA | 0 | 0 | 0 | 0 | 0 | 0 | 0 | 0 | 0 | 0 | 0 | 0 | 0 | 0 | 0 | 0 | 0 | 0 | 0 | 0 |
| RI | 0 | 0 | 0 | 0 | 0 | 0 | 0 | 0 | 0 | 0 | 0 | 0 | 0 | 0 | 0 | 0 | 0 | 0 | 0 | 0 |
| SC | 0 | 0 | 0 | 0 | 0 | 0 | 0 | 0 | 0 | 0 | 0 | 0 | 0 | 0 | 0 | 0 | 0 | 0 | 0 | 0 |
| SD | 0 | 0 | 0 | 0 | 0 | 0 | 0 | 0 | 0 | 0 | 0 | 0 | 0 | 0 | 0 | 0 | 0 | 0 | 0 | 0 |
| TN | 0 | 0 | 0 | 0 | 0 | 0 | 0 | 0 | 0 | 0 | 0 | 0 | 0 | 0 | 0 | 0 | 0 | 0 | 0 | 0 |
| TX | 0 | 0 | 0 | 0 | 0 | 0 | 0 | 0 | 0 | 0 | 0 | 0 | 0 | 0 | 0 | 0 | 0 | 0 | 0 | 0 |
| UT | 0 | 0 | 0 | 0 | 0 | 0 | 0 | 0 | 0 | 0 | 0 | 0 | 0 | 0 | 0 | 0 | 0 | 0 | 0 | 0 |
| VA | 0 | 0 | 0 | 0 | 0 | 0 | 0 | 0 | 0 | 0 | 0 | 0 | 0 | 0 | 0 | 0 | 0 | 0 | 0 | 0 |
| VT | 0 | 0 | 0 | 0 | 0 | 0 | 0 | 0 | 0 | 0 | 0 | 0 | 0 | 0 | 0 | 0 | 0 | 0 | 0 | 0 |
| WA | 0 | 0 | 0 | 0 | 0 | 0 | 0 | 0 | 0 | 0 | 0 | 0 | 0 | 0 | 0 | 0 | 0 | 0 | 0 | 0 |
| WI | 0 | 0 | 0 | 0 | 0 | 0 | 0 | 0 | 0 | 0 | 0 | 0 | 0 | 0 | 0 | 0 | 0 | 0 | 0 | 0 |
| WV | 0 | 0 | 0 | 0 | 0 | 0 | 0 | 0 | 0 | 0 | 0 | 0 | 0 | 0 | 0 | 0 | 0 | 0 | 0 | 0 |
| WY | 0 | 0 | 0 | 0 | 0 | 0 | 0 | 0 | 0 | 0 | 0 | 0 | 0 | 0 | 0 | 0 | 0 | 0 | 0 | 0 |

| 2/11/2020 | 2/12/2020 | 2/13/2020 | 2/14/2020 | 2/15/2020 | 2/16/2020 | 2/17/2020 | 2/18/2020 | 2/19/2020 | 2/20/2020 | 2/21/2020 | 2/22/2020 | 2/23/2020 | 2/24/2020 | 2/25/2020 | 2/26/2020 | 2/27/2020 | 2/28/2020 | 2/29/2020 | 3/1/2020 |
|---|---|---|---|---|---|---|---|---|---|---|---|---|---|---|---|---|---|---|---|
| 0 | 0 | 0 | 0 | 0 | 0 | 0 | 0 | 0 | 0 | 0 | 0 | 0 | 0 | 0 | 0 | 0 | 0 | 0 | 0 |
| 0 | 0 | 0 | 0 | 0 | 0 | 0 | 0 | 0 | 0 | 0 | 0 | 0 | 0 | 0 | 0 | 0 | 0 | 0 | 0 |
| 0 | 0 | 0 | 0 | 0 | 0 | 0 | 0 | 0 | 0 | 0 | 0 | 0 | 0 | 0 | 0 | 0 | 0 | 0 | 0 |
| 0 | 0 | 0 | 0 | 0 | 0 | 0 | 0 | 0 | 0 | 0 | 0 | 0 | 0 | 0 | 0 | 0 | 0 | 0 | 0 |
| 0 | 0 | 0 | 0 | 0 | 0 | 0 | 0 | 0 | 0 | 0 | 0 | 0 | 0 | 0 | 0 | 0 | 0 | 0 | 0 |
| 0 | 0 | 0 | 0 | 0 | 0 | 0 | 0 | 0 | 0 | 0 | 0 | 0 | 0 | 0 | 0 | 0 | 0 | 0 | 0 |
| 0 | 0 | 0 | 0 | 0 | 0 | 0 | 0 | 0 | 0 | 0 | 0 | 0 | 0 | 0 | 0 | 0 | 0 | 0 | 0 |
| 0 | 0 | 0 | 0 | 0 | 0 | 0 | 0 | 0 | 0 | 0 | 0 | 0 | 0 | 0 | 0 | 0 | 0 | 0 | 0 |
| 0 | 0 | 0 | 0 | 0 | 0 | 0 | 0 | 0 | 0 | 0 | 0 | 0 | 0 | 0 | 0 | 0 | 0 | 0 | 0 |
| 0 | 0 | 0 | 0 | 0 | 0 | 0 | 0 | 0 | 0 | 0 | 0 | 0 | 0 | 0 | 0 | 0 | 0 | 0 | 0 |
| 0 | 0 | 0 | 0 | 0 | 0 | 0 | 0 | 0 | 0 | 0 | 0 | 0 | 0 | 0 | 0 | 0 | 0 | 0 | 0 |
| 0 | 0 | 0 | 0 | 0 | 0 | 0 | 0 | 0 | 0 | 0 | 0 | 0 | 0 | 0 | 0 | 0 | 0 | 0 | 0 |
| 0 | 0 | 0 | 0 | 0 | 0 | 0 | 0 | 0 | 0 | 0 | 0 | 0 | 0 | 0 | 0 | 0 | 0 | 0 | 0 |
| 0 | 0 | 0 | 0 | 0 | 0 | 0 | 0 | 0 | 0 | 0 | 0 | 0 | 0 | 0 | 0 | 0 | 0 | 0 | 0 |
| 0 | 0 | 0 | 0 | 0 | 0 | 0 | 0 | 0 | 0 | 0 | 0 | 0 | 0 | 0 | 0 | 0 | 0 | 0 | 0 |
| 0 | 0 | 0 | 0 | 0 | 0 | 0 | 0 | 0 | 0 | 0 | 0 | 0 | 0 | 0 | 0 | 0 | 0 | 0 | 0 |
| 0 | 0 | 0 | 0 | 0 | 0 | 0 | 0 | 0 | 0 | 0 | 0 | 0 | 0 | 0 | 0 | 0 | 0 | 0 | 0 |
| 0 | 0 | 0 | 0 | 0 | 0 | 0 | 0 | 0 | 0 | 0 | 0 | 0 | 0 | 0 | 0 | 0 | 0 | 0 | 0 |
| 0 | 0 | 0 | 0 | 0 | 0 | 0 | 0 | 0 | 0 | 0 | 0 | 0 | 0 | 0 | 0 | 0 | 0 | 0 | 0 |
| 0 | 0 | 0 | 0 | 0 | 0 | 0 | 0 | 0 | 0 | 0 | 0 | 0 | 0 | 0 | 0 | 0 | 0 | 0 | 0 |
| 0 | 0 | 0 | 0 | 0 | 0 | 0 | 0 | 0 | 0 | 0 | 0 | 0 | 0 | 0 | 0 | 0 | 0 | 0 | 0 |
| 0 | 0 | 0 | 0 | 0 | 0 | 0 | 0 | 0 | 0 | 0 | 0 | 0 | 0 | 0 | 0 | 0 | 0 | 0 | 0 |
| 0 | 0 | 0 | 0 | 0 | 0 | 0 | 0 | 0 | 0 | 0 | 0 | 0 | 0 | 0 | 0 | 0 | 0 | 0 | 0 |
| 0 | 0 | 0 | 0 | 0 | 0 | 0 | 0 | 0 | 0 | 0 | 0 | 0 | 0 | 0 | 0 | 0 | 0 | 0 | 0 |
| 0 | 0 | 0 | 0 | 0 | 0 | 0 | 0 | 0 | 0 | 0 | 0 | 0 | 0 | 0 | 0 | 0 | 0 | 0 | 0 |
| 0 | 0 | 0 | 0 | 0 | 0 | 0 | 0 | 0 | 0 | 0 | 0 | 0 | 0 | 0 | 0 | 0 | 0 | 0 | 0 |
| 0 | 0 | 0 | 0 | 0 | 0 | 0 | 0 | 0 | 0 | 0 | 0 | 0 | 0 | 0 | 0 | 0 | 0 | 0 | 0 |
| 0 | 0 | 0 | 0 | 0 | 0 | 0 | 0 | 0 | 0 | 0 | 0 | 0 | 0 | 0 | 0 | 0 | 0 | 0 | 0 |
| 0 | 0 | 0 | 0 | 0 | 0 | 0 | 0 | 0 | 0 | 0 | 0 | 0 | 0 | 0 | 0 | 0 | 0 | 0 | 0 |
| 0 | 0 | 0 | 0 | 0 | 0 | 0 | 0 | 0 | 0 | 0 | 0 | 0 | 0 | 0 | 0 | 0 | 0 | 0 | 0 |
| 0 | 0 | 0 | 0 | 0 | 0 | 0 | 0 | 0 | 0 | 0 | 0 | 0 | 0 | 0 | 0 | 0 | 0 | 0 | 0 |
| 0 | 0 | 0 | 0 | 0 | 0 | 0 | 0 | 0 | 0 | 0 | 0 | 0 | 0 | 0 | 0 | 0 | 0 | 0 | 0 |
| 0 | 0 | 0 | 0 | 0 | 0 | 0 | 0 | 0 | 0 | 0 | 0 | 0 | 0 | 0 | 0 | 0 | 0 | 0 | 0 |
| 0 | 0 | 0 | 0 | 0 | 0 | 0 | 0 | 0 | 0 | 0 | 0 | 0 | 0 | 0 | 0 | 0 | 0 | 0 | 0 |
| 0 | 0 | 0 | 0 | 0 | 0 | 0 | 0 | 0 | 0 | 0 | 0 | 0 | 0 | 0 | 0 | 0 | 0 | 0 | 0 |
| 0 | 0 | 0 | 0 | 0 | 0 | 0 | 0 | 0 | 0 | 0 | 0 | 0 | 0 | 0 | 0 | 0 | 0 | 0 | 0 |
| 0 | 0 | 0 | 0 | 0 | 0 | 0 | 0 | 0 | 0 | 0 | 0 | 0 | 0 | 0 | 0 | 0 | 0 | 0 | 0 |
| 0 | 0 | 0 | 0 | 0 | 0 | 0 | 0 | 0 | 0 | 0 | 0 | 0 | 0 | 0 | 0 | 0 | 0 | 0 | 0 |
| 0 | 0 | 0 | 0 | 0 | 0 | 0 | 0 | 0 | 0 | 0 | 0 | 0 | 0 | 0 | 0 | 0 | 0 | 0 | 0 |
| 0 | 0 | 0 | 0 | 0 | 0 | 0 | 0 | 0 | 0 | 0 | 0 | 0 | 0 | 0 | 0 | 0 | 0 | 0 | 0 |
| 0 | 0 | 0 | 0 | 0 | 0 | 0 | 0 | 0 | 0 | 0 | 0 | 0 | 0 | 0 | 0 | 0 | 0 | 0 | 0 |
| 0 | 0 | 0 | 0 | 0 | 0 | 0 | 0 | 0 | 0 | 0 | 0 | 0 | 0 | 0 | 0 | 0 | 0 | 0 | 0 |
| 0 | 0 | 0 | 0 | 0 | 0 | 0 | 0 | 0 | 0 | 0 | 0 | 0 | 0 | 0 | 0 | 0 | 0 | 0 | 0 |
| 0 | 0 | 0 | 0 | 0 | 0 | 0 | 0 | 0 | 0 | 0 | 0 | 0 | 0 | 0 | 0 | 0 | 0 | 0 | 0 |
| 0 | 0 | 0 | 0 | 0 | 0 | 0 | 0 | 0 | 0 | 0 | 0 | 0 | 0 | 0 | 0 | 0 | 0 | 0 | 0 |
| 0 | 0 | 0 | 0 | 0 | 0 | 0 | 0 | 0 | 0 | 0 | 0 | 0 | 0 | 0 | 0 | 0 | 0 | 0 | 0 |
| 0 | 0 | 0 | 0 | 0 | 0 | 0 | 0 | 0 | 0 | 0 | 0 | 0 | 0 | 0 | 0 | 0 | 0 | 0 | 0 |
| 0 | 0 | 0 | 0 | 0 | 0 | 0 | 0 | 0 | 0 | 0 | 0 | 0 | 0 | 0 | 0 | 0 | 0 | 1 | 1 |
| 0 | 0 | 0 | 0 | 0 | 0 | 0 | 0 | 0 | 0 | 0 | 0 | 0 | 0 | 0 | 0 | 0 | 0 | 0 | 0 |
| 0 | 0 | 0 | 0 | 0 | 0 | 0 | 0 | 0 | 0 | 0 | 0 | 0 | 0 | 0 | 0 | 0 | 0 | 0 | 0 |
| 0 | 0 | 0 | 0 | 0 | 0 | 0 | 0 | 0 | 0 | 0 | 0 | 0 | 0 | 0 | 0 | 0 | 0 | 0 | 0 |

| 3/2/2020 | 3/3/2020 | 3/4/2020 | 3/5/2020 | 3/6/2020 | 3/7/2020 | 3/8/2020 | 3/9/2020 | 3/10/2020 | 3/11/2020 | 3/12/2020 | 3/13/2020 | 3/14/2020 | 3/15/2020 | 3/16/2020 | 3/17/2020 | 3/18/2020 | 3/19/2020 | 3/20/2020 | 3/21/2020 | 3/22/2020 |
|---|---|---|---|---|---|---|---|---|---|---|---|---|---|---|---|---|---|---|---|---|
| 0 | 0 | 0 | 0 | 0 | 0 | 0 | 0 | 0 | 0 | 0 | 0 | 0 | 0 | 0 | 0 | 0 | 0 | 0 | 0 | 0 |
| 0 | 0 | 0 | 0 | 0 | 0 | 0 | 0 | 0 | 0 | 0 | 0 | 0 | 0 | 0 | 0 | 0 | 0 | 0 | 0 | 0 |
| 0 | 0 | 0 | 0 | 0 | 0 | 0 | 0 | 0 | 0 | 0 | 0 | 0 | 0 | 0 | 0 | 0 | 0 | 0 | 0 | 0 |
| 0 | 0 | 0 | 0 | 0 | 0 | 0 | 0 | 0 | 0 | 0 | 0 | 0 | 0 | 0 | 0 | 0 | 0 | 1 | 1 | 2 |
| 0 | 0 | 0 | 1 | 1 | 1 | 1 | 2 | 2 | 3 | 4 | 4 | 4 | 5 | 7 | 10 | 14 | 18 | 24 | 27 | 32 |
| 0 | 0 | 0 | 0 | 0 | 0 | 0 | 0 | 0 | 0 | 0 | 0 | 0 | 1 | 1 | 2 | 2 | 2 | 4 | 5 | 6 |
| 0 | 0 | 0 | 0 | 0 | 0 | 0 | 0 | 0 | 0 | 0 | 0 | 0 | 0 | 0 | 0 | 1 | 3 | 4 | 5 | 8 |
| 0 | 0 | 0 | 0 | 0 | 0 | 0 | 0 | 0 | 0 | 0 | 0 | 0 | 0 | 0 | 0 | 0 | 0 | 1 | 1 | 2 |
| 0 | 0 | 0 | 0 | 0 | 0 | 0 | 0 | 0 | 0 | 0 | 0 | 0 | 0 | 0 | 0 | 0 | 0 | 0 | 0 | 0 |
| 0 | 0 | 0 | 0 | 0 | 0 | 0 | 2 | 2 | 2 | 2 | 2 | 3 | 4 | 5 | 7 | 8 | 9 | 11 | 12 | 13 |
| 0 | 0 | 0 | 0 | 0 | 0 | 0 | 0 | 0 | 0 | 1 | 1 | 1 | 1 | 1 | 1 | 6 | 13 | 14 | 20 | 25 |
| 0 | 0 | 0 | 0 | 0 | 0 | 0 | 0 | 0 | 0 | 0 | 0 | 0 | 0 | 0 | 0 | 0 | 0 | 0 | 0 | 0 |
| 0 | 0 | 0 | 0 | 0 | 0 | 0 | 0 | 0 | 0 | 0 | 0 | 0 | 0 | 0 | 0 | 0 | 0 | 0 | 0 | 0 |
| 0 | 0 | 0 | 0 | 0 | 0 | 0 | 0 | 0 | 0 | 0 | 0 | 0 | 0 | 0 | 0 | 0 | 0 | 0 | 0 | 0 |
| 0 | 0 | 0 | 0 | 0 | 0 | 0 | 0 | 0 | 0 | 0 | 0 | 0 | 0 | 0 | 1 | 1 | 4 | 5 | 6 | 9 |
| 0 | 0 | 0 | 0 | 0 | 0 | 0 | 0 | 0 | 0 | 0 | 0 | 0 | 0 | 1 | 2 | 2 | 2 | 3 | 4 | 6 |
| 0 | 0 | 0 | 0 | 0 | 0 | 0 | 0 | 0 | 0 | 1 | 1 | 1 | 1 | 1 | 1 | 1 | 1 | 1 | 2 | 2 |
| 0 | 0 | 0 | 0 | 0 | 0 | 0 | 0 | 0 | 0 | 0 | 0 | 0 | 0 | 0 | 0 | 1 | 2 | 2 | 3 | 3 |
| 0 | 0 | 0 | 0 | 0 | 0 | 0 | 0 | 0 | 0 | 0 | 0 | 0 | 2 | 3 | 4 | 7 | 10 | 14 | 20 | 29 |
| 0 | 0 | 0 | 0 | 0 | 0 | 0 | 0 | 0 | 0 | 0 | 0 | 0 | 0 | 0 | 0 | 0 | 0 | 1 | 1 | 5 |
| 0 | 0 | 0 | 0 | 0 | 0 | 0 | 0 | 0 | 0 | 0 | 0 | 0 | 0 | 0 | 0 | 1 | 1 | 2 | 3 | 3 |
| 0 | 0 | 0 | 0 | 0 | 0 | 0 | 0 | 0 | 0 | 0 | 0 | 0 | 0 | 0 | 0 | 0 | 0 | 0 | 0 | 0 |
| 0 | 0 | 0 | 0 | 0 | 0 | 0 | 0 | 0 | 0 | 0 | 0 | 0 | 0 | 0 | 0 | 1 | 1 | 3 | 5 | 8 |
| 0 | 0 | 0 | 0 | 0 | 0 | 0 | 0 | 0 | 0 | 0 | 0 | 0 | 0 | 0 | 0 | 0 | 0 | 0 | 1 | 1 |
| 0 | 0 | 0 | 0 | 0 | 0 | 0 | 0 | 0 | 0 | 0 | 0 | 0 | 0 | 0 | 0 | 1 | 1 | 3 | 3 | 3 |
| 0 | 0 | 0 | 0 | 0 | 0 | 0 | 0 | 0 | 0 | 0 | 0 | 0 | 0 | 0 | 0 | 0 | 1 | 1 | 1 | 1 |
| 0 | 0 | 0 | 0 | 0 | 0 | 0 | 0 | 0 | 0 | 0 | 0 | 0 | 0 | 0 | 0 | 0 | 0 | 0 | 0 | 0 |
| 0 | 0 | 0 | 0 | 0 | 0 | 0 | 0 | 0 | 0 | 0 | 0 | 0 | 0 | 0 | 0 | 0 | 0 | 0 | 0 | 0 |
| 0 | 0 | 0 | 0 | 0 | 0 | 0 | 0 | 0 | 0 | 0 | 0 | 0 | 0 | 0 | 0 | 0 | 0 | 0 | 0 | 0 |
| 0 | 0 | 0 | 0 | 0 | 0 | 0 | 0 | 0 | 0 | 0 | 0 | 0 | 0 | 0 | 0 | 0 | 0 | 0 | 0 | 0 |
| 0 | 0 | 0 | 0 | 0 | 0 | 0 | 0 | 0 | 0 | 0 | 0 | 0 | 0 | 0 | 0 | 0 | 0 | 0 | 0 | 0 |
| 0 | 0 | 0 | 0 | 0 | 0 | 0 | 0 | 1 | 1 | 1 | 1 | 1 | 2 | 2 | 3 | 5 | 9 | 11 | 16 | 20 |
| 0 | 0 | 0 | 0 | 0 | 0 | 0 | 0 | 0 | 0 | 0 | 0 | 0 | 0 | 0 | 0 | 0 | 0 | 0 | 0 | 0 |
| 0 | 0 | 0 | 0 | 0 | 0 | 0 | 0 | 0 | 0 | 0 | 0 | 0 | 0 | 1 | 1 | 1 | 1 | 2 | 2 | 2 |
| 0 | 0 | 0 | 0 | 0 | 0 | 0 | 0 | 0 | 0 | 0 | 0 | 0 | 6 | 6 | 12 | 22 | 34 | 44 | 64 | 87 |
| 0 | 0 | 0 | 0 | 0 | 0 | 0 | 0 | 0 | 0 | 0 | 0 | 0 | 0 | 0 | 0 | 0 | 0 | 1 | 3 | 3 |
| 0 | 0 | 0 | 0 | 0 | 0 | 0 | 0 | 0 | 0 | 0 | 0 | 0 | 0 | 0 | 0 | 0 | 1 | 1 | 1 | 2 |
| 0 | 0 | 0 | 0 | 0 | 0 | 0 | 0 | 0 | 0 | 0 | 0 | 0 | 1 | 1 | 1 | 3 | 3 | 3 | 4 | 5 |
| 0 | 0 | 0 | 0 | 0 | 0 | 0 | 0 | 0 | 0 | 0 | 0 | 0 | 0 | 0 | 0 | 1 | 1 | 1 | 2 | 2 |
| 0 | 0 | 0 | 0 | 0 | 0 | 0 | 0 | 0 | 0 | 0 | 0 | 0 | 0 | 0 | 0 | 0 | 0 | 0 | 0 | 0 |
| 0 | 0 | 0 | 0 | 0 | 0 | 0 | 0 | 0 | 0 | 0 | 0 | 0 | 0 | 1 | 1 | 1 | 1 | 3 | 3 | 3 |
| 0 | 0 | 0 | 0 | 0 | 0 | 0 | 0 | 0 | 1 | 1 | 1 | 1 | 1 | 1 | 1 | 1 | 1 | 1 | 1 | 1 |
| 0 | 0 | 0 | 0 | 0 | 0 | 0 | 0 | 0 | 0 | 0 | 0 | 0 | 0 | 0 | 0 | 0 | 0 | 1 | 1 | 2 |
| 0 | 0 | 0 | 0 | 0 | 0 | 0 | 0 | 0 | 0 | 0 | 0 | 0 | 0 | 0 | 1 | 2 | 3 | 5 | 5 | 5 |
| 0 | 0 | 0 | 0 | 0 | 0 | 0 | 0 | 0 | 0 | 0 | 0 | 0 | 0 | 0 | 0 | 0 | 0 | 0 | 0 | 1 |
| 0 | 0 | 0 | 0 | 0 | 0 | 0 | 0 | 0 | 0 | 0 | 0 | 0 | 1 | 2 | 2 | 2 | 2 | 2 | 2 | 6 |
| 0 | 0 | 0 | 0 | 0 | 0 | 0 | 0 | 0 | 0 | 0 | 0 | 0 | 0 | 0 | 0 | 0 | 2 | 2 | 2 | 2 |
| 6 | 9 | 10 | 10 | 11 | 16 | 18 | 21 | 24 | 29 | 31 | 37 | 40 | 42 | 42 | 47 | 66 | 74 | 83 | 94 | 95 |
| 0 | 0 | 0 | 0 | 0 | 0 | 0 | 0 | 0 | 0 | 0 | 0 | 0 | 0 | 0 | 0 | 0 | 0 | 3 | 4 | 4 |
| 0 | 0 | 0 | 0 | 0 | 0 | 0 | 0 | 0 | 0 | 0 | 0 | 0 | 0 | 0 | 0 | 0 | 0 | 0 | 0 | 0 |
| 0 | 0 | 0 | 0 | 0 | 0 | 0 | 0 | 0 | 0 | 0 | 0 | 0 | 0 | 0 | 0 | 0 | 0 | 0 | 0 | 0 |

| 3/23/2020 | 3/24/2020 | 3/25/2020 | 3/26/2020 | 3/27/2020 | 3/28/2020 | 3/29/2020 | 3/30/2020 | 3/31/2020 | 4/1/2020 | 4/2/2020 | 4/3/2020 | 4/4/2020 | 4/5/2020 | 4/6/2020 | 4/7/2020 | 4/8/2020 | 4/9/2020 | 4/10/2020 | 4/11/2020 | 4/12/2020 |
|---|---|---|---|---|---|---|---|---|---|---|---|---|---|---|---|---|---|---|---|---|
| 0 | 1 | 1 | 1 | 2 | 2 | 3 | 3 | 3 | 3 | 3 | 3 | 5 | 6 | 6 | 6 | 7 | 7 | 7 | 8 | 8 |
| 0 | 0 | 0 | 1 | 3 | 3 | 4 | 6 | 13 | 17 | 17 | 21 | 26 | 31 | 32 | 39 | 48 | 48 | 58 | 60 | 61 |
| 0 | 2 | 2 | 3 | 3 | 5 | 6 | 7 | 8 | 10 | 12 | 12 | 14 | 16 | 16 | 18 | 18 | 21 | 24 | 27 | 29 |
| 2 | 5 | 6 | 8 | 13 | 15 | 17 | 20 | 24 | 29 | 32 | 41 | 52 | 64 | 65 | 73 | 80 | 89 | 97 | 108 | 115 |
| 35 | 44 | 57 | 64 | 107 | 119 | 137 | 151 | 172 | 172 | 203 | 270 | 276 | 319 | 343 | 374 | 442 | 492 | 541 | 609 | 651 |
| 7 | 11 | 19 | 24 | 31 | 44 | 47 | 51 | 69 | 80 | 97 | 111 | 126 | 140 | 150 | 179 | 193 | 226 | 250 | 274 | 290 |
| 10 | 12 | 19 | 21 | 27 | 33 | 34 | 36 | 69 | 85 | 112 | 132 | 165 | 189 | 206 | 277 | 335 | 380 | 444 | 494 | 554 |
| 2 | 2 | 3 | 3 | 4 | 5 | 9 | 9 | 9 | 11 | 12 | 15 | 21 | 22 | 24 | 24 | 27 | 32 | 38 | 47 | 50 |
| 0 | 0 | 0 | 2 | 2 | 5 | 6 | 7 | 10 | 11 | 12 | 14 | 14 | 14 | 15 | 16 | 19 | 23 | 32 | 33 | 35 |
| 17 | 20 | 23 | 29 | 46 | 56 | 60 | 71 | 85 | 101 | 144 | 170 | 195 | 221 | 254 | 296 | 323 | 371 | 419 | 446 | 461 |
| 31 | 38 | 47 | 60 | 65 | 79 | 84 | 103 | 130 | 154 | 176 | 198 | 208 | 219 | 319 | 351 | 371 | 413 | 420 | 432 | 447 |
| 0 | 0 | 0 | 0 | 0 | 0 | 0 | 0 | 1 | 1 | 2 | 3 | 4 | 4 | 5 | 5 | 5 | 6 | 8 | 9 | 9 |
| 0 | 0 | 1 | 1 | 3 | 3 | 4 | 6 | 7 | 9 | 11 | 11 | 14 | 22 | 25 | 26 | 27 | 29 | 31 | 34 | 41 |
| 0 | 0 | 0 | 3 | 4 | 5 | 6 | 7 | 9 | 9 | 9 | 10 | 10 | 10 | 13 | 15 | 18 | 24 | 25 | 27 | 27 |
| 12 | 16 | 19 | 26 | 34 | 47 | 65 | 73 | 99 | 141 | 157 | 210 | 243 | 274 | 307 | 380 | 462 | 528 | 596 | 677 | 720 |
| 7 | 12 | 14 | 17 | 24 | 31 | 32 | 35 | 49 | 65 | 78 | 102 | 116 | 127 | 139 | 173 | 203 | 245 | 300 | 350 | 350 |
| 2 | 2 | 2 | 3 | 4 | 5 | 6 | 8 | 9 | 10 | 13 | 17 | 21 | 23 | 25 | 27 | 39 | 44 | 52 | 56 | 57 |
| 4 | 4 | 5 | 5 | 8 | 9 | 9 | 11 | 18 | 20 | 31 | 37 | 40 | 45 | 59 | 65 | 73 | 79 | 90 | 94 | 97 |
| 34 | 46 | 73 | 83 | 119 | 137 | 151 | 185 | 239 | 273 | 310 | 370 | 409 | 477 | 512 | 582 | 652 | 702 | 755 | 806 | 840 |
| 9 | 11 | 15 | 25 | 35 | 44 | 48 | 56 | 89 | 122 | 154 | 192 | 216 | 231 | 260 | 356 | 433 | 503 | 599 | 692 | 756 |
| 3 | 4 | 4 | 5 | 5 | 5 | 10 | 18 | 18 | 31 | 36 | 42 | 53 | 91 | 103 | 124 | 138 | 171 | 171 | 206 | 262 |
| 0 | 0 | 0 | 0 | 1 | 1 | 3 | 5 | 5 | 7 | 7 | 9 | 10 | 10 | 10 | 12 | 14 | 16 | 17 | 19 | 19 |
| 15 | 24 | 43 | 60 | 92 | 111 | 132 | 184 | 259 | 337 | 417 | 478 | 540 | 617 | 727 | 845 | 959 | 1,076 | 1,278 | 1,391 | 1,487 |
| 1 | 1 | 2 | 4 | 4 | 5 | 10 | 10 | 17 | 18 | 22 | 22 | 24 | 29 | 34 | 39 | 50 | 57 | 57 | 67 | 70 |
| 3 | 5 | 8 | 8 | 9 | 10 | 12 | 13 | 14 | 18 | 19 | 19 | 24 | 34 | 39 | 53 | 58 | 77 | 96 | 109 | 110 |
| 1 | 2 | 5 | 7 | 8 | 13 | 16 | 20 | 22 | 26 | 29 | 29 | 35 | 51 | 59 | 67 | 76 | 82 | 82 | 93 | 98 |
| 0 | 0 | 0 | 1 | 1 | 1 | 2 | 4 | 5 | 5 | 5 | 5 | 6 | 6 | 6 | 6 | 6 | 6 | 6 | 6 | 6 |
| 0 | 0 | 1 | 3 | 3 | 4 | 6 | 8 | 9 | 16 | 19 | 21 | 24 | 33 | 33 | 46 | 65 | 65 | 74 | 80 | 81 |
| 0 | 0 | 0 | 0 | 1 | 1 | 1 | 3 | 3 | 3 | 3 | 3 | 3 | 3 | 3 | 4 | 4 | 5 | 6 | 7 | 8 |
| 0 | 0 | 0 | 0 | 2 | 2 | 2 | 2 | 3 | 4 | 5 | 6 | 8 | 8 | 8 | 10 | 14 | 15 | 17 | 17 | 17 |
| 1 | 1 | 1 | 1 | 2 | 2 | 3 | 3 | 3 | 4 | 5 | 7 | 9 | 9 | 9 | 13 | 18 | 21 | 22 | 23 | 23 |
| 27 | 44 | 62 | 81 | 108 | 140 | 161 | 198 | 267 | 355 | 537 | 646 | 846 | 917 | 1,003 | 1,232 | 1,504 | 1,700 | 1,932 | 2,187 | 2,350 |
| 0 | 0 | 1 | 1 | 1 | 2 | 2 | 4 | 5 | 6 | 7 | 10 | 11 | 12 | 12 | 13 | 16 | 17 | 19 | 20 | 26 |
| 4 | 4 | 6 | 10 | 29 | 14 | 15 | 17 | 26 | 32 | 38 | 43 | 45 | 46 | 58 | 71 | 80 | 85 | 102 | 112 | 114 |
| 156 | 231 | 332 | 424 | 546 | 779 | 895 | 1069 | 1307 | 1672 | 1957 | 2375 | 2820 | 3489 | 5940 | 5489 | 6240 | 7451 | 7844 | 8627 | 9162 |
| 6 | 8 | 10 | 15 | 19 | 25 | 29 | 39 | 55 | 65 | 81 | 91 | 102 | 119 | 142 | 167 | 193 | 213 | 231 | 247 | 248 |
| 2 | 3 | 7 | 7 | 8 | 15 | 17 | 17 | 23 | 30 | 34 | 38 | 42 | 46 | 51 | 67 | 79 | 80 | 88 | 94 | 96 |
| 5 | 8 | 10 | 11 | 12 | 13 | 13 | 16 | 18 | 18 | 21 | 22 | 26 | 27 | 29 | 33 | 38 | 44 | 48 | 51 | 52 |
| 2 | 7 | 10 | 16 | 22 | 34 | 38 | 49 | 63 | 74 | 90 | 102 | 136 | 150 | 162 | 240 | 310 | 338 | 416 | 494 | 507 |
| 0 | 0 | 0 | 0 | 0 | 2 | 3 | 4 | 8 | 10 | 12 | 14 | 17 | 25 | 27 | 30 | 35 | 47 | 49 | 56 | 63 |
| 5 | 7 | 7 | 9 | 13 | 15 | 16 | 18 | 22 | 26 | 31 | 34 | 40 | 44 | 48 | 51 | 63 | 67 | 72 | 80 | 82 |
| 1 | 1 | 1 | 1 | 1 | 1 | 1 | 1 | 1 | 2 | 2 | 2 | 2 | 2 | 4 | 6 | 6 | 6 | 6 | 6 | 6 |
| 2 | 2 | 3 | 3 | 6 | 6 | 7 | 13 | 23 | 23 | 32 | 36 | 42 | 44 | 65 | 72 | 79 | 94 | 98 | 100 | 101 |
| 8 | 11 | 12 | 18 | 23 | 27 | 34 | 38 | 41 | 58 | 70 | 90 | 105 | 127 | 140 | 154 | 177 | 199 | 226 | 254 | 271 |
| 1 | 1 | 1 | 2 | 2 | 2 | 3 | 5 | 5 | 7 | 7 | 7 | 8 | 8 | 13 | 13 | 13 | 15 | 17 | 18 | 18 |
| 6 | 9 | 13 | 14 | 14 | 17 | 25 | 27 | 34 | 41 | 46 | 46 | 52 | 54 | 63 | 75 | 109 | 121 | 121 | 130 | 149 |
| 5 | 7 | 8 | 8 | 10 | 12 | 12 | 12 | 13 | 16 | 17 | 17 | 20 | 22 | 23 | 23 | 23 | 23 | 24 | 25 | 27 |
| 110 | 123 | 132 | 147 | 175 | 189 | 189 | 195 | 195 | 246 | 261 | 284 | 310 | 337 | 371 | 394 | 420 | 445 | 475 | 491 | 508 |
| 5 | 5 | 6 | 8 | 13 | 13 | 13 | 14 | 16 | 24 | 31 | 37 | 56 | 68 | 77 | 92 | 99 | 111 | 128 | 137 | 144 |
| 0 | 0 | 0 | 0 | 0 | 0 | 1 | 1 | 1 | 2 | 2 | 2 | 2 | 3 | 4 | 4 | 4 | 5 | 5 | 6 | 8 |
| 0 | 0 | 0 | 0 | 0 | 0 | 0 | 0 | 0 | 0 | 0 | 0 | 0 | 0 | 0 | 0 | 0 | 0 | 0 | 0 | 0 |

| 4/13/2020 | 4/14/2020 | 4/15/2020 | 4/16/2020 | 4/17/2020 | 4/18/2020 | 4/19/2020 | 4/20/2020 | 4/21/2020 | 4/22/2020 | 4/23/2020 | 4/24/2020 | 4/25/2020 | 4/26/2020 | 4/27/2020 | 4/28/2020 | 4/29/2020 | 4/30/2020 | 5/1/2020 | 5/2/2020 | 5/3/2020 |
|---|---|---|---|---|---|---|---|---|---|---|---|---|---|---|---|---|---|---|---|---|
| 8 | 9 | 9 | 9 | 9 | 9 | 9 | 9 | 9 | 9 | 9 | 9 | 9 | 9 | 9 | 9 | 9 | 9 | 9 | 9 | 9 |
| 62 | 73 | 75 | 82 | 96 | 113 | 165 | 147 | 180 | 187 | 198 | 209 | 213 | 220 | 228 | 243 | 260 | 280 | 289 | 288 | 290 |
| 30 | 32 | 33 | 37 | 40 | 41 | 40 | 42 | 43 | 42 | 45 | 47 | 48 | 49 | 50 | 52 | 59 | 61 | 64 | 72 | 76 |
| 122 | 131 | 142 | 150 | 169 | 177 | 184 | 187 | 208 | 229 | 249 | 266 | 273 | 275 | 275 | 293 | 254 | 320 | 330 | 348 | 362 |
| 687 | 758 | 821 | 890 | 985 | 1,072 | 1,072 | 1,208 | 1,268 | 1,354 | 1,469 | 1,562 | 1,651 | 1,710 | 1,755 | 1,809 | 1,887 | 1,982 | 2,073 | 2,171 | 2,215 |
| 308 | 329 | 357 | 373 | 391 | 411 | 421 | 448 | 485 | 507 | 551 | 672 | 670 | 679 | 706 | 736 | 766 | 777 | 820 | 832 | 842 |
| 602 | 608 | 868 | 971 | 1,036 | 1,086 | 1,127 | 1,331 | 1,423 | 1,544 | 1,639 | 1,764 | 1,862 | 1,924 | 2,012 | 2,089 | 2,168 | 2,257 | 2,339 | 2,436 | 2,495 |
| 52 | 67 | 72 | 81 | 86 | 91 | 96 | 105 | 112 | 127 | 139 | 153 | 165 | 178 | 185 | 190 | 205 | 224 | 231 | 240 | 251 |
| 41 | 43 | 46 | 52 | 61 | 67 | 72 | 72 | 89 | 92 | 100 | 100 | 112 | 125 | 137 | 144 | 152 | 152 | 159 | 168 | 177 |
| 499 | 571 | 614 | 668 | 726 | 748 | 774 | 823 | 867 | 927 | 987 | 1,046 | 1,055 | 1,074 | 1,088 | 1,171 | 1,218 | 1,268 | 1,314 | 1,364 | 1,399 |
| 481 | 526 | 579 | 624 | 661 | 670 | 691 | 782 | 821 | 838 | 873 | 899 | 897 | 912 | 944 | 1,025 | 1,054 | 1,111 | 1,167 | 1,174 | 1,178 |
| 9 | 9 | 9 | 9 | 9 | 9 | 10 | 10 | 12 | 12 | 12 | 13 | 14 | 14 | 16 | 16 | 16 | 16 | 16 | 16 | 17 |
| 43 | 49 | 53 | 60 | 64 | 74 | 75 | 79 | 83 | 90 | 96 | 107 | 112 | 118 | 127 | 136 | 148 | 162 | 170 | 175 | 184 |
| 33 | 39 | 41 | 41 | 43 | 44 | 45 | 48 | 51 | 54 | 54 | 54 | 56 | 56 | 58 | 60 | 60 | 63 | 63 | 64 | 64 |
| 794 | 868 | 948 | 1,072 | 1,134 | 1,259 | 1,290 | 1,349 | 1,468 | 1,565 | 1,688 | 1,795 | 1,874 | 1,933 | 1,983 | 2,125 | 2,215 | 2,355 | 2,457 | 2,559 | 2,618 |
| 350 | 387 | 436 | 477 | 519 | 545 | 562 | 569 | 630 | 661 | 706 | 741 | 872 | 901 | 932 | 992 | 1,065 | 1,114 | 1,175 | 1,229 | 1,246 |
| 62 | 71 | 76 | 80 | 88 | 88 | 92 | 100 | 107 | 110 | 112 | 111 | 117 | 118 | 120 | 124 | 125 | 129 | 130 | 131 | 134 |
| 104 | 115 | 122 | 129 | 137 | 144 | 148 | 154 | 171 | 185 | 191 | 200 | 205 | 208 | 213 | 225 | 235 | 240 | 248 | 248 | 253 |
| 884 | 1,013 | 1,103 | 1,156 | 1,213 | 1,267 | 1,296 | 1,328 | 1,405 | 1,532 | 1,599 | 1,601 | 1,644 | 1,729 | 1,740 | 1,801 | 1,845 | 1,905 | 1,970 | 1,950 | 2,012 |
| 844 | 957 | 1,108 | 1,245 | 1,404 | 1,560 | 1,706 | 1,809 | 1,961 | 2,182 | 2,360 | 2,556 | 2,730 | 2,899 | 3,003 | 3,153 | 3,405 | 3,562 | 3,716 | 3,846 | 4,004 |
| 302 | 413 | 459 | 494 | 494 | 534 | 582 | 652 | 698 | 748 | 798 | 798 | 875 | 945 | 1,016 | 1,078 | 1,140 | 1,192 | 1,192 | 1,251 | 1,317 |
| 19 | 20 | 24 | 27 | 29 | 32 | 34 | 35 | 36 | 39 | 44 | 47 | 50 | 50 | 51 | 51 | 53 | 55 | 55 | 56 | 57 |
| 1,602 | 1,768 | 1,921 | 2,093 | 2,226 | 2,307 | 2,391 | 2,468 | 2,700 | 2,813 | 2,977 | 3,084 | 3,273 | 3,315 | 3,407 | 3,567 | 3,670 | 3,789 | 3,866 | 4,020 | 4,049 |
| 79 | 87 | 90 | 101 | 115 | 121 | 143 | 160 | 179 | 179 | 221 | 221 | 244 | 272 | 301 | 319 | 343 | 343 | 371 | 395 | 428 |
| 114 | 133 | 147 | 152 | 165 | 175 | 176 | 177 | 189 | 208 | 218 | 262 | 273 | 274 | 288 | 314 | 318 | 329 | 337 | 351 | 352 |
| 111 | 122 | 129 | 140 | 140 | 152 | 169 | 183 | 183 | 201 | 209 | 209 | 221 | 229 | 239 | 250 | 250 | 282 | 281 | 291 | 310 |
| 7 | 7 | 7 | 8 | 8 | 10 | 10 | 12 | 13 | 14 | 14 | 14 | 14 | 14 | 16 | 15 | 16 | 16 | 16 | 16 | 16 |
| 86 | 108 | 117 | 131 | 152 | 164 | 172 | 179 | 213 | 242 | 253 | 269 | 289 | 299 | 306 | 342 | 354 | 378 | 399 | 420 | 422 |
| 8 | 9 | 9 | 9 | 9 | 9 | 10 | 13 | 13 | 14 | 15 | 15 | 16 | 17 | 19 | 19 | 19 | 19 | 23 | 24 | 25 |
| 18 | 20 | 21 | 24 | 24 | 28 | 28 | 33 | 38 | 42 | 47 | 50 | 53 | 56 | 55 | 55 | 68 | 70 | 73 | 76 | 78 |
| 23 | 27 | 32 | 34 | 37 | 38 | 41 | 42 | 42 | 48 | 51 | 53 | 60 | 60 | 60 | 60 | 66 | 72 | 81 | 84 | 86 |
| 2,443 | 2,805 | 3,156 | 3,518 | 3,840 | 4,070 | 4,202 | 4,377 | 4,753 | 5,063 | 5,368 | 5,617 | 5,863 | 5,938 | 6,044 | 6,442 | 6,770 | 7,228 | 7,538 | 7,742 | 7,871 |
| 31 | 36 | 36 | 44 | 51 | 53 | 55 | 58 | 65 | 71 | 78 | 84 | 93 | 99 | 104 | 110 | 112 | 123 | 131 | 139 | 151 |
| 120 | 130 | 137 | 142 | 151 | 155 | 157 | 164 | 170 | 179 | 197 | 203 | 206 | 213 | 213 | 232 | 237 | 252 | 254 | 257 | 262 |
| 8843 | 9522 | 14306 | 14724 | 15759 | 16274 | 17027 | 18367 | 19103 | 19256 | 20255 | 20400 | 20985 | 21504 | 21883 | 22275 | 22759 | 23211 | 23673 | 23826 | 24560 |
| 268 | 324 | 361 | 389 | 418 | 451 | 471 | 509 | 557 | 610 | 656 | 690 | 711 | 728 | 753 | 799 | 937 | 975 | 1,002 | 1,021 | 1,038 |
| 99 | 108 | 123 | 131 | 136 | 139 | 140 | 143 | 164 | 170 | 179 | 188 | 194 | 195 | 197 | 207 | 214 | 222 | 230 | 238 | 238 |
| 53 | 55 | 58 | 64 | 70 | 72 | 74 | 75 | 78 | 78 | 83 | 86 | 87 | 91 | 92 | 99 | 101 | 103 | 104 | 109 | 109 |
| 524 | 584 | 647 | 707 | 756 | 836 | 1,112 | 1,204 | 1,564 | 1,622 | 1,421 | 1,492 | 1,537 | 1,550 | 1,597 | 1,716 | 2,195 | 2,292 | 2,354 | 2,418 | 2,444 |
| 73 | 80 | 87 | 114 | 118 | 137 | 150 | 155 | 181 | 186 | 189 | 202 | 215 | 226 | 233 | 239 | 251 | 266 | 279 | 296 | 320 |
| 87 | 97 | 107 | 109 | 116 | 119 | 120 | 124 | 135 | 140 | 150 | 157 | 166 | 174 | 177 | 192 | 203 | 244 | 256 | 267 | 275 |
| 6 | 6 | 6 | 7 | 7 | 7 | 7 | 7 | 8 | 9 | 9 | 10 | 10 | 11 | 11 | 11 | 13 | 17 | 21 | 21 | 21 |
| 109 | 124 | 134 | 141 | 141 | 145 | 147 | 152 | 157 | 165 | 170 | 168 | 176 | 181 | 184 | 188 | 192 | 199 | 204 | 209 | 210 |
| 287 | 318 | 364 | 393 | 428 | 453 | 477 | 495 | 517 | 543 | 561 | 593 | 623 | 648 | 663 | 690 | 732 | 782 | 816 | 847 | 867 |
| 18 | 20 | 21 | 22 | 23 | 25 | 28 | 32 | 34 | 35 | 38 | 39 | 41 | 41 | 44 | 45 | 46 | 46 | 46 | 49 | 50 |
| 154 | 195 | 208 | 231 | 231 | 258 | 300 | 324 | 324 | 372 | 410 | 436 | 436 | 458 | 492 | 522 | 552 | 581 | 581 | 616 | 684 |
| 28 | 29 | 30 | 35 | 35 | 38 | 38 | 38 | 40 | 40 | 43 | 44 | 46 | 46 | 47 | 47 | 47 | 49 | 50 | 51 | 52 |
| 508 | 546 | 567 | 583 | 603 | 624 | 635 | 652 | 682 | 692 | 711 | 723 | 738 | 749 | 765 | 786 | 801 | 814 | 824 | 830 | 834 |
| 154 | 170 | 182 | 197 | 205 | 211 | 220 | 230 | 242 | 246 | 257 | 262 | 266 | 272 | 281 | 300 | 308 | 316 | 327 | 334 | 339 |
| 9 | 10 | 12 | 13 | 16 | 18 | 18 | 24 | 26 | 29 | 31 | 32 | 33 | 34 | 37 | 38 | 40 | 44 | 47 | 50 | 50 |
| 1 | 1 | 2 | 2 | 2 | 2 | 2 | 2 | 6 | 6 | 7 | 7 | 7 | 7 | 7 | 7 | 7 | 7 | 7 | 7 | 7 |

| 5/4/2020 | 5/5/2020 | 5/6/2020 | 5/7/2020 | 5/8/2020 | 5/9/2020 | 5/10/2020 | 5/11/2020 | 5/12/2020 | 5/13/2020 | 5/14/2020 | 5/15/2020 | 5/16/2020 | 5/17/2020 | 5/18/2020 | 5/19/2020 | 5/20/2020 | 5/21/2020 | 5/22/2020 | 5/23/2020 | 5/24/2020 |
|---|---|---|---|---|---|---|---|---|---|---|---|---|---|---|---|---|---|---|---|---|
| 9 | 9 | 10 | 10 | 10 | 10 | 10 | 10 | 10 | 10 | 10 | 10 | 10 | 10 | 10 | 10 | 10 | 10 | 10 | 10 | 10 |
| 300 | 323 | 343 | 370 | 386 | 394 | 394 | 424 | 433 | 451 | 475 | 484 | 487 | 488 | 496 | 504 | 524 | 534 | 543 | 551 | 554 |
| 80 | 83 | 87 | 88 | 88 | 90 | 91 | 94 | 95 | 97 | 98 | 98 | 98 | 98 | 100 | 102 | 107 | 110 | 113 | 115 | 116 |
| 362 | 395 | 426 | 450 | 517 | 532 | 536 | 542 | 562 | 594 | 624 | 651 | 679 | 680 | 686 | 704 | 747 | 763 | 775 | 799 | 800 |
| 2,254 | 2,317 | 2,412 | 2,504 | 2,585 | 2,678 | 2,745 | 2,770 | 2,847 | 2,934 | 3,032 | 3,108 | 3,204 | 3,261 | 3,302 | 3,334 | 3,436 | 3,542 | 3,630 | 3,708 | 3,774 |
| 851 | 903 | 921 | 944 | 960 | 967 | 971 | 986 | 1,006 | 1,059 | 1,090 | 1,150 | 1,192 | 1,215 | 1,224 | 1,257 | 1,299 | 1,309 | 1,323 | 1,327 | 1,330 |
| 2,556 | 2,633 | 2,718 | 2,797 | 2,874 | 2,932 | 2,967 | 3,008 | 3,041 | 3,125 | 3,219 | 3,285 | 3,339 | 3,408 | 3,449 | 3,472 | 3,529 | 3,582 | 3,637 | 3,675 | 3,693 |
| 258 | 264 | 277 | 285 | 304 | 311 | 323 | 328 | 336 | 350 | 358 | 368 | 375 | 383 | 392 | 400 | 407 | 412 | 418 | 427 | 432 |
| 182 | 187 | 193 | 213 | 213 | 224 | 225 | 237 | 247 | 260 | 271 | 271 | 286 | 297 | 304 | 310 | 317 | 322 | 324 | 324 | 326 |
| 1,399 | 1,471 | 1,539 | 1,600 | 1,669 | 1,715 | 1,735 | 1,735 | 1,779 | 1,827 | 1,875 | 1,917 | 1,964 | 1,973 | 1,997 | 2,052 | 2,096 | 2,144 | 2,190 | 2,233 | 2,237 |
| 1,211 | 1,258 | 1,327 | 1,336 | 1,362 | 1,404 | 1,404 | 1,442 | 1,464 | 1,505 | 1,527 | 1,557 | 1,592 | 1,606 | 1,642 | 1,668 | 1,689 | 1,758 | 1,808 | 1,814 | 1,827 |
| 17 | 17 | 17 | 17 | 17 | 17 | 17 | 17 | 17 | 17 | 17 | 17 | 17 | 17 | 17 | 17 | 17 | 17 | 17 | 17 | 17 |
| 188 | 207 | 219 | 231 | 243 | 252 | 265 | 271 | 289 | 306 | 318 | 336 | 346 | 351 | 364 | 372 | 386 | 407 | 424 | 445 | 454 |
| 64 | 65 | 66 | 67 | 67 | 67 | 67 | 70 | 69 | 69 | 72 | 73 | 73 | 73 | 74 | 77 | 77 | 77 | 79 | 79 | 79 |
| 2,662 | 2,838 | 2,974 | 3,111 | 3,241 | 3,349 | 3,406 | 3,459 | 3,601 | 3,792 | 3,928 | 4,058 | 4,129 | 4,177 | 4,234 | 4,379 | 4,225 | 4,607 | 4,715 | 4,790 | 4,856 |
| 1,264 | 1,326 | 1,377 | 1,414 | 1,447 | 1,490 | 1,508 | 1,540 | 1,578 | 1,619 | 1,646 | 1,691 | 1,741 | 1,751 | 1,765 | 1,824 | 1,864 | 1,913 | 1,941 | 1,964 | 1,976 |
| 136 | 137 | 144 | 147 | 152 | 157 | 157 | 158 | 158 | 164 | 164 | 172 | 172 | 172 | 173 | 173 | 178 | 178 | 185 | 185 | 185 |
| 261 | 275 | 283 | 294 | 298 | 304 | 304 | 311 | 321 | 326 | 328 | 332 | 334 | 334 | 346 | 366 | 376 | 386 | 391 | 391 | 391 |
| 2,064 | 2,115 | 2,094 | 2,208 | 2,227 | 2,267 | 2,286 | 2,308 | 2,347 | 2,381 | 2,417 | 2,448 | 2,479 | 2,491 | 2,563 | 2,581 | 2,608 | 2,629 | 2,668 | 2,683 | 2,690 |
| 4,090 | 4,212 | 4,420 | 4,552 | 4,702 | 4,840 | 4,979 | 5,108 | 5,141 | 5,315 | 5,649 | 5,592 | 5,705 | 5,797 | 5,862 | 5,938 | 6,066 | 6,148 | 6,228 | 6,304 | 6,372 |
| 1,390 | 1,437 | 1,437 | 1,560 | 1,614 | 1,644 | 1,683 | 1,756 | 1,809 | 1,866 | 1,911 | 1,957 | 1,992 | 1,992 | 2,080 | 2,123 | 2,159 | 2,159 | 2,243 | 2,277 | 2,302 |
| 61 | 62 | 62 | 62 | 63 | 64 | 64 | 65 | 66 | 66 | 69 | 70 | 70 | 71 | 73 | 73 | 73 | 75 | 75 | 77 | 78 |
| 4,135 | 4,179 | 4,250 | 4,343 | 4,393 | 4,526 | 4,551 | 4,584 | 4,674 | 4,714 | 4,787 | 4,825 | 4,880 | 4,891 | 4,915 | 5,017 | 5,060 | 5,129 | 5,158 | 5,223 | 5,228 |
| 428 | 455 | 485 | 534 | 534 | 578 | 578 | 591 | 646 | 638 | 691 | 692 | 730 | 739 | 757 | 786 | 786 | 851 | 861 | 861 | 878 |
| 358 | 377 | 396 | 418 | 449 | 472 | 482 | 488 | 524 | 542 | 562 | 576 | 589 | 594 | 605 | 616 | 631 | 661 | 671 | 676 | 681 |
| 342 | 374 | 374 | 409 | 422 | 430 | 435 | 457 | 466 | 480 | 493 | 511 | 521 | 528 | 555 | 570 | 580 | 595 | 615 | 625 | 635 |
| 16 | 16 | 16 | 16 | 16 | 16 | 16 | 16 | 16 | 16 | 16 | 16 | 16 | 16 | 16 | 16 | 16 | 16 | 16 | 16 | 16 |
| 430 | 452 | 477 | 507 | 527 | 544 | 547 | 550 | 577 | 597 | 615 | 641 | 652 | 659 | 661 | 691 | 702 | 716 | 728 | 737 | 744 |
| 25 | 25 | 31 | 31 | 33 | 35 | 35 | 36 | 38 | 40 | 40 | 42 | 42 | 43 | 44 | 45 | 49 | 51 | 52 | 52 | 53 |
| 78 | 82 | 86 | 90 | 92 | 96 | 98 | 100 | 103 | 107 | 113 | 119 | 123 | 123 | 125 | 132 | 138 | 143 | 147 | 147 | 150 |
| 86 | 92 | 111 | 114 | 121 | 131 | 133 | 133 | 142 | 150 | 151 | 159 | 171 | 172 | 172 | 182 | 190 | 199 | 204 | 208 | 209 |
| 7,910 | 8,244 | 8,549 | 8,801 | 8,952 | 9,116 | 9,255 | 9,310 | 9,508 | 9,702 | 9,946 | 10,138 | 10,249 | 10,356 | 10,435 | 10,586 | 10,747 | 10,843 | 10,985 | 11,081 | 11,133 |
| 156 | 162 | 169 | 172 | 181 | 191 | 200 | 208 | 219 | 231 | 242 | 253 | 259 | 265 | 270 | 276 | 283 | 294 | 302 | 308 | 317 |
| 275 | 285 | 286 | 293 | 301 | 306 | 317 | 317 | 332 | 342 | 339 | 354 | 349 | 365 | 379 | 373 | 394 | 397 | 387 | 392 | 410 |
| 24717 | 25014 | 25779 | 26120 | 26359 | 26697 | 26923 | 27184 | 27282 | 27448 | 27617 | 27755 | 28097 | 28168 | 28302 | 28484 | 28609 | 28663 | 28802 | 28978 | 29046 |
| 1,056 | 1,135 | 1,225 | 1,271 | 1,306 | 1,331 | 1,341 | 1,357 | 1,436 | 1,483 | 1,534 | 1,581 | 1,610 | 1,625 | 1,657 | 1,720 | 1,781 | 1,836 | 1,872 | 1,956 | 1,969 |
| 238 | 239 | 253 | 254 | 254 | 261 | 272 | 269 | 269 | 280 | 283 | 288 | 283 | 288 | 293 | 293 | 293 | 300 | 300 | 305 | 310 |
| 109 | 113 | 115 | 121 | 124 | 127 | 127 | 130 | 130 | 134 | 137 | 137 | 137 | 137 | 138 | 140 | 144 | 145 | 147 | 147 | 148 |
| 2,458 | 3,012 | 3,106 | 3,416 | 3,616 | 3,688 | 3,707 | 3,731 | 3,806 | 3,943 | 4,218 | 4,342 | 4,403 | 4,418 | 4,418 | 4,418 | 4,767 | 4,418 | 5,096 | 5,124 | 5,139 |
| 341 | 355 | 370 | 388 | 399 | 418 | 422 | 430 | 444 | 462 | 468 | 479 | 489 | 499 | 506 | 532 | 538 | 556 | 579 | 597 | 608 |
| 283 | 296 | 305 | 316 | 320 | 330 | 331 | 346 | 355 | 362 | 371 | 380 | 380 | 385 | 391 | 399 | 407 | 416 | 419 | 425 | 435 |
| 21 | 24 | 29 | 31 | 31 | 34 | 34 | 34 | 39 | 39 | 43 | 44 | 44 | 44 | 44 | 46 | 46 | 48 | 50 | 50 | 50 |
| 219 | 226 | 239 | 237 | 241 | 242 | 243 | 251 | 264 | 273 | 287 | 290 | 295 | 298 | 301 | 305 | 310 | 314 | 316 | 330 | 337 |
| 884 | 906 | 948 | 973 | 1,004 | 1,049 | 1,088 | 1,100 | 1,133 | 1,158 | 1,216 | 1,272 | 1,305 | 1,336 | 1,347 | 1,369 | 1,419 | 1,440 | 1,480 | 1,506 | 1,519 |
| 56 | 58 | 58 | 61 | 66 | 67 | 67 | 72 | 75 | 75 | 77 | 78 | 79 | 80 | 85 | 91 | 92 | 94 | 96 | 97 | 98 |
| 684 | 713 | 713 | 812 | 812 | 827 | 850 | 891 | 927 | 955 | 977 | 1,002 | 1,009 | 1,014 | 1,041 | 1,074 | 1,099 | 1,136 | 1,159 | 1,171 | 1,208 |
| 52 | 52 | 52 | 53 | 53 | 53 | 53 | 53 | 53 | 53 | 53 | 53 | 53 | 54 | 54 | 54 | 54 | 54 | 54 | 54 | 54 |
| 841 | 862 | 870 | 891 | 905 | 921 | 931 | 945 | 962 | 975 | 983 | 992 | 1,000 | 1,001 | 1,002 | 1,031 | 1,037 | 1,044 | 1,050 | 1,057 | 1,061 |
| 340 | 353 | 362 | 374 | 384 | 398 | 400 | 409 | 418 | 421 | 434 | 445 | 453 | 453 | 459 | 467 | 481 | 487 | 496 | 507 | 510 |
| 50 | 50 | 51 | 51 | 52 | 53 | 54 | 57 | 58 | 59 | 62 | 64 | 65 | 67 | 68 | 68 | 69 | 71 | 72 | 72 | 72 |
| 7 | 7 | 7 | 7 | 7 | 7 | 7 | 7 | 7 | 7 | 7 | 7 | 7 | 8 | 10 | 10 | 11 | 12 | 12 | 12 | 12 |

| 5/25/2020 | 5/26/2020 | 5/27/2020 | 5/28/2020 | 5/29/2020 | 5/30/2020 | 5/31/2020 | 6/1/2020 | 6/2/2020 | 6/3/2020 | 6/4/2020 | 6/5/2020 | 6/6/2020 | 6/7/2020 | 6/8/2020 | 6/9/2020 | 6/10/2020 | 6/11/2020 | 6/12/2020 | 6/13/2020 | 6/14/2020 |
|---|---|---|---|---|---|---|---|---|---|---|---|---|---|---|---|---|---|---|---|---|
| 10 | 10 | 10 | 10 | 10 | 10 | 10 | 10 | 10 | 10 | 10 | 10 | 10 | 10 | 10 | 11 | 11 | 11 | 12 | 12 | 12 |
| 568 | 579 | 584 | 594 | 613 | 620 | 632 | 646 | 653 | 653 | 653 | 674 | 688 | 692 | 728 | 729 | 744 | 755 | 769 | 774 | 776 |
| 117 | 119 | 120 | 125 | 132 | 133 | 133 | 133 | 136 | 142 | 151 | 152 | 154 | 154 | 155 | 161 | 165 | 171 | 176 | 177 | 179 |
| 806 | 807 | 831 | 857 | 885 | 903 | 906 | 917 | 941 | 981 | 996 | 1,012 | 1,042 | 1,044 | 1,047 | 1,070 | 1,095 | 1,127 | 1,144 | 1,183 | 1,186 |
| 3,795 | 3,814 | 3,884 | 3,973 | 4,068 | 4,156 | 4,213 | 4,251 | 4,286 | 4,361 | 4,422 | 4,485 | 4,559 | 4,626 | 4,653 | 4,697 | 4,776 | 4,881 | 4,943 | 4,989 | 5,063 |
| 1,331 | 1,350 | 1,392 | 1,421 | 1,436 | 1,443 | 1,445 | 1,458 | 1,474 | 1,494 | 1,512 | 1,524 | 1,527 | 1,527 | 1,543 | 1,553 | 1,573 | 1,583 | 1,595 | 1,598 | 1,599 |
| 3,742 | 3,769 | 3,803 | 3,826 | 3,868 | 3,912 | 3,944 | 3,964 | 3,972 | 3,989 | 4,007 | 4,038 | 4,055 | 4,071 | 4,084 | 4,097 | 4,120 | 4,146 | 4,159 | 4,186 | 4,201 |
| 440 | 440 | 445 | 453 | 460 | 462 | 466 | 468 | 470 | 473 | 475 | 479 | 483 | 489 | 491 | 495 | 499 | 502 | 506 | 511 | 515 |
| 337 | 344 | 345 | 356 | 361 | 366 | 369 | 373 | 375 | 386 | 388 | 390 | 398 | 398 | 410 | 413 | 414 | 414 | 418 | 422 | 423 |
| 2,252 | 2,259 | 2,319 | 2,364 | 2,413 | 2,447 | 2,451 | 2,460 | 2,530 | 2,566 | 2,607 | 2,660 | 2,688 | 2,700 | 2,712 | 2,765 | 2,801 | 2,848 | 2,877 | 2,925 | 2,931 |
| 1,830 | 1,874 | 1,909 | 1,973 | 1,971 | 2,003 | 2,052 | 2,074 | 2,102 | 2,123 | 2,147 | 2,174 | 2,160 | 2,180 | 2,208 | 2,285 | 2,329 | 2,375 | 2,418 | 2,446 | 2,451 |
| 17 | 17 | 17 | 17 | 17 | 17 | 17 | 17 | 17 | 17 | 17 | 17 | 17 | 17 | 17 | 17 | 17 | 17 | 17 | 17 | 17 |
| 459 | 475 | 492 | 506 | 523 | 531 | 534 | 549 | 561 | 574 | 580 | 592 | 598 | 604 | 614 | 624 | 631 | 640 | 643 | 650 | 652 |
| 79 | 81 | 82 | 82 | 82 | 82 | 82 | 83 | 83 | 83 | 83 | 83 | 83 | 83 | 83 | 85 | 85 | 86 | 87 | 87 | 87 |
| 4,884 | 4,923 | 5,083 | 5,186 | 5,270 | 5,330 | 5,390 | 5,412 | 5,525 | 5,621 | 5,736 | 5,795 | 5,864 | 5,904 | 6,102 | 6,196 | 6,273 | 6,363 | 6,441 | 6,470 | 6,489 |
| 1,984 | 2,004 | 2,030 | 2,068 | 2,110 | 2,125 | 2,134 | 2,142 | 2,197 | 2,207 | 2,231 | 2,258 | 2,292 | 2,303 | 2,316 | 2,339 | 2,355 | 2,380 | 2,396 | 2,413 | 2,422 |
| 188 | 188 | 205 | 205 | 208 | 208 | 208 | 217 | 217 | 222 | 222 | 232 | 232 | 232 | 236 | 236 | 240 | 240 | 243 | 243 | 243 |
| 391 | 394 | 400 | 409 | 418 | 431 | 431 | 439 | 442 | 450 | 458 | 466 | 470 | 470 | 472 | 477 | 484 | 493 | 497 | 499 | 499 |
| 2,690 | 2,701 | 2,722 | 2,740 | 2,766 | 2,785 | 2,791 | 2,801 | 2,835 | 2,870 | 2,883 | 2,912 | 2,925 | 2,936 | 2,944 | 2,957 | 2,968 | 2,987 | 2,996 | 3,004 | 3,014 |
| 6,416 | 6,473 | 6,547 | 6,640 | 6,718 | 6,768 | 6,846 | 7,035 | 7,085 | 7,012 | 7,201 | 7,235 | 7,289 | 7,316 | 7,353 | 7,408 | 7,454 | 7,492 | 7,538 | 7,576 | 7,624 |
| 2,333 | 2,392 | 2,428 | 2,466 | 2,509 | 2,532 | 2,552 | 2,597 | 2,641 | 2,668 | 2,702 | 2,740 | 2,749 | 2,776 | 2,811 | 2,844 | 2,875 | 2,900 | 2,926 | 2,939 | 2,947 |
| 79 | 79 | 84 | 85 | 89 | 89 | 89 | 94 | 95 | 95 | 95 | 98 | 99 | 99 | 99 | 100 | 100 | 100 | 100 | 100 | 101 |
| 5,240 | 5,266 | 5,334 | 5,372 | 5,406 | 5,463 | 5,491 | 5,516 | 5,553 | 5,570 | 5,585 | 5,855 | 5,891 | 5,895 | 5,912 | 5,943 | 5,955 | 5,985 | 5,990 | 6,013 | 6,016 |
| 908 | 908 | 942 | 1,006 | 1,006 | 1,050 | 1,060 | 1,060 | 1,082 | 1,097 | 1,126 | 1,181 | 1,197 | 1,208 | 1,228 | 1,267 | 1,280 | 1,280 | 1,305 | 1,314 | 1,335 |
| 685 | 686 | 696 | 707 | 738 | 771 | 772 | 773 | 783 | 786 | 789 | 799 | 809 | 809 | 819 | 840 | 848 | 860 | 872 | 879 | 879 |
| 652 | 671 | 692 | 710 | 723 | 734 | 739 | 768 | 782 | 794 | 794 | 803 | 817 | 837 | 837 | 868 | 868 | 868 | 889 | 889 | 895 |
| 16 | 17 | 17 | 17 | 17 | 17 | 17 | 17 | 17 | 17 | 17 | 18 | 18 | 18 | 18 | 18 | 18 | 18 | 18 | 18 | 19 |
| 764 | 766 | 794 | 827 | 859 | 877 | 886 | 898 | 921 | 939 | 960 | 966 | 992 | 996 | 1,006 | 1,029 | 1,053 | 1,064 | 1,092 | 1,104 | 1,109 |
| 54 | 54 | 56 | 57 | 59 | 60 | 61 | 61 | 65 | 66 | 66 | 71 | 72 | 72 | 72 | 72 | 73 | 74 | 74 | 74 | 74 |
| 150 | 153 | 163 | 164 | 170 | 170 | 170 | 178 | 181 | 187 | 186 | 186 | 188 | 188 | 188 | 191 | 195 | 212 | 216 | 216 | 216 |
| 210 | 214 | 223 | 232 | 238 | 242 | 245 | 245 | 256 | 265 | 273 | 278 | 283 | 286 | 286 | 294 | 301 | 308 | 315 | 318 | 320 |
| 11,144 | 11,191 | 11,339 | 11,401 | 11,531 | 11,634 | 11,698 | 11,721 | 11,770 | 11,880 | 11,970 | 12,049 | 12,106 | 12,176 | 12,214 | 12,303 | 12,377 | 12,443 | 12,489 | 12,589 | 12,625 |
| 320 | 325 | 329 | 335 | 344 | 351 | 356 | 362 | 367 | 375 | 383 | 387 | 392 | 396 | 400 | 404 | 410 | 420 | 426 | 431 | 435 |
| 412 | 396 | 402 | 427 | 415 | 417 | 438 | 421 | 420 | 445 | 429 | 452 | 437 | 438 | 461 | 444 | 448 | 476 | 462 | 481 | 464 |
| 29193 | 29289 | 29392 | 29500 | 29535 | 29612 | 29737 | 29766 | 29886 | 29918 | 30011 | 30066 | 30152 | 30216 | 30280 | 30351 | 30409 | 30481 | 30511 | 30565 | 30605 |
| 1,987 | 2,002 | 2,044 | 2,098 | 2,131 | 2,149 | 2,155 | 2,206 | 2,258 | 2,299 | 2,339 | 2,355 | 2,370 | 2,377 | 2,404 | 2,421 | 2,457 | 2,490 | 2,508 | 2,554 | 2,557 |
| 312 | 316 | 317 | 319 | 328 | 333 | 333 | 338 | 343 | 343 | 347 | 350 | 354 | 354 | 359 | 361 | 360 | 366 | 367 | 369 | 359 |
| 148 | 148 | 148 | 151 | 151 | 153 | 153 | 154 | 157 | 159 | 159 | 161 | 163 | 164 | 164 | 169 | 169 | 171 | 173 | 174 | 176 |
| 5,152 | 5,265 | 5,373 | 5,464 | 5,537 | 5,555 | 5,567 | 5,667 | 5,742 | 5,817 | 5,886 | 5,931 | 5,943 | 5,953 | 6,014 | 6,062 | 6,113 | 6,162 | 6,211 | 6,211 | 6,243 |
| 608 | 634 | 655 | 677 | 693 | 711 | 718 | 720 | 732 | 742 | 756 | 772 | 772 | 772 | 799 | 808 | 812 | 823 | 833 | 833 | 833 |
| 440 | 446 | 466 | 470 | 483 | 487 | 494 | 500 | 501 | 518 | 525 | 538 | 545 | 546 | 557 | 568 | 575 | 588 | 593 | 599 | 600 |
| 50 | 50 | 54 | 54 | 59 | 62 | 62 | 62 | 62 | 62 | 64 | 65 | 65 | 65 | 65 | 68 | 69 | 73 | 74 | 75 | 75 |
| 339 | 344 | 354 | 357 | 361 | 366 | 366 | 369 | 396 | 403 | 416 | 423 | 432 | 433 | 436 | 450 | 456 | 461 | 468 | 472 | 475 |
| 1,527 | 1,536 | 1,562 | 1,601 | 1,626 | 1,648 | 1,672 | 1,678 | 1,698 | 1,734 | 1,767 | 1,788 | 1,819 | 1,830 | 1,836 | 1,853 | 1,885 | 1,920 | 1,939 | 1,957 | 1,976 |
| 101 | 103 | 106 | 107 | 110 | 112 | 113 | 114 | 113 | 117 | 119 | 120 | 121 | 121 | 126 | 128 | 131 | 134 | 139 | 139 | 139 |
| 1,236 | 1,281 | 1,338 | 1,358 | 1,370 | 1,375 | 1,392 | 1,407 | 1,428 | 1,445 | 1,453 | 1,460 | 1,472 | 1,477 | 1,496 | 1,514 | 1,520 | 1,534 | 1,541 | 1,546 | 1,552 |
| 54 | 54 | 54 | 55 | 55 | 55 | 55 | 55 | 55 | 55 | 55 | 55 | 55 | 55 | 55 | 55 | 55 | 55 | 55 | 55 | 55 |
| 1,070 | 1,078 | 1,095 | 1,106 | 1,111 | 1,118 | 1,118 | 1,124 | 1,129 | 1,135 | 1,138 | 1,149 | 1,153 | 1,159 | 1,161 | 1,176 | 1,190 | 1,194 | 1,204 | 1,213 | 1,217 |
| 514 | 517 | 539 | 550 | 568 | 588 | 592 | 595 | 607 | 616 | 626 | 633 | 645 | 647 | 646 | 661 | 671 | 682 | 695 | 697 | 698 |
| 73 | 74 | 74 | 74 | 74 | 75 | 75 | 76 | 78 | 78 | 79 | 84 | 84 | 84 | 84 | 84 | 85 | 86 | 88 | 88 | 88 |
| 12 | 13 | 14 | 15 | 15 | 16 | 17 | 17 | 17 | 17 | 17 | 17 | 17 | 17 | 17 | 17 | 18 | 18 | 18 | 18 | 18 |

| 6/15/2020 | 6/16/2020 | 6/17/2020 | 6/18/2020 | 6/19/2020 | 6/20/2020 | 6/21/2020 | 6/22/2020 | 6/23/2020 | 6/24/2020 | 6/25/2020 | 6/26/2020 | 6/27/2020 | 6/28/2020 | 6/29/2020 | 6/30/2020 | 7/1/2020 | 7/2/2020 | 7/3/2020 | 7/4/2020 | 7/5/2020 |
|---|---|---|---|---|---|---|---|---|---|---|---|---|---|---|---|---|---|---|---|---|
| 12 | 12 | 12 | 12 | 12 | 12 | 12 | 12 | 12 | 12 | 12 | 14 | 14 | 14 | 14 | 14 | 14 | 14 | 15 | 16 | 16 |
| 781 | 785 | 792 | 810 | 823 | 838 | 839 | 843 | 870 | 890 | 895 | 910 | 923 | 924 | 931 | 947 | 969 | 985 | 1,006 | 1,007 | 1,009 |
| 182 | 188 | 197 | 208 | 214 | 224 | 225 | 227 | 237 | 240 | 242 | 249 | 259 | 264 | 265 | 270 | 277 | 279 | 281 | 286 | 287 |
| 1,194 | 1,219 | 1,239 | 1,271 | 1,312 | 1,338 | 1,339 | 1,342 | 1,384 | 1,463 | 1,490 | 1,535 | 1,579 | 1,588 | 1,588 | 1,632 | 1,720 | 1,757 | 1,788 | 1,805 | 1,809 |
| 5,089 | 5,121 | 5,208 | 5,290 | 5,360 | 5,424 | 5,495 | 5,515 | 5,580 | 5,632 | 5,733 | 5,812 | 5,872 | 5,905 | 5,936 | 5,980 | 6,090 | 6,163 | 6,263 | 6,313 | 6,331 |
| 1,605 | 1,617 | 1,631 | 1,638 | 1,643 | 1,647 | 1,647 | 1,651 | 1,665 | 1,667 | 1,669 | 1,673 | 1,674 | 1,676 | 1,682 | 1,690 | 1,697 | 1,701 | 1,701 | 1,701 | 1,701 |
| 4,204 | 4,210 | 4,219 | 4,226 | 4,238 | 4,251 | 4,260 | 4,263 | 4,277 | 4,287 | 4,298 | 4,307 | 4,311 | 4,316 | 4,320 | 4,322 | 4,324 | 4,326 | 4,335 | 4,335 | 4,335 |
| 515 | 520 | 523 | 527 | 530 | 531 | 533 | 535 | 537 | 541 | 543 | 546 | 548 | 550 | 551 | 551 | 553 | 554 | 555 | 557 | 559 |
| 424 | 426 | 431 | 433 | 433 | 435 | 435 | 504 | 505 | 507 | 507 | 507 | 507 | 507 | 507 | 509 | 510 | 510 | 512 | 512 | 512 |
| 2,938 | 2,993 | 3,018 | 3,061 | 3,104 | 3,144 | 3,161 | 3,173 | 3,238 | 3,281 | 3,327 | 3,366 | 3,390 | 3,419 | 3,447 | 3,505 | 3,550 | 3,617 | 3,684 | 3,702 | 3,731 |
| 2,494 | 2,529 | 2,575 | 2,605 | 2,636 | 2,642 | 2,643 | 2,648 | 2,687 | 2,698 | 2,745 | 2,770 | 2,776 | 2,778 | 2,784 | 2,805 | 2,827 | 2,849 | 2,856 | 2,857 | 2,860 |
| 17 | 17 | 17 | 17 | 17 | 17 | 17 | 17 | 17 | 17 | 17 | 17 | 17 | 18 | 18 | 18 | 18 | 18 | 19 | 19 | 19 |
| 655 | 669 | 673 | 680 | 681 | 682 | 685 | 686 | 688 | 692 | 695 | 704 | 704 | 705 | 708 | 714 | 717 | 718 | 721 | 721 | 721 |
| 88 | 88 | 88 | 89 | 89 | 89 | 89 | 89 | 89 | 90 | 90 | 90 | 91 | 91 | 91 | 92 | 92 | 92 | 113 | 93 | 93 |
| 6,507 | 6,579 | 6,666 | 6,718 | 6,784 | 6,829 | 6,851 | 6,875 | 6,911 | 6,974 | 7,014 | 7,048 | 7,074 | 7,089 | 7,103 | 7,124 | 7,152 | 7,152 | 7,215 | 7,224 | 7,230 |
| 2,433 | 2,447 | 2,475 | 2,491 | 2,516 | 2,536 | 2,540 | 2,553 | 2,569 | 2,578 | 2,586 | 2,595 | 2,616 | 2,619 | 2,624 | 2,640 | 2,650 | 2,662 | 2,681 | 2,687 | 2,693 |
| 245 | 245 | 247 | 247 | 254 | 254 | 254 | 259 | 259 | 261 | 261 | 264 | 264 | 264 | 270 | 270 | 272 | 272 | 277 | 277 | 277 |
| 505 | 512 | 518 | 520 | 522 | 524 | 526 | 526 | 537 | 538 | 546 | 553 | 554 | 558 | 560 | 565 | 572 | 581 | 585 | 585 | 585 |
| 3,018 | 3,042 | 3,062 | 3,062 | 3,084 | 3,104 | 3,105 | 3,117 | 3,134 | 3,152 | 3,164 | 3,190 | 3,190 | 3,199 | 3,199 | 3,221 | 3,238 | 3,255 | 3,278 | 3,278 | 3,288 |
| 7,647 | 7,665 | 7,734 | 7,770 | 7,800 | 7,828 | 7,858 | 7,874 | 7,890 | 7,938 | 7,963 | 8,013 | 8,041 | 8,060 | 8,095 | 8,054 | 8,081 | 8,132 | 8,149 | 8,172 | 8,183 |
| 2,982 | 2,996 | 3,016 | 3,030 | 3,052 | 3,066 | 3,074 | 3,092 | 3,108 | 3,129 | 3,142 | 3,157 | 3,168 | 3,175 | 3,190 | 3,205 | 3,212 | 3,223 | 3,236 | 3,243 | 3,246 |
| 101 | 101 | 102 | 102 | 102 | 102 | 102 | 102 | 102 | 103 | 103 | 103 | 104 | 105 | 105 | 105 | 105 | 105 | 107 | 107 | 109 |
| 6,017 | 6,034 | 6,036 | 6,061 | 6,067 | 6,087 | 6,090 | 6,097 | 6,109 | 6,114 | 6,133 | 6,134 | 6,153 | 6,157 | 6,161 | 6,193 | 6,198 | 6,212 | 6,212 | 6,218 | 6,218 |
| 1,344 | 1,357 | 1,357 | 1,376 | 1,406 | 1,412 | 1,416 | 1,425 | 1,432 | 1,432 | 1,446 | 1,446 | 1,460 | 1,470 | 1,470 | 1,482 | 1,495 | 1,503 | 1,503 | 1,508 | 1,511 |
| 880 | 882 | 909 | 946 | 948 | 955 | 956 | 961 | 966 | 975 | 982 | 990 | 996 | 997 | 998 | 1,015 | 1,017 | 1,022 | 1,026 | 1,027 | 1,028 |
| 915 | 939 | 938 | 938 | 938 | 938 | 938 | 978 | 989 | 1,011 | 1,016 | 1,035 | 1,039 | 1,059 | 1,073 | 1,082 | 1,092 | 1,092 | 1,107 | 1,111 | 1,114 |
| 19 | 20 | 20 | 20 | 20 | 20 | 20 | 21 | 21 | 21 | 22 | 22 | 22 | 22 | 22 | 22 | 22 | 23 | 23 | 23 | 23 |
| 1,118 | 1,154 | 1,168 | 1,175 | 1,197 | 1,212 | 1,220 | 1,223 | 1,251 | 1,271 | 1,290 | 1,303 | 1,318 | 1,322 | 1,325 | 1,343 | 1,373 | 1,343 | 1,392 | 1,392 | 1,396 |
| 77 | 74 | 74 | 75 | 76 | 76 | 77 | 77 | 78 | 78 | 78 | 87 | 78 | 79 | 79 | 79 | 80 | 80 | 89 | 80 | 80 |
| 220 | 231 | 234 | 240 | 244 | 244 | 244 | 249 | 256 | 257 | 260 | 266 | 267 | 267 | 269 | 274 | 276 | 282 | 284 | 284 | 284 |
| 320 | 326 | 330 | 331 | 331 | 339 | 339 | 339 | 343 | 347 | 357 | 357 | 365 | 367 | 367 | 371 | 373 | 375 | 375 | 376 | 381 |
| 12,676 | 12,727 | 12,769 | 12,800 | 12,835 | 12,857 | 12,870 | 12,895 | 12,949 | 12,995 | 14,872 | 14,914 | 14,948 | 14,975 | 14,992 | 15,035 | 15,080 | 15,107 | 15,164 | 15,189 | 15,211 |
| 440 | 447 | 452 | 456 | 464 | 466 | 469 | 469 | 476 | 480 | 485 | 489 | 491 | 492 | 493 | 497 | 500 | 503 | 511 | 513 | 513 |
| 483 | 467 | 491 | 493 | 499 | 507 | 508 | 510 | 513 | 516 | 517 | 520 | 522 | 522 | 536 | 529 | 533 | 548 | 551 | 553 | 557 |
| 30645 | 30709 | 30750 | 30804 | 30824 | 30880 | 30927 | 30956 | 30992 | 31020 | 31066 | 31095 | 31134 | 31744 | 31769 | 31784 | 31810 | 31831 | 31855 | 31891 | 31906 |
| 2,573 | 2,597 | 2,611 | 2,633 | 2,667 | 2,697 | 2,700 | 2,704 | 2,735 | 2,755 | 2,772 | 2,788 | 2,804 | 2,807 | 2,818 | 2,863 | 2,876 | 2,886 | 2,903 | 2,907 | 2,911 |
| 371 | 371 | 376 | 377 | 379 | 380 | 381 | 382 | 382 | 382 | 375 | 390 | 393 | 401 | 401 | 401 | 406 | 407 | 407 | 398 | 416 |
| 180 | 182 | 183 | 187 | 188 | 189 | 190 | 192 | 192 | 195 | 197 | 202 | 202 | 202 | 204 | 207 | 208 | 209 | 209 | 213 | 215 |
| 6,276 | 6,319 | 6,361 | 6,399 | 6,419 | 6,423 | 6,426 | 6,426 | 6,518 | 6,557 | 6,579 | 6,603 | 6,606 | 6,614 | 6,649 | 6,687 | 6,712 | 6,746 | 6,740 | 6,749 | 6,754 |
| 851 | 865 | 876 | 885 | 894 | 894 | 894 | 903 | 906 | 912 | 920 | 927 | 927 | 927 | 946 | 950 | 956 | 959 | 960 | 960 | 960 |
| 602 | 607 | 617 | 621 | 639 | 644 | 653 | 659 | 673 | 683 | 693 | 694 | 711 | 716 | 720 | 739 | 766 | 784 | 793 | 813 | 820 |
| 75 | 77 | 78 | 78 | 81 | 81 | 81 | 81 | 83 | 84 | 87 | 88 | 91 | 91 | 91 | 91 | 93 | 97 | 97 | 97 | 97 |
| 483 | 493 | 497 | 509 | 515 | 524 | 526 | 531 | 542 | 556 | 567 | 577 | 584 | 584 | 592 | 604 | 609 | 620 | 633 | 637 | 646 |
| 1,983 | 2,029 | 2,062 | 2,105 | 2,140 | 2,165 | 2,182 | 2,192 | 2,220 | 2,249 | 2,296 | 2,324 | 2,366 | 2,393 | 2,403 | 2,424 | 2,481 | 2,525 | 2,575 | 2,608 | 2,637 |
| 144 | 146 | 149 | 155 | 156 | 155 | 158 | 160 | 163 | 165 | 168 | 168 | 170 | 168 | 171 | 173 | 175 | 183 | 182 | 181 | 185 |
| 1,570 | 1,583 | 1,586 | 1,602 | 1,607 | 1,611 | 1,620 | 1,645 | 1,661 | 1,675 | 1,700 | 1,724 | 1,732 | 1,740 | 1,763 | 1,763 | 1,786 | 1,816 | 1,845 | 1,853 | 1,853 |
| 55 | 55 | 55 | 56 | 56 | 56 | 56 | 56 | 56 | 56 | 56 | 56 | 56 | 56 | 56 | 56 | 56 | 56 | 56 | 56 | 56 |
| 1,221 | 1,231 | 1,226 | 1,245 | 1,255 | 1,265 | 1,270 | 1,276 | 1,284 | 1,293 | 1,300 | 1,304 | 1,310 | 1,310 | 1,320 | 1,332 | 1,339 | 1,342 | 1,352 | 1,354 | 1,359 |
| 700 | 709 | 718 | 725 | 736 | 751 | 751 | 751 | 757 | 764 | 773 | 773 | 784 | 784 | 784 | 791 | 793 | 800 | 803 | 803 | 803 |
| 88 | 88 | 88 | 88 | 88 | 88 | 89 | 90 | 92 | 92 | 92 | 92 | 93 | 93 | 93 | 93 | 93 | 93 | 93 | 94 | 95 |
| 18 | 18 | 18 | 18 | 20 | 20 | 20 | 20 | 20 | 20 | 20 | 20 | 20 | 20 | 20 | 20 | 20 | 20 | 20 | 20 | 20 |

| 7/6/2020 | 7/7/2020 | 7/8/2020 | 7/9/2020 | 7/10/2020 | 7/11/2020 | 7/12/2020 | 7/13/2020 | 7/14/2020 | 7/15/2020 | 7/16/2020 | 7/17/2020 | 7/18/2020 | 7/19/2020 | 7/20/2020 | 7/21/2020 | 7/22/2020 | 7/23/2020 | 7/24/2020 | 7/25/2020 | 7/26/2020 |
|---|---|---|---|---|---|---|---|---|---|---|---|---|---|---|---|---|---|---|---|---|
| 16 | 17 | 17 | 17 | 17 | 17 | 17 | 17 | 17 | 17 | 17 | 17 | 18 | 18 | 18 | 18 | 19 | 19 | 19 | 20 | 20 |
| 1,009 | 1,032 | 1,057 | 1,067 | 1,103 | 1,114 | 1,121 | 1,124 | 1,166 | 1,211 | 1,230 | 1,263 | 1,286 | 1,287 | 1,292 | 1,307 | 1,363 | 1,399 | 1,439 | 1,456 | 1,473 |
| 292 | 301 | 305 | 309 | 313 | 319 | 321 | 323 | 331 | 335 | 341 | 353 | 357 | 357 | 363 | 374 | 380 | 386 | 394 | 399 | 401 |
| 1,810 | 1,927 | 1,963 | 2,038 | 2,082 | 2,151 | 2,237 | 2,245 | 2,337 | 2,434 | 2,492 | 2,583 | 2,730 | 2,761 | 2,784 | 2,918 | 2,974 | 3,063 | 3,142 | 3,286 | 3,305 |
| 6,337 | 6,488 | 6,562 | 6,711 | 6,851 | 6,945 | 7,017 | 7,040 | 7,087 | 7,227 | 7,345 | 7,475 | 7,595 | 7,685 | 7,694 | 7,755 | 7,870 | 8,027 | 8,186 | 8,337 | 8,416 |
| 1,691 | 1,696 | 1,704 | 1,706 | 1,724 | 1,725 | 1,725 | 1,727 | 1,738 | 1,744 | 1,745 | 1,751 | 1,752 | 1,752 | 1,758 | 1,763 | 1,771 | 1,786 | 1,790 | 1,794 | 1,794 |
| 4,338 | 4,338 | 4,343 | 4,348 | 4,348 | 4,348 | 4,348 | 4,371 | 4,372 | 4,380 | 4,389 | 4,396 | 4,396 | 4,396 | 4,406 | 4,406 | 4,406 | 4,410 | 4,413 | 4,413 | 4,413 |
| 561 | 561 | 564 | 568 | 568 | 568 | 568 | 568 | 568 | 571 | 574 | 577 | 578 | 578 | 579 | 580 | 580 | 581 | 581 | 581 | 581 |
| 514 | 515 | 517 | 518 | 518 | 518 | 517 | 518 | 521 | 521 | 521 | 523 | 523 | 523 | 525 | 527 | 529 | 529 | 578 | 579 | 579 |
| 3,778 | 3,841 | 3,953 | 4,009 | 4,102 | 4,197 | 4,242 | 4,277 | 4,409 | 4,521 | 4,677 | 4,805 | 4,895 | 4,982 | 5,072 | 5,206 | 5,345 | 5,518 | 5,653 | 5,777 | 5,854 |
| 2,878 | 2,899 | 2,922 | 2,930 | 2,965 | 2,996 | 3,001 | 3,026 | 3,054 | 3,091 | 3,104 | 3,132 | 3,168 | 3,173 | 3,176 | 3,254 | 3,335 | 3,360 | 3,442 | 3,495 | 3,498 |
| 19 | 19 | 19 | 19 | 19 | 19 | 19 | 22 | 22 | 22 | 22 | 23 | 24 | 24 | 24 | 24 | 25 | 26 | 26 | 26 | 26 |
| 723 | 725 | 735 | 740 | 743 | 748 | 750 | 754 | 757 | 767 | 778 | 784 | 787 | 793 | 797 | 802 | 808 | 818 | 823 | 826 | 827 |
| 94 | 94 | 98 | 100 | 101 | 102 | 102 | 102 | 103 | 110 | 114 | 118 | 119 | 119 | 122 | 126 | 135 | 138 | 144 | 146 | 146 |
| 7,236 | 7,273 | 7,309 | 7,329 | 7,345 | 7,369 | 7,388 | 7,394 | 7,419 | 7,427 | 7,452 | 7,465 | 7,483 | 7,488 | 7,494 | 7,517 | 7,540 | 7,560 | 7,577 | 7,589 | 7,590 |
| 2,698 | 2,717 | 2,732 | 2,739 | 2,748 | 2,756 | 2,760 | 2,762 | 2,775 | 2,785 | 2,795 | 2,803 | 2,820 | 2,822 | 2,825 | 2,846 | 2,863 | 2,880 | 2,884 | 2,895 | 2,903 |
| 280 | 280 | 282 | 282 | 284 | 284 | 284 | 288 | 288 | 299 | 299 | 299 | 299 | 299 | 307 | 307 | 308 | 308 | 326 | 326 | 326 |
| 593 | 602 | 608 | 612 | 620 | 622 | 625 | 629 | 635 | 645 | 650 | 658 | 667 | 670 | 671 | 674 | 677 | 684 | 691 | 696 | 700 |
| 3,296 | 3,319 | 3,339 | 3,355 | 3,380 | 3,403 | 3,416 | 3,423 | 3,445 | 3,461 | 3,485 | 3,509 | 3,509 | 3,543 | 3,572 | 3,608 | 3,670 | 3,686 | 3,715 | 3,715 | 3,763 |
| 8,198 | 8,213 | 8,243 | 8,268 | 8,296 | 8,310 | 8,325 | 8,330 | 8,340 | 8,368 | 8,380 | 8,402 | 8,419 | 8,431 | 8,433 | 8,450 | 8,468 | 8,484 | 8,498 | 8,510 | 8,529 |
| 3,266 | 3,275 | 3,288 | 3,303 | 3,310 | 3,319 | 3,325 | 3,334 | 3,341 | 3,347 | 3,359 | 3,368 | 3,377 | 3,382 | 3,402 | 3,405 | 3,409 | 3,422 | 3,433 | 3,440 | 3,447 |
| 110 | 110 | 111 | 111 | 111 | 114 | 114 | 114 | 114 | 114 | 115 | 117 | 117 | 117 | 118 | 118 | 118 | 118 | 118 | 119 | 119 |
| 6,221 | 6,251 | 6,262 | 6,271 | 6,285 | 6,313 | 6,314 | 6,321 | 6,326 | 6,330 | 6,348 | 6,355 | 6,364 | 6,366 | 6,373 | 6,382 | 6,388 | 6,395 | 6,400 | 6,400 | 6,400 |
| 1,514 | 1,514 | 1,528 | 1,533 | 1,537 | 1,537 | 1,542 | 1,548 | 1,548 | 1,564 | 1,573 | 1,578 | 1,581 | 1,581 | 1,588 | 1,592 | 1,601 | 1,606 | 1,611 | 1,611 | 1,616 |
| 1,028 | 1,042 | 1,046 | 1,051 | 1,064 | 1,069 | 1,069 | 1,083 | 1,093 | 1,103 | 1,113 | 1,121 | 1,130 | 1,129 | 1,132 | 1,143 | 1,159 | 1,179 | 1,178 | 1,182 | 1,197 |
| 1,114 | 1,158 | 1,204 | 1,215 | 1,215 | 1,249 | 1,250 | 1,273 | 1,272 | 1,308 | 1,333 | 1,332 | 1,357 | 1,358 | 1,389 | 1,423 | 1,436 | 1,463 | 1,480 | 1,495 | 1,495 |
| 23 | 23 | 25 | 27 | 28 | 29 | 31 | 32 | 34 | 34 | 37 | 37 | 37 | 37 | 39 | 40 | 42 | 45 | 46 | 46 | 47 |
| 1,398 | 1,420 | 1,441 | 1,461 | 1,479 | 1,499 | 1,503 | 1,510 | 1,552 | 1,568 | 1,588 | 1,606 | 1,629 | 1,634 | 1,642 | 1,668 | 1,698 | 1,726 | 1,746 | 1,778 | 1,785 |
| 80 | 84 | 89 | 85 | 85 | 87 | 87 | 87 | 88 | 89 | 89 | 90 | 92 | 93 | 94 | 96 | 97 | 99 | 99 | 99 | 99 |
| 283 | 282 | 282 | 284 | 286 | 285 | 285 | 288 | 286 | 291 | 299 | 301 | 301 | 301 | 313 | 310 | 311 | 316 | 316 | 316 | 316 |
| 382 | 384 | 386 | 387 | 390 | 391 | 391 | 391 | 392 | 394 | 395 | 395 | 396 | 398 | 398 | 400 | 402 | 405 | 407 | 409 | 409 |
| 15,229 | 15,281 | 15,423 | 15,448 | 15,479 | 15,525 | 15,541 | 15,560 | 15,582 | 15,634 | 15,665 | 15,684 | 15,699 | 15,706 | 15,715 | 15,737 | 15,707 | 15,730 | 15,765 | 15,776 | 15,787 |
| 515 | 519 | 527 | 533 | 539 | 543 | 545 | 548 | 551 | 557 | 562 | 565 | 569 | 571 | 578 | 588 | 591 | 596 | 601 | 607 | 614 |
| 560 | 571 | 576 | 594 | 603 | 621 | 617 | 617 | 636 | 643 | 651 | 663 | 672 | 673 | 674 | 702 | 730 | 735 | 735 | 758 | 760 |
| 31928 | 31940 | 31968 | 31999 | 32015 | 32024 | 32069 | 32088 | 32109 | 32133 | 32139 | 32158 | 32179 | 32198 | 32203 | 32222 | 32263 | 32272 | 32289 | 32304 | 32320 |
| 2,927 | 2,970 | 2,991 | 3,006 | 3,032 | 3,036 | 3,058 | 3,064 | 3,069 | 3,075 | 3,103 | 3,112 | 3,132 | 3,174 | 3,189 | 3,219 | 3,235 | 3,256 | 3,297 | 3,297 | 3,307 |
| 415 | 416 | 424 | 428 | 432 | 432 | 441 | 424 | 424 | 430 | 432 | 438 | 451 | 451 | 451 | 461 | 473 | 477 | 483 | 495 | 496 |
| 215 | 220 | 224 | 230 | 232 | 232 | 234 | 237 | 244 | 247 | 249 | 254 | 257 | 260 | 262 | 269 | 271 | 273 | 282 | 286 | 289 |
| 6,787 | 6,812 | 6,848 | 6,880 | 6,897 | 6,904 | 6,911 | 6,931 | 6,957 | 6,973 | 6,992 | 7,007 | 7,015 | 7,018 | 7,038 | 7,063 | 7,079 | 7,101 | 7,114 | 7,118 | 7,122 |
| 960 | 969 | 971 | 974 | 976 | 976 | 976 | 984 | 984 | 987 | 988 | 990 | 990 | 990 | 995 | 996 | 997 | 1,001 | 1,002 | 1,002 | 1,002 |
| 827 | 846 | 884 | 905 | 929 | 951 | 961 | 972 | 993 | 998 | 1,070 | 1,096 | 1,135 | 1,155 | 1,164 | 1,221 | 1,285 | 1,334 | 1,385 | 1,465 | 1,491 |
| 97 | 98 | 98 | 101 | 107 | 109 | 109 | 109 | 109 | 111 | 115 | 116 | 116 | 118 | 118 | 118 | 119 | 121 | 122 | 122 | 123 |
| 653 | 665 | 685 | 710 | 723 | 738 | 741 | 749 | 767 | 783 | 796 | 815 | 838 | 843 | 847 | 871 | 888 | 925 | 938 | 964 | 967 |
| 2,655 | 2,715 | 2,813 | 2,918 | 3,013 | 3,112 | 3,192 | 3,235 | 3,322 | 3,432 | 3,561 | 3,735 | 3,865 | 3,958 | 4,020 | 4,151 | 4,348 | 4,521 | 4,717 | 4,885 | 5,038 |
| 190 | 199 | 201 | 209 | 213 | 215 | 216 | 220 | 226 | 234 | 235 | 243 | 243 | 243 | 249 | 258 | 263 | 267 | 274 | 273 | 275 |
| 1,881 | 1,905 | 1,937 | 1,958 | 1,962 | 1,966 | 1,968 | 1,977 | 1,992 | 2,007 | 2,013 | 2,025 | 2,027 | 2,031 | 2,048 | 2,051 | 2,054 | 2,067 | 2,075 | 2,078 | 2,082 |
| 56 | 56 | 56 | 56 | 56 | 56 | 56 | 56 | 56 | 56 | 56 | 56 | 56 | 56 | 56 | 56 | 56 | 56 | 56 | 56 | 56 |
| 1,370 | 1,384 | 1,394 | 1,409 | 1,424 | 1,424 | 1,438 | 1,399 | 1,404 | 1,421 | 1,427 | 1,434 | 1,444 | 1,447 | 1,453 | 1,465 | 1,468 | 1,482 | 1,495 | 1,494 | 1,501 |
| 803 | 812 | 814 | 816 | 821 | 828 | 820 | 827 | 833 | 834 | 838 | 840 | 850 | 851 | 853 | 866 | 872 | 885 | 885 | 898 | 899 |
| 95 | 95 | 95 | 95 | 95 | 96 | 96 | 97 | 97 | 98 | 99 | 100 | 100 | 100 | 100 | 101 | 102 | 103 | 103 | 103 | 103 |
| 20 | 20 | 21 | 21 | 21 | 21 | 21 | 21 | 22 | 22 | 24 | 24 | 24 | 24 | 24 | 25 | 25 | 25 | 25 | 25 | 25 |

| 7/27/2020 | 7/28/2020 | 7/29/2020 | 7/30/2020 | 7/31/2020 | 8/1/2020 | 8/2/2020 | 8/3/2020 | 8/4/2020 | 8/5/2020 | 8/6/2020 | 8/7/2020 | 8/8/2020 | 8/9/2020 | 8/10/2020 | 8/11/2020 | 8/12/2020 | 8/13/2020 | 8/14/2020 | 8/15/2020 | 8/16/2020 |
|---|---|---|---|---|---|---|---|---|---|---|---|---|---|---|---|---|---|---|---|---|
| 21 | 22 | 22 | 23 | 23 | 24 | 24 | 25 | 25 | 25 | 25 | 25 | 26 | 26 | 26 | 26 | 27 | 27 | 27 | 28 | 28 |
| 1,492 | 1,493 | 1,538 | 1,567 | 1,581 | 1,603 | 1,627 | 1,633 | 1,668 | 1,698 | 1,714 | 1,731 | 1,751 | 1,768 | 1,797 | 1,847 | 1,882 | 1,890 | 1,893 | 1,896 | 1,898 |
| 408 | 428 | 434 | 442 | 453 | 458 | 464 | 475 | 490 | 508 | 515 | 521 | 535 | 544 | 555 | 566 | 573 | 582 | 587 | 600 | 599 |
| 3,304 | 3,408 | 3,454 | 3,626 | 3,694 | 3,747 | 3,765 | 3,779 | 3,845 | 3,932 | 4,002 | 4,081 | 4,137 | 4,150 | 4,154 | 4,199 | 4,347 | 4,383 | 4,423 | 4,492 | 4,506 |
| 8,445 | 8,518 | 8,715 | 8,909 | 9,005 | 9,224 | 9,356 | 9,388 | 9,501 | 9,703 | 9,869 | 10,011 | 10,189 | 10,293 | 10,359 | 10,468 | 10,648 | 10,808 | 10,996 | 11,147 | 11,224 |
| 1,799 | 1,807 | 1,822 | 1,822 | 1,838 | 1,844 | 1,844 | 1,844 | 1,849 | 1,851 | 1,852 | 1,857 | 1,857 | 1,858 | 1,863 | 1,875 | 1,875 | 1,882 | 1,888 | 1,896 | 1,896 |
| 4,418 | 4,423 | 4,425 | 4,431 | 4,432 | 4,432 | 4,432 | 4,437 | 4,437 | 4,437 | 4,437 | 4,441 | 4,441 | 4,441 | 4,444 | 4,444 | 4,450 | 4,450 | 4,453 | 4,453 | 4,453 |
| 582 | 583 | 584 | 584 | 585 | 585 | 586 | 586 | 587 | 587 | 587 | 589 | 590 | 591 | 591 | 593 | 593 | 594 | 594 | 597 | 597 |
| 580 | 581 | 581 | 581 | 585 | 585 | 585 | 585 | 587 | 587 | 587 | 590 | 591 | 591 | 591 | 591 | 593 | 593 | 593 | 593 | 593 |
| 5,931 | 6,117 | 6,333 | 6,586 | 6,843 | 7,022 | 7,084 | 7,292 | 7,402 | 7,627 | 7,747 | 7,927 | 8,109 | 8,186 | 8,359 | 8,553 | 8,765 | 8,913 | 9,141 | 9,345 | 9,452 |
| 3,509 | 3,563 | 3,642 | 3,671 | 3,752 | 3,825 | 3,840 | 3,842 | 3,921 | 3,984 | 4,026 | 4,117 | 4,186 | 4,199 | 4,229 | 4,351 | 4,456 | 4,538 | 4,573 | 4,669 | 4,702 |
| 26 | 26 | 26 | 26 | 26 | 26 | 26 | 26 | 27 | 27 | 29 | 30 | 31 | 31 | 34 | 34 | 38 | 38 | 40 | 40 | 40 |
| 834 | 839 | 845 | 857 | 867 | 871 | 876 | 883 | 887 | 899 | 908 | 913 | 925 | 930 | 933 | 940 | 951 | 959 | 966 | 973 | 975 |
| 152 | 160 | 173 | 177 | 189 | 197 | 197 | 200 | 210 | 217 | 223 | 229 | 235 | 237 | 239 | 246 | 246 | 251 | 265 | 269 | 269 |
| 7,608 | 7,638 | 7,654 | 7,670 | 7,692 | 7,700 | 7,714 | 7,723 | 7,742 | 7,770 | 7,791 | 7,822 | 7,840 | 7,845 | 7,846 | 7,866 | 7,881 | 7,905 | 7,932 | 7,937 | 7,955 |
| 2,906 | 2,924 | 2,932 | 2,946 | 2,965 | 2,971 | 2,975 | 2,980 | 2,996 | 3,007 | 3,013 | 3,023 | 3,036 | 3,041 | 3,044 | 3,069 | 3,086 | 3,105 | 3,113 | 3,128 | 3,133 |
| 335 | 335 | 349 | 349 | 358 | 358 | 358 | 365 | 365 | 368 | 368 | 380 | 380 | 380 | 387 | 387 | 395 | 395 | 402 | 402 | 402 |
| 709 | 719 | 724 | 731 | 735 | 740 | 742 | 744 | 751 | 752 | 760 | 764 | 772 | 773 | 775 | 783 | 790 | 796 | 804 | 810 | 813 |
| 3,786 | 3,812 | 3,883 | 3,925 | 3,949 | 3,949 | 4,007 | 4,024 | 4,051 | 4,096 | 4,146 | 4,207 | 4,207 | 4,263 | 4,287 | 4,313 | 4,361 | 4,402 | 4,430 | 4,430 | 4,507 |
| 8,536 | 8,551 | 8,580 | 8,595 | 8,609 | 8,626 | 8,638 | 8,648 | 8,657 | 8,659 | 8,691 | 8,709 | 8,721 | 8,735 | 8,741 | 8,751 | 8,769 | 8,790 | 8,804 | 8,813 | 8,837 |
| 3,458 | 3,478 | 3,488 | 3,493 | 3,506 | 3,515 | 3,523 | 3,530 | 3,536 | 3,551 | 3,565 | 3,577 | 3,585 | 3,591 | 3,604 | 3,612 | 3,620 | 3,631 | 3,636 | 3,639 | 3,641 |
| 121 | 121 | 121 | 123 | 123 | 123 | 123 | 123 | 124 | 124 | 124 | 125 | 125 | 125 | 126 | 126 | 126 | 126 | 127 | 127 | 127 |
| 6,405 | 6,421 | 6,422 | 6,443 | 6,450 | 6,457 | 6,457 | 6,463 | 6,471 | 6,478 | 6,506 | 6,524 | 6,520 | 6,519 | 6,526 | 6,533 | 6,539 | 6,555 | 6,566 | 6,586 | 6,592 |
| 1,620 | 1,629 | 1,634 | 1,640 | 1,646 | 1,654 | 1,654 | 1,660 | 1,670 | 1,677 | 1,681 | 1,689 | 1,698 | 1,701 | 1,707 | 1,707 | 1,726 | 1,739 | 1,745 | 1,752 | 1,759 |
| 1,201 | 1,213 | 1,220 | 1,233 | 1,243 | 1,253 | 1,253 | 1,255 | 1,266 | 1,273 | 1,280 | 1,301 | 1,301 | 1,307 | 1,307 | 1,312 | 1,323 | 1,325 | 1,335 | 1,346 | 1,367 |
| 1,501 | 1,563 | 1,611 | 1,611 | 1,693 | 1,693 | 1,711 | 1,753 | 1,753 | 1,804 | 1,825 | 1,874 | 1,874 | 1,896 | 1,912 | 1,944 | 1,989 | 2,011 | 2,043 | 2,080 | 2,084 |
| 47 | 52 | 55 | 58 | 60 | 61 | 61 | 64 | 65 | 66 | 70 | 74 | 75 | 75 | 75 | 79 | 79 | 81 | 81 | 82 | 82 |
| 1,790 | 1,820 | 1,865 | 1,903 | 1,924 | 1,964 | 1,969 | 1,982 | 2,010 | 2,050 | 2,092 | 2,134 | 2,160 | 2,168 | 2,172 | 2,204 | 2,249 | 2,287 | 2,313 | 2,343 | 2,347 |
| 100 | 102 | 103 | 103 | 103 | 105 | 105 | 107 | 108 | 109 | 110 | 112 | 112 | 113 | 118 | 120 | 120 | 121 | 121 | 125 | 126 |
| 317 | 321 | 324 | 328 | 332 | 332 | 332 | 332 | 332 | 335 | 340 | 345 | 345 | 345 | 348 | 351 | 356 | 360 | 361 | 361 | 361 |
| 409 | 409 | 411 | 415 | 415 | 416 | 417 | 417 | 418 | 418 | 419 | 419 | 419 | 419 | 419 | 419 | 420 | 422 | 423 | 423 | 423 |
| 15,804 | 15,825 | 15,798 | 15,809 | 15,819 | 15,830 | 15,836 | 15,846 | 15,857 | 15,842 | 15,849 | 15,860 | 15,869 | 15,874 | 15,878 | 15,890 | 15,885 | 15,893 | 15,903 | 15,876 | 15,903 |
| 619 | 626 | 632 | 635 | 642 | 651 | 654 | 655 | 658 | 667 | 669 | 675 | 681 | 685 | 690 | 693 | 695 | 697 | 703 | 711 | 714 |
| 765 | 785 | 806 | 827 | 856 | 858 | 858 | 874 | 889 | 917 | 927 | 947 | 977 | 985 | 991 | 1,009 | 1,024 | 1,058 | 1,073 | 1,097 | 1,100 |
| 32329 | 32338 | 32355 | 32368 | 32377 | 32395 | 32410 | 32419 | 32429 | 32431 | 32438 | 32449 | 32455 | 32465 | 32472 | 32487 | 32497 | 32509 | 32512 | 32526 | 32535 |
| 3,344 | 3,382 | 3,422 | 3,442 | 3,489 | 3,515 | 3,529 | 3,539 | 3,570 | 3,596 | 3,618 | 3,652 | 3,668 | 3,669 | 3,673 | 3,708 | 3,734 | 3,755 | 3,784 | 3,824 | 3,826 |
| 496 | 503 | 523 | 536 | 541 | 549 | 550 | 551 | 566 | 583 | 593 | 599 | 603 | 603 | 605 | 618 | 627 | 637 | 644 | 657 | 657 |
| 289 | 303 | 311 | 316 | 322 | 325 | 326 | 328 | 333 | 338 | 339 | 348 | 355 | 356 | 357 | 368 | 375 | 383 | 385 | 386 | 393 |
| 7,146 | 7,162 | 7,176 | 7,189 | 7,204 | 7,204 | 7,209 | 7,209 | 7,232 | 7,244 | 7,282 | 7,297 | 7,313 | 7,314 | 7,317 | 7,352 | 7,385 | 7,409 | 7,445 | 7,465 | 7,468 |
| 1,004 | 1,005 | 1,007 | 1,007 | 1,007 | 1,007 | 1,007 | 1,010 | 1,011 | 1,012 | 1,014 | 1,014 | 1,014 | 1,014 | 1,015 | 1,016 | 1,018 | 1,019 | 1,021 | 1,021 | 1,021 |
| 1,506 | 1,565 | 1,615 | 1,667 | 1,712 | 1,751 | 1,777 | 1,793 | 1,847 | 1,894 | 1,943 | 1,962 | 2,007 | 2,031 | 2,049 | 2,098 | 2,144 | 2,186 | 2,204 | 2,260 | 2,269 |
| 123 | 123 | 129 | 129 | 130 | 134 | 135 | 135 | 136 | 137 | 141 | 144 | 146 | 146 | 146 | 146 | 147 | 148 | 150 | 152 | 153 |
| 978 | 999 | 1,020 | 1,033 | 1,060 | 1,067 | 1,073 | 1,092 | 1,117 | 1,144 | 1,186 | 1,206 | 1,215 | 1,223 | 1,233 | 1,271 | 1,289 | 1,313 | 1,326 | 1,345 | 1,366 |
| 5,713 | 5,877 | 6,190 | 6,274 | 6,569 | 6,837 | 6,837 | 7,016 | 7,261 | 7,497 | 7,803 | 8,096 | 8,343 | 8,459 | 8,490 | 8,710 | 9,034 | 9,289 | 9,602 | 9,840 | 9,983 |
| 283 | 292 | 293 | 303 | 307 | 311 | 311 | 319 | 324 | 328 | 331 | 336 | 336 | 337 | 347 | 350 | 353 | 357 | 361 | 362 | 363 |
| 2,095 | 2,125 | 2,141 | 2,174 | 2,215 | 2,218 | 2,218 | 2,244 | 2,274 | 2,299 | 2,317 | 2,322 | 2,326 | 2,327 | 2,344 | 2,352 | 2,364 | 2,370 | 2,381 | 2,382 | 2,385 |
| 56 | 56 | 56 | 57 | 57 | 57 | 57 | 57 | 57 | 57 | 58 | 58 | 58 | 58 | 58 | 58 | 58 | 58 | 58 | 58 | 58 |
| 1,518 | 1,548 | 1,554 | 1,564 | 1,564 | 1,592 | 1,596 | 1,600 | 1,619 | 1,624 | 1,653 | 1,672 | 1,688 | 1,688 | 1,697 | 1,716 | 1,724 | 1,736 | 1,755 | 1,766 | 1,781 |
| 900 | 913 | 918 | 926 | 941 | 954 | 955 | 956 | 968 | 977 | 985 | 997 | 1,003 | 1,005 | 1,005 | 1,013 | 1,018 | 1,025 | 1,032 | 1,045 | 1,046 |
| 106 | 111 | 112 | 115 | 116 | 116 | 117 | 117 | 124 | 124 | 124 | 127 | 131 | 139 | 141 | 147 | 153 | 153 | 157 | 160 | 160 |
| 25 | 26 | 26 | 26 | 26 | 26 | 26 | 27 | 27 | 27 | 27 | 28 | 28 | 28 | 28 | 29 | 29 | 30 | 30 | 30 | 30 |

| 8/17/2020 | 8/18/2020 | 8/19/2020 | 8/20/2020 | 8/21/2020 | 8/22/2020 | 8/23/2020 |
|---|---|---|---|---|---|---|
| 28 | 29 | 29 | 29 | 30 | 31 | 32 |
| 1,925 | 1,936 | 1,944 | 1,974 | 1,996 | 2,011 | 2,017 |
| 603 | 619 | 631 | 641 | 663 | 674 | 687 |
| 4,506 | 4,529 | 4,634 | 4,684 | 4,688 | 4,756 | 4,771 |
| 11,242 | 11,342 | 11,523 | 11,686 | 11,821 | 11,988 | 12,134 |
| 1,896 | 1,899 | 1,900 | 1,903 | 1,910 | 1,918 | 1,918 |
| 4,456 | 4,456 | 4,457 | 4,458 | 4,460 | 4,460 | 4,460 |
| 597 | 599 | 600 | 601 | 602 | 604 | 604 |
| 593 | 595 | 595 | 600 | 600 | 600 | 604 |
| 9,539 | 9,758 | 9,932 | 10,049 | 10,168 | 10,274 | 10,325 |
| 4,727 | 4,794 | 4,849 | 4,904 | 4,998 | 5,092 | 5,132 |
| 40 | 41 | 43 | 45 | 46 | 47 | 47 |
| 987 | 989 | 1,005 | 1,016 | 1,021 | 1,030 | 1,036 |
| 273 | 282 | 291 | 298 | 304 | 306 | 307 |
| 7,967 | 7,993 | 8,017 | 8,044 | 8,066 | 8,083 | 8,089 |
| 3,135 | 3,165 | 3,180 | 3,191 | 3,208 | 3,218 | 3,220 |
| 405 | 405 | 411 | 411 | 419 | 419 | 419 |
| 818 | 830 | 842 | 856 | 864 | 872 | 881 |
| 4,526 | 4,554 | 4,609 | 4,637 | 4,687 | 4,687 | 4,746 |
| 8,842 | 8,848 | 8,876 | 8,888 | 8,901 | 8,921 | 8,921 |
| 3,650 | 3,661 | 3,669 | 3,674 | 3,685 | 3,691 | 3,694 |
| 127 | 127 | 128 | 129 | 130 | 131 | 131 |
| 6,592 | 6,608 | 6,618 | 6,634 | 6,634 | 6,655 | 6,659 |
| 1,758 | 1,767 | 1,792 | 1,800 | 1,799 | 1,813 | 1,817 |
| 1,396 | 1,402 | 1,414 | 1,417 | 1,419 | 1,425 | 1,426 |
| 2,128 | 2,128 | 2,190 | 2,190 | 2,237 | 2,240 | 2,248 |
| 84 | 84 | 84 | 89 | 89 | 90 | 91 |
| 2,348 | 2,396 | 2,431 | 2,465 | 2,494 | 2,521 | 2,531 |
| 128 | 130 | 130 | 132 | 135 | 136 | 137 |
| 362 | 368 | 371 | 373 | 376 | 376 | 378 |
| 423 | 424 | 427 | 428 | 428 | 429 | 429 |
| 15,916 | 15,925 | 15,926 | 15,932 | 15,941 | 15,943 | 15,946 |
| 718 | 723 | 729 | 734 | 739 | 743 | 745 |
| 1,105 | 1,130 | 1,162 | 1,200 | 1,215 | 1,227 | 1,227 |
| 32548 | 32553 | 32553 | 32559 | 32567 | 32582 | 32592 |
| 3,832 | 3,871 | 3,907 | 3,929 | 3,955 | 3,975 | 3,978 |
| 665 | 682 | 699 | 709 | 715 | 725 | 726 |
| 396 | 397 | 408 | 412 | 414 | 417 | 417 |
| 7,468 | 7,499 | 7,523 | 7,523 | 7,558 | 7,576 | 7,578 |
| 1,023 | 1,024 | 1,027 | 1,028 | 1,030 | 1,030 | 1,030 |
| 2,288 | 2,343 | 2,360 | 2,401 | 2,459 | 2,493 | 2,504 |
| 153 | 154 | 155 | 157 | 159 | 160 | 161 |
| 1,387 | 1,426 | 1,452 | 1,488 | 1,549 | 1,563 | 1,567 |
| 10,034 | 10,250 | 10,559 | 10,793 | 11,051 | 11,266 | 11,370 |
| 365 | 371 | 377 | 382 | 383 | 385 | 385 |
| 2,397 | 2,410 | 2,427 | 2,436 | 2,443 | 2,443 | 2,471 |
| 58 | 58 | 58 | 58 | 58 | 58 | 58 |
| 1,785 | 1,809 | 1,822 | 1,837 | 1,850 | 1,857 | 1,863 |
| 1,046 | 1,059 | 1,067 | 1,074 | 1,075 | 1,089 | 1,089 |
| 160 | 164 | 166 | 166 | 170 | 176 | 178 |
| 33 | 34 | 34 | 34 | 37 | 37 | 37 |
